# Personalized digital behavior interventions increase short term physical activity: a randomized control crossover trial substudy of the MyHeart Counts Cardiovascular Health study

**DOI:** 10.1101/2023.04.09.23287650

**Authors:** Ali Javed, Daniel Seung Kim, Steven G Hershman, Anna Shcherbina, Anders Johnson, Alexander Tolas, Jack W O’Sullivan, Michael V McConnell, Laura Lazzeroni, Abby C King, Jeffrey W Christle, Marily Oppezzo, C. Mikael Mattsson, Robert A Harrington, Matthew T Wheeler, Euan A Ashley

## Abstract

**Background:** Physical activity is strongly protective against the development of chronic diseases associated with aging. We previously demonstrated that digital interventions delivered through a smartphone app can increase short-term physical activity. Our randomized crossover trial has continued to digitally enroll participants, allowing increasing statistical power for greater precision in subsequent analyses.

**Methods:** We offered enrollment to adults aged ≥18 years with access to an iPhone and the MyHeart Counts app. After completion of a 1-week baseline period, e-consented participants were randomly allocated to four 7-day interventions. Interventions consisted of: 1) daily personalized e-coaching based on the individual’s baseline activity patterns, 2) daily prompts to complete 10,000 steps, 3) hourly prompts to stand following inactivity, and 4) daily instructions to read guidelines from the American Heart Association website. The trial was completed in a free-living setting, where neither the participants or investigators were blinded to the intervention. The primary outcome was change in mean daily step count from baseline for each of the four interventions, assessed in a modified intention-to-treat analysis. This trial is registered with ClinicalTrials.gov, NCT03090321.

**Findings:** Between January 1, 2017 and April 1, 2022, 4500 participants consented to enroll in the trial, of whom 2458 completed 7-days of baseline monitoring (mean daily steps 4232±73) and at least one day of one of the four interventions. The greater statistical power afforded by continued passive enrollment revealed that e-coaching prompts, tailored to an individual, increased step count significantly more than other interventions (402±71 steps, *P=*7.1×10^−8^).

**Interpretation:** Digital studies can continuously recruit participants in a cost-effective manner, allowing for new insights provided by increased statistical power and refinement of prior signals. Here, we show that digital interventions tailored to an individual are effective in increasing short-term physical activity in a free-living cohort. These data suggest that participants are more likely to react positively and increase their physical activity when prompts are personalized.

**Funding:** Stanford Data Science Initiative and Catalyst Program, Apple, Google

## INTRODUCTION

All forms of physical activity are protective against the onset of chronic diseases ^1,2^. However, with the rapid increase in industrialization during the 20th century, humans have become sedentary, with an accompanying increase in the incidence and prevalence of cancer, cardiovascular disease, and dementia^3^. Modern Americans achieve a mean of 4700 daily steps, and the majority do not meet the modest recommendation of 150 minutes of exercise per week^4,5^.

Step count, as measured by a pedometer or more recently a smart device (phone and/or watch), has been studied as a proxy for physical activity^6^. Interventions that successfully increase step count are associated with reduced incidence of hypertension, reduced body-mass index, and improved cardiometabolic biomarkers^7^, highlighting the importance of step count as a metric of overall cardiovascular health.

The MyHeart Counts Cardiovascular Health Study was launched to leverage the rapid improvement in handheld technologies^8^, to better elucidate health patterns of participants and test digital interventions on short-term physical activity. Through the MyHeart Counts smartphone and watch app^9^, researchers are able to continuously enroll participants using automated protocols, benefit from advances in technological platforms, and reveal previously obscure patterns. Since its launch, the MyHeart Counts Cardiovascular Health Study has become an international, multi-arm, federated study with numerous sub-studies in the Netherlands, Canada, and the United Kingdom, with specific sub -study focuses on SARS-CoV2 infection, public health, and cardiovascular health. We first published on the feasibility of a fully digital study examining these varied outcomes using smartphones and smartwatches ^10^. Later, we released the data of 50,000 participants who agreed to broad data sharing with the larger scientific community^9^.

We recently reported findings from our first analysis of the randomized crossover trial subset of the MyHeart Counts Cardiovascular Health Study^11^. In this work, we determined that all smartphone-delivered digital interventions increased the mean daily step count ^11^. However, this first analysis did not have sufficient statistical power to demonstrate differences between interventions, and could not answer the question as to whether personalized or more frequent interventions were superior to more general prompts.

Given the unique design of our fully digital trial, we continued to have passive enrollment into the randomized crossover trial. This coupled with increased adoption of wearable devices allowed for the statistical power to detect differences between different types of digital interventions. Leveraging these unique advantages to digital trials, we present the latest results of an entirely digital randomized crossover trial using a subset of data from the MyHeart Counts Cardiovascular Health Study, and examine the effects of app-driven randomized interventions on step counts.

## METHODS

### Study design and participants

Ethical approval for the study was obtained from Stanford University’s Research Compliance Office.

The participants for this randomized controlled crossover trial were a substudy of the larger MyHeart Counts Cardiovascular Health Study^9–11^. All iPhone (Apple, Cupertino, CA, USA; version 5S or newer; iOS mobile operating system version 9 or newer) users in the USA, the UK, and Hong Kong, aged 18 years or older, who were able to read and understand English, and who had downloaded the latest version of the MyHeart Counts app (version ≥2.0) were eligible to participate in this crossover trial. A full description of the MyHeart Counts app has previously been reported, including a complete set of app screenshots ^11^.

After downloading the MyHeart Counts app from the Apple App Store, users were guided through an e-consent process^12^ (screenshots of this consent process previously published^11^ and available in Appendix pages 13-43). Participants had the option to either share their data narrowly (with Stanford University researchers only) or broadly (with qualified researchers worldwide). All participants had to make an active choice to complete the consent process, as no default choice was selected^12^.

### Randomization and masking

After completing the active consent process, users underwent a 1 -week period of baseline interaction with the MyHeart Counts app. After this 1-week period, participants were assigned to one of five clusters based on their level of physical activity during the weekdays and the weekend^10^. The five clusters consisted of: (1) individuals active throughout the baseline week, (2) individuals active on weekends but sedentary on weekdays, (3) individuals active on weekdays but sedentary on weekends, (4) individuals who were largely sedentary throughout the week, and (5) individuals who spent at least 15% of their awake time driving.

When opening the MyHeart Counts app for the first time after completion of the baseline week of monitoring, a pop-up notification with a second consent that required an active choice was presented: with the option to participate in a 4-week randomized crossover study of fully digital coaching (see Appendix pages 44-65). After enrollment, participants were randomly assigned to receive one of four interventions in a random order of 24 total permutations (4 combinations of 4 one-week digital interventions)^11^. The interventions consisted of: (1) daily instructions to read guidelines from the American Heart Association website, (2) daily prompts to complete 10,000 steps, (3) hourly prompts to stand for 1-minute after 1-hour of inactivity/sitting, and (4) daily e-coaching personalized to the individual’s personal activity patterns derived from the baseline week of data collection (see above). Thus, all participants were presented with all interventions, albeit at different times and in a different order.

Due to the nature of the study and interventions, it was not possible to blind participants to intervention assignment.

### Procedures

The HealthKit toolkit collected information on daily step count, distance walked, time spent in bed, and time spent asleep during the 1-week baseline interaction period of the MyHeart Counts app^13^. Motion sensors (either from a smartphone or smartwatch) were used to record time spent walking, running, cycling, resting, and driving each day. After completion of the 1-week baseline monitoring period and consenting to the e-coaching study (see above), participants were randomly assigned to each intervention for 7-days.

The four interventions were delivered serially to users as daily (or hourly, in the case of the stand reminders) prompts to their smartphones. Examples of the specific prompts have previously been published^11^. For the intervention to read the American Heart Association website, a daily prompt was sent with a link to the website. For the daily 10,000 step prompt intervention, if users had not completed 5000 steps by 1500h local time, they received a prompt indicating the number of steps remaining to reach the goal. If the user had completed more than 5000 steps by 1500h local time, no prompt was triggered. For the hourly stand intervention, if the user had been seated for the past 1h, they received a prompt advising them to stand for 60 seconds. If the user had been active in the past hour, they received no prompt. Finally, for the personalized e-coaching intervention, a daily message was sent to the participant, tailored to the activity cluster that they had been assigned to (see Appendix **eTable 1**).

Activity cluster identification has previously been described in detail ^10,11^. In brief, data collected by the 1-week baseline monitoring period was used for k-means clustering along 10-dimensions: time spent stationary, driving, walking, cycling, and running, stratified by weekday vs weekend days. Participants were then clustered into the closest activity group based on Euclidean distance in the feature space.

Amazon Web Services (Amazon, Seattle, WA, USA) was used to store information on user interactions with the four intervention prompts. Through confirmation with mobile analytics, it was ascertained whether or not the participants received intervention prompt messages on their smartphones daily (or at least one hourly stand reminder a day). While this analysis did not guarantee that a participant opened their notification, it did confirm whether or not a notification was displayed on the smart device, and not missed because of other reasons (e.g., cell phone switch off, privacy settings, and connectivity etc).

### Outcomes

Mean daily step count, as recorded by participants’ smartphones via HealthKit, was used as the primary outcome. A *post-hoc* sensitivity analysis was performed to determine if there were differences in intervention effects in a subset of individuals who (1) confirmed that they had received intervention prompt messages on their smartphones for the given intervention and (2) who had both smartphone and Apple Watch data.

### Statistical Analyses

Analyses were performed in a modified intention-to-treat population ^14^, which included all participants that completed the 1-week period of baseline monitoring and at least 1 day of one of the four interventions. Prior exploratory analyses demonstrated no differences between using data from participants with 1-, 4-, and 7-days of at least 200 steps.^11^ Hence, we continued to use the least stringent inclusion criteria for our analyses. Daily step count for each participant was calculated by summing values for the HKQuantityTypeIdentifierStepCount field that flowed from HealthKit into the MyHeart Counts app. Days during which the participant did not reach 200 steps as measured by HealthKit on their smartphone were excluded from analyses. Users were considered to have been provided the intervention on days in which they were active and exceeded step count (for the daily step count intervention) or inactivity (for the hourly stand intervention) to avoid triggering reminder prompts.

All analyses were performed in R^15,16^. Tukey’s Honestly Significant Difference (HSD) test was used with a significance level of 5% to assess for statistically significant differences in mean daily step count between each pair of interventions and baseline ^17^. In addition, a false discovery rate-adjusted *P*-value threshold of 0.05 was used to determine whether differences between two interventions was statistically significant.

### Data Availability

The data that support the findings of this study are available on request from the corresponding authors, DSK and EAA. The data are not publicly available due to the Health Insurance Portability and Accountability Act of 1996 (HIPAA).

## RESULTS

Between January 1, 2017, and April 1, 2022, 4500 participants consented to participate in the e - coaching study (**Figure 1**). Of these 4500 participants, 2458 completed the one-week period of baseline monitoring and at least one day of one of four interventions and were included in the modified intention-to-treat analyses (**Table 1**). These 2458 participants were majority men (68.6%) and self -identified white (82.3%) with a mean age of 46.1 years (SD 15.5). The distribution of participants body mass index (BMI) categories was 4.9% underweight (BMI<18.5), 34.4% normal weight (BMI 18.5 to <25), 35.0% overweight (BMI 25 to <30), 15.3% obese (BMI 30 to <35), and 10.4% severely obese (BMI 35 to <40).

**Figure 1.**
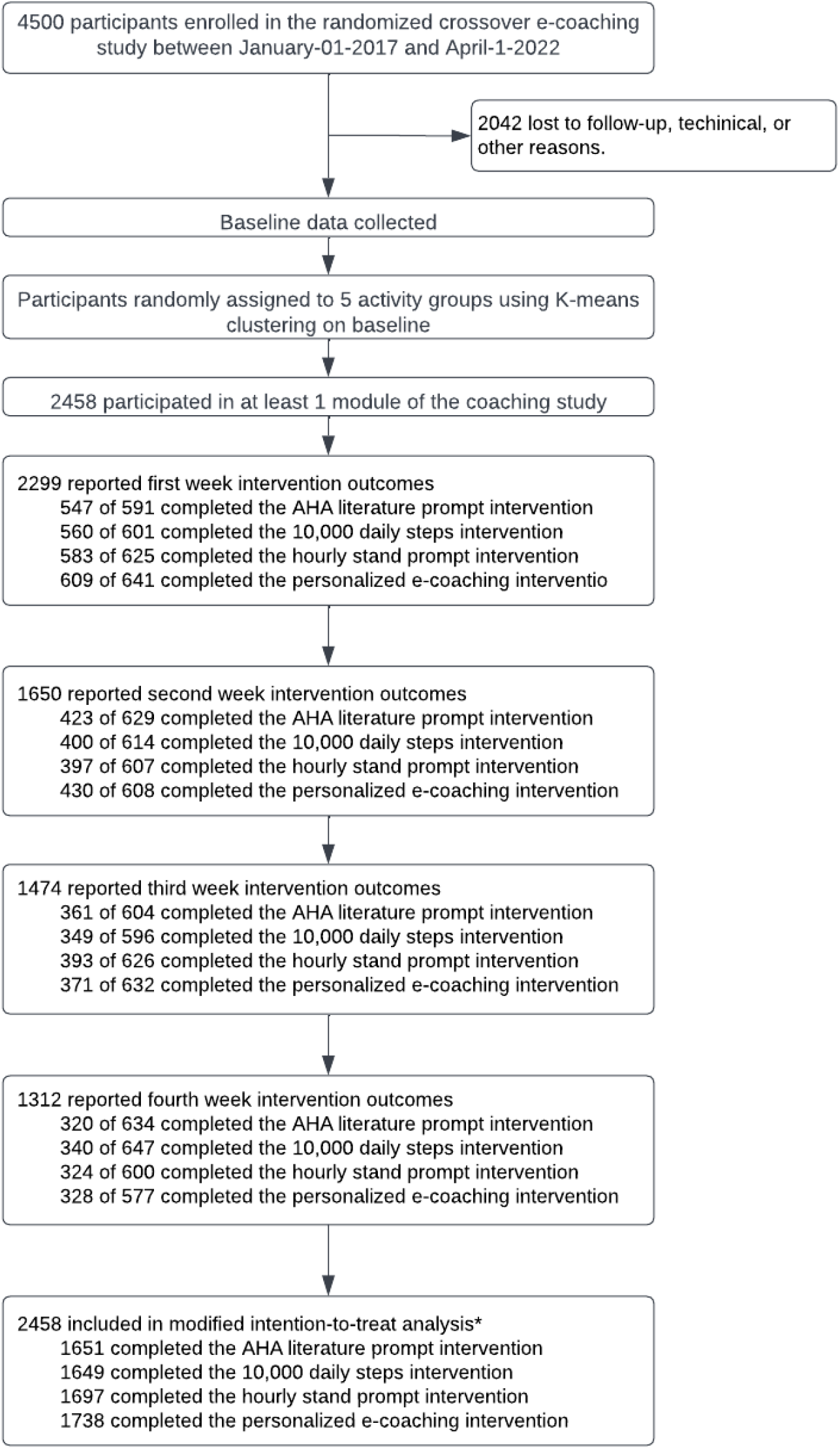
Flow of participants through the study.

**Table 1.** Baseline self-reported characteristics of the studied subset of the MyHeart Counts Cardiovascular Health Study included in modified intention-to-treat analyses.

|  |  |  | Participants who completed baseline and at least one intervention (n=2458)* |
| --- | --- | --- | --- |
| Age, years |  | N = 2216 | 46.1 (15.5) |
| Self-Identified Biological Sex, n (%) |  |  |  |
|  | Male |  | 1544 (68.6%) |
|  | Female |  | 707 (31.4%) |
|  | Unknown |  | 207 |
| Self-Identified Race/Ethnicity, n (%) |  | N = 2017 |  |
|  | White |  | 1660 (82.3%) |
|  | East/Southeast Asian |  | 101 (5.0%) |
|  | Black |  | 62 (3.1%) |
|  | South Asian |  | 44 (2.2%) |
|  | Hispanic |  | 18 (0.89%) |
|  | American Indian |  | 6 (0.3%) |
|  | Multi-Racial |  | 55 (2.7%) |
|  | Other |  | 71 (3.5%) |
|  | Unknown |  | 441 |
| Body mass index (BMI) category, n (%) |  | N = 2209 |  |
|  | Underweight (BMI<18.5) |  | 110 (4.9%) |
|  | Normal Weight (BMI 18.5 to <25) |  | 760 (34.4%) |
|  | Overweight (BMI 25 to <30) |  | 773 (35.0%) |
|  | Obese (BMI 30 to <35) |  | 337 (15.3%) |
|  | Obese II (BMI 35 to <40) |  | 229 (10.4%) |
|  | Unknown |  | 249 |
| Height, inches |  | N = 2229 | 68.5 (3.9) |
| Weight, pounds |  | N = 2210 | 183.9 (45.7) |
| Device use, n (%) |  | N = 2458 |  |
|  | Smartphone + Smartwatch |  | 1823 (74.2) |
|  | Smartphone only |  | 393 (16.0) |
|  | Smartwatch only |  | 194 (7.9) |
|  | Unknown |  | 48 (1.9) |

\*Unknown device use due to interpretation of device naming. For example, "John's iPhone" would be classified as a smartphone, while "John's Apple Watch" would be classified as a smartwatch. However, a device named "John" would be classified as "unknown".

During the 1-week baseline monitoring period, the 2458 participants walked an average of 4233 steps daily (SE 73, **Table 2**). Relative to this baseline monitoring period, all interventions with the exception of the 10,000 step prompt intervention significantly increased mean daily step count (**Table 2, Figure 2**). The number of steps recorded by smartphone increased from baseline by 215 (SE 71) for participants in the read American Heart Association website prompt group (*P*=0.021), by 170 steps (SE 71) for participants in the 10,000 step daily prompt group (*P*=0.11), by 292 steps (SE 70) for participants in the hourly stand prompt group (*P*=0.00029), and by 402 steps (SE 70) for participants in the personalized e-coaching prompt group (*P=*7.1×10^−8^).

**Table 2.**
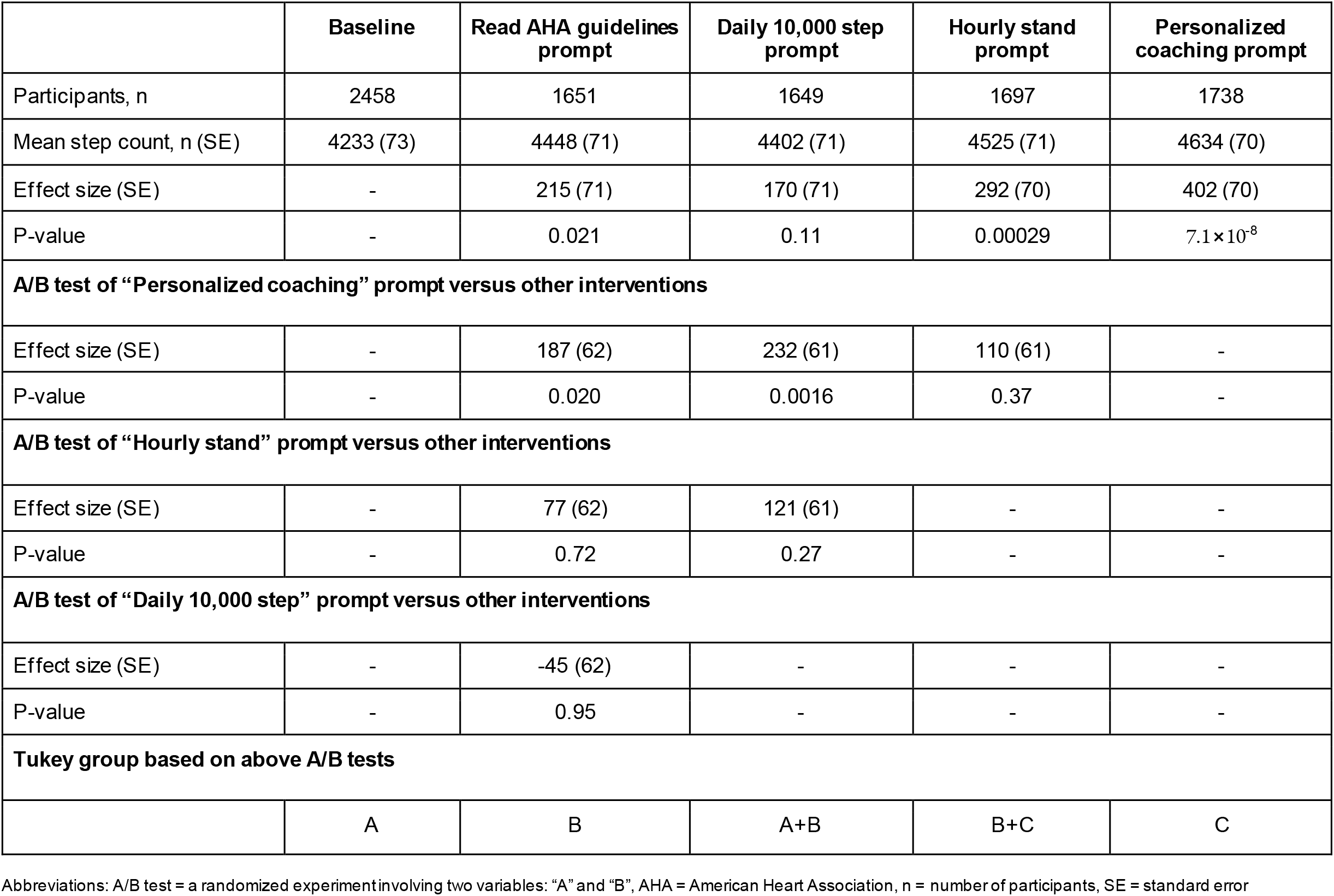
Intervention effects on mean daily step count in the modified intention-to-treat subset of the MyHeart Counts randomized crossover trial.

**Figure 2.**
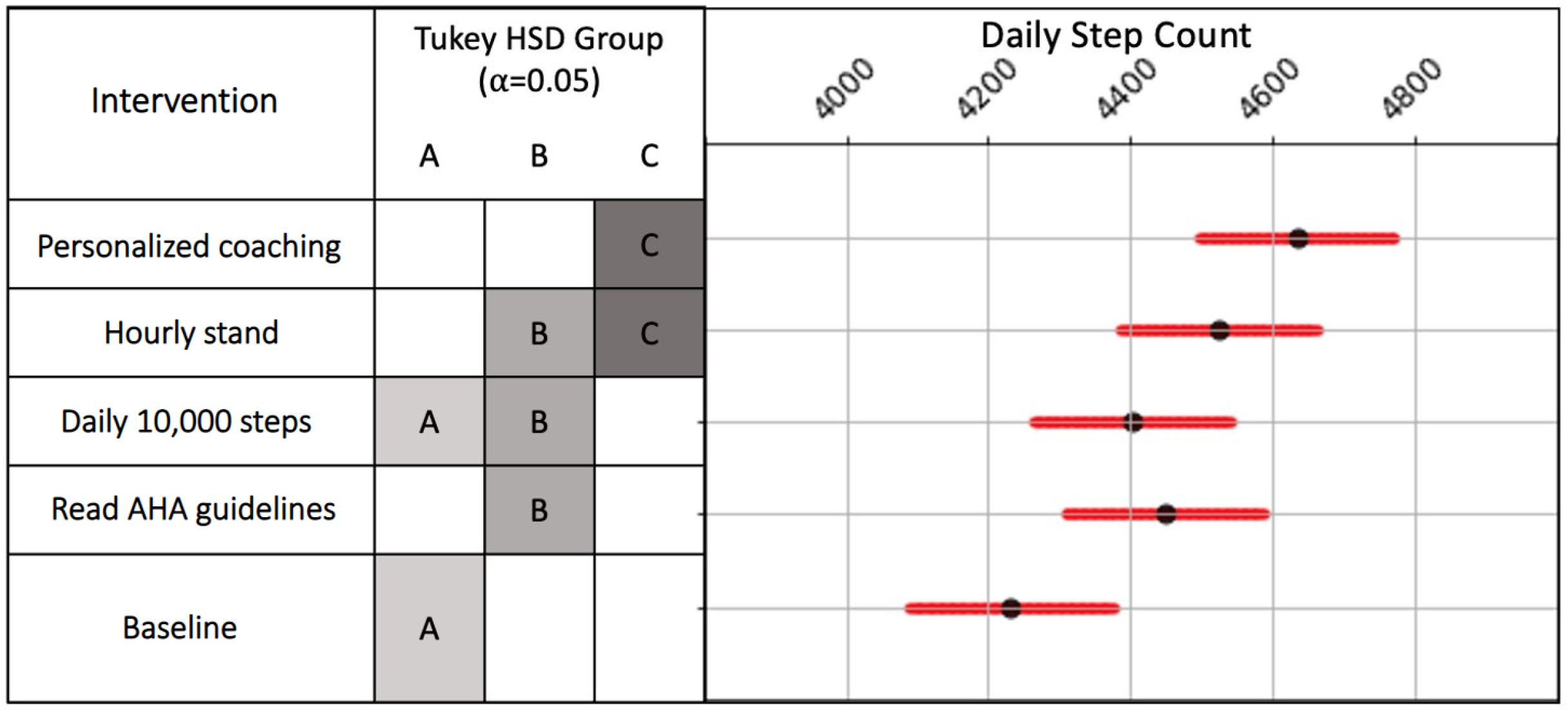
Mean daily step count for the 1-week baseline period and for the four interventions in the modified intention-to-treat subset of the randomized crossover MyHeart Counts Study. Mean daily step count was recorded by the MyHeart Counts iPhone app. Error bars show 95% CI. Interventions assigned to different Tukey groups are significantly different from each other.

When limiting analyses to a subset of participants for whom mobile analytics confirmed that the device received the intervention prompt (N=2452), similar results were obtained (Appendix **eTable 2**). As with the primary analyses, the personalized e-coaching group had the greatest increase in mean daily step count at 531 steps (SE 70). As well, the same groups were assigned to interventions when using Tukey HSD analysis (Appendix **eTable 2**).

A sensitivity analysis was performed to ensure that results were similar in a subset of participants who had both smartphone and smartwatch data (N=1823). The same Tukey HSD groups were obtained for the different digital interventions (Appendix **eTable 3)**. As with the primary analysis, the personalized e-coaching intervention resulted in the greatest increase in mean daily steps from baseline with 411 steps (SE 75) (Appendix **eTable 3**).

When comparing the present analysis (N=2458) to our previously reported work ^11^ (N=1075), there is refinement in both effect and error estimation (**Figure 3**). With a smaller sample size, we previously reported that no digital intervention was significantly different than another. However, with the addition of 1383 participants, effect estimation was improved and the personalized coaching intervention was the only intervention to increase its mean daily step count estimate compared with the prior analysis.

**Figure 3.**
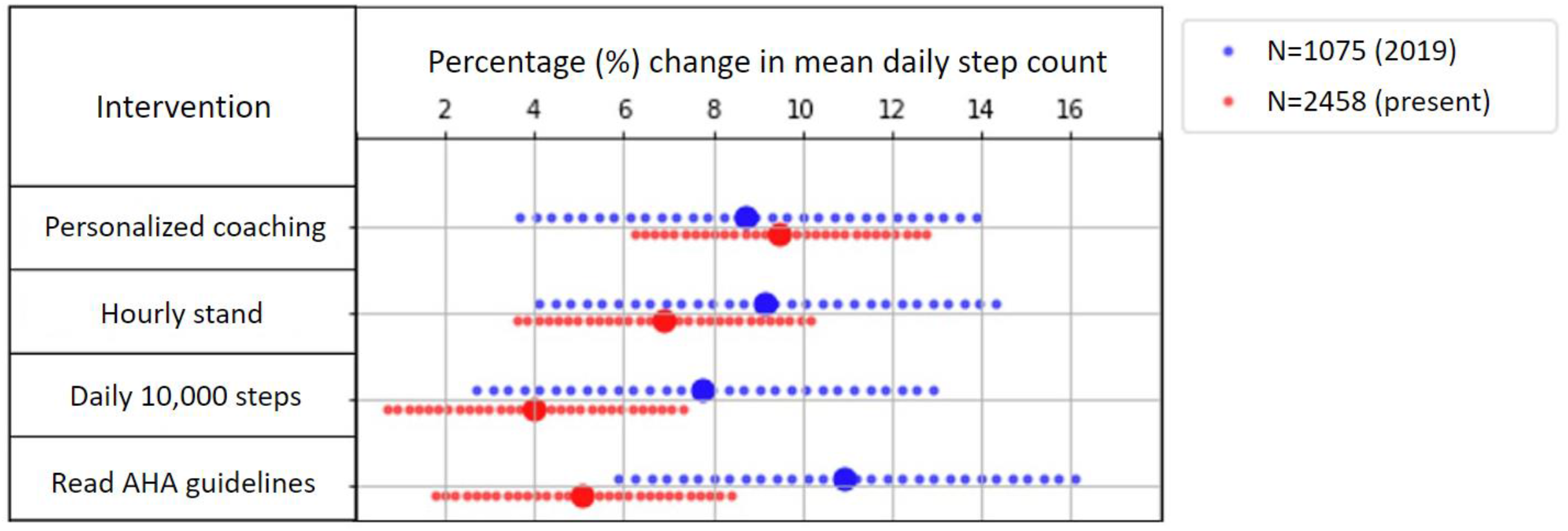
Comparison of intervention effects in present data (N=2458) as compared to previously published data (N=1075) in the modified intention-to-treat subset of the MyHeart Counts randomized crossover trial. Error bars show 95% CI.

## DISCUSSION

This follow-up analysis of a randomized crossover trial demonstrates the power of digital studies to continually enroll participants through a smartphone app and complete interventions in a fully automated manner. Leveraging the increased statistical power provided by further passive enrollment, we determined that personalized e-coaching resulted in the greatest increase in mean daily step count (9.5% increase; our prior work was unable to differentiate between the effects of the varied interventions ^11^). These data highlight the potential of digital studies to extend the reach of clinical trials in a cost-effective manner ^18^. Moreover, our results suggest that tailoring interventions to the individual is superior to a “one-size-fits-all” approach^19–21^.

Digital trials have the potential to greatly improve future research given that they are cheaper and less labor intensive than traditional clinical trials^22,23^. Moreover, as the only requirement to participate in such trials is a smartphone, digital trials have the potential to increase equity by including people around the world, rather than focusing on those only in first -world countries. Particularly during the SARS-CoV2 pandemic, digital trials had a significant advantage over traditional trials, due to continued cost-effective enrollment without requiring face-to-face interactions on the investigators’ end^24^. Specific to MyHeart Counts, our present work was made possible only through the unique and fully digital design of our randomized crossover trial ^9–11^. By then leveraging this continued passive enrollment into the MyHeart Counts coaching trial and the resultant increased statistical power, we were able to further refine prior signals and identify that personalized e-coaching interventions were superior in increasing short-term physical activity (**Figure 3**). Our results can be extrapolated to other ongoing digital trials, and demonstrate another significant advantage of fully mobile/automated studies as compared to traditional trials.

Our present data suggests that personalized e-coaching prompts, specific to an individual’s baseline activity level, is the most effective intervention in increasing their short -term physical activity. Although “precision medicine” has become synonymous with tailoring therapies to an individual’s genetic background^19–21^, the broader principle reflects integrating all available data to make the most informed clinical decision for each individual patient ^20^. Within this context, our findings highlight the importance of tailoring interventions (whether genetic, medication, or digital) to each individual to achieve maximum efficacy.

Interestingly, we found that daily prompts to complete 10,000 steps did not significantly increase the mean daily step count from baseline, in contrast to our prior report from 1075 participants ^11^. This may be due to the timing of this prompt, which triggered at 1500h local time if the participant had not yet reached 5000 steps, leading to a prompt that was too late to elicit significant behavioral change for the day. In contrast, hourly reminders to stand after inactivity were nearly as effective as personalized e-coaching in our study, possibly due to increased prompting (hourly versus daily). These data suggest that, in addition to personalizing interventions to the individual, more frequent interventions and those earlier in the day are more effective in increasing short-term physical activity.

Other strengths of our study include the increase in smartwatch use in the general population with time, which resulted in a significantly higher proportion of participants with smartwatch data in our current analyses as compared to our prior work (82.1% vs 24.7%). In our present data and in prior analyses^11^, we found that those with smartwatches had higher mean daily step count as compared to smartphone-only users, likely due to smartwatches being worn for all daily activities. As smartwatches are more accurate than phones in tracking step count ^25^, this allows for greater sensitivity to detect changes with each intervention.

Another strength of our study is internal validity demonstrated by our sensitivity analyses. In separate *post hoc* analyses of those who confirmed received notifications and those with smartwatches, consistent results with personalized e-coaching yielding the greatest increase in mean daily step count were obtained. We previously demonstrated that there were no significant differences in attrition of participants across the four interventions and separately, that there were no demographic differences in participants who dropped out as compared to those who were included in the modified intention-to-treat analyses^11^.

Several limitations of our study should be considered. As with other digital trials ^26,27^, our study had significantly decreased user retention as compared with traditional trials. Other studies have demonstrated that gamification, such as badges earned for completing activities, improves retention^28^ and this was previously implemented into the MyHeart Counts study ^11^. A separate meta-analysis suggested that useful feedback and financial compensation were effective in increasing retention rates in digital health trials^29^. These possibilities will be explored in further extensions of the MyHeart Counts Cardiovascular Health Study; however, with data available, the high attrition rate combined with the average user being a middle-aged white male, can limit generalizability of our findings. Finally, the long-term effects of e-coaching on changes to physical activity are yet unknown^30^. Studies are ongoing to determine the effects of digital prompts on physical activity with long-term follow-up^22^.

In summary, our extended randomized crossover trial demonstrates the strength and cost-effectiveness of fully digital and automated trials in continuing enrollment after primary analyses have concluded. Through this increased statistical power, we were able to identify that personalized e-coaching interventions were most effective in increasing short-term physical activity, with results that were consistent across sensitivity analyses. These data highlight the promise of digital trials and emphasize the importance of tailoring intervention s to the individual, rather than applying a single therapy to all patients.

## Supporting information

Appendix of all Supplemental Materials

## AUTHOR CONTRIBUTIONS

MVM, ACK, and EAA conceptualized and designed the study. SGH managed app development. ACK and EAA designed the interventions. AJ and DSK did statistical analysis. AJ, DSK, AJ, AT, JWO, MTW, and EAA contributed to acquisition, analysis, or interpretation of the data. DSK and EAA drafted the manuscript. AJ, SGH, AS, JWO, MVM, LL, ACK, JWC, MO, CMM, RAA, MTW, and EAA critically revised the manuscript for important intellectual content. MTW and EAA supervised the study.

