## Appendix of all Supplemental Materials for "Personalized digital behavior interventions increase short term physical activity: a randomized control crossover trial substudy of the MyHeart Counts Cardiovascular Health study"

Affiliations:
1. Division of Cardiovascular Medicine, Department of Medicine, Stanford University School of Medicine, Stanford, CA, USA
2. Biofourmis, Boston, MA, USA
3. identifEYE HEALTH, Redwood City, CA, USA
4. Department of Health Research and Policy, Stanford University, Stanford, CA, USA
5. Stanford Prevention Research Center, Department of Medicine, Stanford University School of Medicine, Stanford, CA, USA
6. Stanford Cardiovascular Institute, Stanford University, Stanford, CA, USA
7. Department of Genetics, Stanford University School of Medicine, Stanford, CA, USA
8. Department of Biomedical Data Science, Stanford University School of Medicine, Stanford, CA, USA

**Statistical analysis plan** 3

**Supplemental e-tables** 4

**CONSORT checklist** 12

**Screenshots of consent process** 13

**Screenshots of MyHeart Counts app interventions** 44

**IRB trial protocol** 66

#### STATISTICAL ANALYSIS PLAN

**Sample size calculation:**

Power analysis was performed to determine the number of subjects needed to identify an effect size of 2000 steps with p-value < 0.05. The 2000 step effect size was deemed feasible based on the findings of prior randomized controlled trials of interventions aimed at increasing physical activity levels(Tudor-Locke 2011). From the initial phase of the MyHeart Counts study(McConnell 2016), the mean and standard deviation of daily step count from HealthKit was determined to be 3200+/-200 steps. Using these heuristics, a one-tailed T-test power calculation was performed, suggesting that an RCT participant sample size of N= 1000 had power=1 to detect an increase of 2000 steps with p- value=0.05. The same approach suggested power=0.99 to detect an increase of 1000 steps, power = 0.94 to detect an increase of 500 steps, and power=0.37 to detect an increase of 200 steps. This analysis resulted in a pre-specified sample size of N=1000 for the previously published analysis of the randomized control trial subset of the MyHeart Counts study(Shcherbina 2019). Since publication of this trial, the MyHeart Counts study has continued to enroll patients via its app on the Apple App Store, with subsequent passive enrollment into the cluster randomized trial.

**Ascertainment of Dropout:**

Data from study participants were included for analysis if they completed one or more days of baseline monitoring and one or more days in at least one of the four interventions. To meet these criteria, a participant must have registered 200 or more steps via HealthKit on either their smartphone or smartwatch. Any days when a participant did not register 200 or more steps on their phone or watch were excluded from analysis.

**Determining Significance of Intervention Effects:**

The effects of the four interventions on the primary outcome of HealthKit daily step count were assessed via multivariate regression modeling. An FDR-corrected p-value threshold of 0.05 was used to determine whether or not differences between two interventions were significant.

The modeling was done with the R “nlme” library. Subjects were treated as random effects. Number of days in study and the intervention groups were treated as fixed linear effects. Interaction terms between days in study and intervention were evaluated for significance in the model. Quadratic terms for days-in-study were evaluated for significance in the model.

Data were analyzed via an intention-to-treat paradigm. It is possible that subjects did not receive an intervention on a given day -- i.e. if they did not remain sedentary for one hour straight, they would not receive the hourly stand prompt; if they completed the daily 10,000 step goal, they would not receive the daily walk prompt. However, those subjects will still be included for analysis in the “Hourly Stand” and “Daily Step Count” intervention groups due to the intention-to-treat design.

For the primary outcome, step counts of less than 200 steps per day or greater than 50,000 steps per day were excluded from analysis.

### SUPPLEMENTAL E-TABLES

**eTable 1. Digital interventions given to different baseline activity clusters for those randomized to the "personalized e-coaching" cluster.**

| | |
|---|---|
| **Busy Bee Cluster** | **Introduction: Congratulations! Your typical physical activity patterns place you in the top group of MyHeart Counts users! You take the appropriate steps to protect your heart health, and we applaud you.**<br><br>● Stay active! App users who stay active throughout the week report, on average, higher overall satisfaction with life than those in less active groups.<br>● Logging all those steps is working! App users in your activity group report, on average, 10 points lower blood glucose levels than those in less active groups.<br>● App users in your group report a significantly lower prevalence of heart disease than the general population.<br>● App users in your activity group report significantly lower incidence of blood vessel disease than the general population. You are definitely making your heart count!<br>● Users in your activity group report lower blood pressure than all other groups. You're doing things right!<br>● Participants as active as you are report feeling less worried than those who are inactive.<br>● App users in your activity group report feeling happier with life than those who are sedentary. |

| | |
|---|---|
| **Sedentary Cluster** | **Introduction: According to your activity profile, you fall in the segment of our app users who need to improve and increase their activity levels. You signed up and made your heart count - now make a conscious effort to increase your activity each day.**<br><br>● Did you know that MyHeart Counts participants who are more physically active throughout the week reported being more satisfied with life than participants in your activity group?<br><br>● MHC users who get active on the weekend report higher life satisfaction than those who are not active on the weekend. Adding just 1 or 2 days of activity makes a difference in how you feel. So, how about a walk this weekend?<br><br>● More active people using this app report feeling more \"worthwhile\" than those who don't get much activity<br><br>● Our most physically active users report feeling less depressed and less worried than less active users. Try moving more regularly and watch your mood brighten!<br><br>● App users who mainly get active on the weekends report on average 12 points lower LDL cholesterol than non-active individuals. Every step helps make a positive impact! How about that Sunday morning walk?<br><br>● Users who don't get much physical activity are more likely to report joint problems than their more active counterparts. Our bodies are made to move – your joints will thank you!<br><br>● App users getting less physical activity are significantly more likely to report heart disease than their more active counterparts. Try adding some steps to your day today. |

<table>
<tr>
<td>Driver Cluster</td>
<td>Introduction: So, we noticed you spend a lot of time in the car, compared to other app users. We know you have to get from point A to point B, and that requires sedentary time in the car. There's no better way to offset the sedentary effects than to incorporate physical activity into your day.<br><br>
● So you drive a lot. Did you know MyHeart Counts users who drive a lot but are also physically active report feeling more satisfied with life than those who report minimal activity?<br><br>
● While users in your activity group report feeling less happy than their more active counterparts, those who drive a lot yet are also physically active report feeling happier than less active individuals.<br><br>
● Participants who drive a lot but still engage in regular physical activity report feeling less depressed than those who get minimal to no activity.<br><br>
● People in your group, who drive a lot but are also very active, report feeling less worried than those who lead more sedentary lives.<br><br>
● Participants in your group report, on average, 10 points higher blood glucose than their more active counterparts. That's all the more reason to walk to your next destination or fit in walking in other ways throughout the day.<br><br>
● Did you know that participants in your activity group report an increased prevalence of heart disease, compared to the general population? Adding more physical activity to your day is a key way to help reduce your risk.<br><br>
● People in your activity group report a higher incidence of vascular disease than the general population. That's one more good reason to walk more today!</td>
</tr>
</table>

<table>
<tr>
<td>Worker Bees Cluster</td>
<td>Introduction: Congratulations! Your activity level puts you in one of our more physically active MyHeart Counts participant groups. Clearly, you know what it takes to make your heart count, so please keep up the good work!
<ul>
<li>You get your activity in during the week – great job! You also tend to be more sedentary during the weekend – what’s up with that? Individuals who are active throughout the whole week report higher overall life satisfaction than those who are only active during the workweek. Try logging some steps this weekend, and see if it affects how you feel.</li>
<li>Being more active throughout the entire week will help lower your risk of heart disease even more.</li>
<li>MyHeart Counts users who get active on the weekends report feeling less worried than those who spent their weekends not being active.</li>
<li>Users who are active throughout the week, including weekends, report feeling less depressed than those who don’t do much activity on weekends.</li>
<li>Keep up the good work! Individuals who are active during the workweek report feeling happier than those who remain sedentary.</li>
<li>Try being active on weekends as well as weekdays. MyHeart Counts users who are active daily report feeling more worthwhile and satisfied with life than those who do not log much activity. Make every day count!</li>
<li>MyHeart Counts users who get their activity daily report feeling more satisfied with life than users who are not active each day.</li>
</ul>
</td>
</tr>
</table>

<table>
<tr><td>Weekend Warrior Cluster</td><td><ul>
<li>Introduction: Looks like you like to get outside and get active on weekends! Your activity levels go up towards the end of the week, and you show more physical activity during that time than our other MyHeart Counts users. Great work on getting out and enjoying your free time! We know you are busy during the week, but every little bit counts so consider increasing your activity on Monday to Friday as well.</li>
<li>Did you know that your activity patterns place you in the MyHeart Counts Weekend Warriors group? Overall, this group gets much of their activity on weekends and reports a lower incidence of heart conditions than the general population. Does this surprise you?</li>
<li>App users in your activity group report, on average, 3 points lower blood pressure than other activity groups. Good work, but there's definitely room to improve. How about adding some activity during the work week?</li>
<li>App users in your group report that they are less worried than more sedentary individuals and those who are only active during the workweek.</li>
<li>MyHeart Counts users in your group report feeling less depressed than participants who engage in very little activity.</li>
<li>Weekend Warriors report feeling more satisfied with life than inactive participants.</li>
<li>Weekend Warriors report feeling more \"worthwhile\" than sedentary participants.</li>
<li>Weekend Warriors report feeling happier than participants who are inactive.</li>
</ul></td></tr>
</table>

**eTable 2. Intervention effects on mean daily step count in a subset of individuals who confirmed that they received the digital intervention during the randomized crossover trial.**

| | **Baseline** | **Read AHA guidelines prompt** | **Daily 10,000 step prompt** | **Hourly stand prompt** | **Personalized coaching prompt** |
|---|---|---|---|---|---|
| Participants, n | 2452 | 845 | 582 | 765 | 957 |
| Mean step count, n (SE) | 4211 (75) | 4549 (96) | 4371 (119) | 4725 (100) | 4742 (92) |
| Effect size (SE) | - | 338 (105) | 161 (127) | 515 (109) | 531 (70) |
| P-value | - | 0.012 | 0.71 | $2.3\times10^{-5}$ | $1.1\times10^{-6}$ |
| **AB test of "Personalized Coaching Prompt" versus other interventions** | | | | | |
| Effect size (SE) | - | 193 (90) | 371 (115) | 17 (98) | - |
| P-value | - | 0.20 | 0.011 | 0.99 | - |
| **AB test of "Hourly stand prompt" versus other interventions** | | | | | |
| Effect size (SE) | - | 176 (100) | 354 (122) | - | - |
| P-value | - | 0.39 | 0.029 | - | - |
| **AB test of "Daily 10,000 step prompt" versus other interventions** | | | | | |
| Effect size (SE) | - | -178 (118) | - | - | - |
| P-value | - | 0.55 | - | - | - |
| **Tukey group based on above AB tests** | | | | | |
| | A | B | A+B | B+C | C |

Abbreviations: A/B test = a randomized experiment involving two variables: "A" and "B", AHA = American Heart Association, n = number of participants, SE = standard error

**eTable 3. Intervention effects on mean daily step count in a subset of individuals who had both smartphone and smartwatch data while enrolled in the randomized crossover trial.**

| | Baseline | Read AHA guidelines prompt | Daily 10,000 step prompt | Hourly stand prompt | Personalized coaching prompt |
|---|---|---|---|---|---|
| Participants, n | 1823 | 1305 | 1314 | 1364 | 1377 |
| Mean step count, n (SE) | 4122 (85) | 4392 (81) | 4292 (81) | 4446 (81) | 4533 (81) |
| Effect size (SE) | - | 270 (76) | 170 (76) | 324 (75) | 411 (75) |
| P-value | - | 0.0033 | 0.16 | 0.00015 | $2.45 \times 10^{-7}$ |
| **A/B test of "Personalized Coaching Prompt" versus other interventions** | | | | | |
| Effect size (SE) | - | 141 (64) | 241 (64) | 87 (64) | - |
| P-value | - | 0.18 | 0.0017 | 0.65 | - |
| **A/B test of "Hourly stand prompt" versus other interventions** | | | | | |
| Effect size (SE) | - | 53 (64) | 154 (64) | - | - |
| P-value | - | 0.92 | 0.12 | - | - |
| **A/B test of "Daily 10,000 step prompt" versus other interventions** | | | | | |
| Effect size (SE) | - | -100 (65) | - | - | - |
| P-value | - | 0.54 | - | - | - |
| **Tukey group based on above A/B tests** | | | | | |
| | A | B | A+B | B+C | C |

### CONSORT CHECKLIST

#### CONSORT 2010 checklist of information to include when reporting a randomised trial*

| Section/Topic | Item No | Checklist item | Reported on page No |
|---|---|---|---|
| **Title and abstract** | | | |
| | 1a | Identification as a randomised trial in the title | 1 |
| | 1b | Structured summary of trial design, methods, results, and conclusions (for specific guidance see CONSORT for abstracts) | 2 |
| **Introduction** | | | |
| Background and objectives | 2a | Scientific background and explanation of rationale | 4 |
| | 2b | Specific objectives or hypotheses | 4 |
| **Methods** | | | |
| Trial design | 3a | Description of trial design (such as parallel, factorial) including allocation ratio | 5-7 |
| | 3b | Important changes to methods after trial commencement (such as eligibility criteria), with reasons | n/a |
| Participants | 4a | Eligibility criteria for participants | 5 |
| | 4b | Settings and locations where the data were collected | 5 |
| Interventions | 5 | The interventions for each group with sufficient details to allow replication, including how and when they were actually administered | 5-7 |
| Outcomes | 6a | Completely defined pre-specified primary and secondary outcome measures, including how and when they were assessed | 6 |
| | 6b | Any changes to trial outcomes after the trial commenced, with reasons | n/a |
| Sample size | 7a | How sample size was determined | Appendix p3 |
| | 7b | When applicable, explanation of any interim analyses and stopping guidelines | n/a |
| Randomisation: | | | |
| Sequence generation | 8a | Method used to generate the random allocation sequence | 5 |
| | 8b | Type of randomisation; details of any restriction (such as blocking and block size) | 5 |
| Allocation concealment mechanism | 9 | Mechanism used to implement the random allocation sequence (such as sequentially numbered containers), describing any steps taken to conceal the sequence until interventions were assigned | 5 |
| Implementation | 10 | Who generated the random allocation sequence, who enrolled participants, and who assigned participants to interventions | 5 |
| Blinding | 11a | If done, who was blinded after assignment to interventions (for example, participants, care providers, those | 5 |
| | | assessing outcomes) and how | 5 |
| | 11b | If relevant, description of the similarity of interventions | 5-6 |
| Statistical methods | 12a | Statistical methods used to compare groups for primary and secondary outcomes | 7 |
| | 12b | Methods for additional analyses, such as subgroup analyses and adjusted analyses | 7 |
| **Results** | | | |
| Participant flow (a diagram is strongly recommended) | 13a | For each group, the numbers of participants who were randomly assigned, received intended treatment, and were analysed for the primary outcome | 15 |
| | 13b | For each group, losses and exclusions after randomisation, together with reasons | 15 |
| Recruitment | 14a | Dates defining the periods of recruitment and follow-up | 5, 15 |
| | 14b | Why the trial ended or was stopped | n/a |
| Baseline data | 15 | A table showing baseline demographic and clinical characteristics for each group | 14 |
| Numbers analysed | 16 | For each group, number of participants (denominator) included in each analysis and whether the analysis was by original assigned groups | 17 |
| Outcomes and estimation | 17a | For each primary and secondary outcome, results for each group, and the estimated effect size and its precision (such as 95% confidence interval) | 17 |
| | 17b | For binary outcomes, presentation of both absolute and relative effect sizes is recommended | 16-18 |
| Ancillary analyses | 18 | Results of any other analyses performed, including subgroup analyses and adjusted analyses, distinguishing pre-specified from exploratory | Appendix p10-11 |
| Harms | 19 | All important harms or unintended effects in each group (for specific guidance see CONSORT for harms) | n/a |
| **Discussion** | | | |
| Limitations | 20 | Trial limitations, addressing sources of potential bias, imprecision, and, if relevant, multiplicity of analyses | 10 |
| Generalisability | 21 | Generalisability (external validity, applicability) of the trial findings | 10 |
| Interpretation | 22 | Interpretation consistent with results, balancing benefits and harms, and considering other relevant evidence | 9-10 |
| **Other information** | | | |
| Registration | 23 | Registration number and name of trial registry | 2 |
| Protocol | 24 | Where the full trial protocol can be accessed, if available | Appendix p66-100 |
| Funding | 25 | Sources of funding and other support (such as supply of drugs), role of funders | 2, 7, 11 |

### SCREENSHOTS OF CONSENT PROCESS TO THE GENERAL STUDY

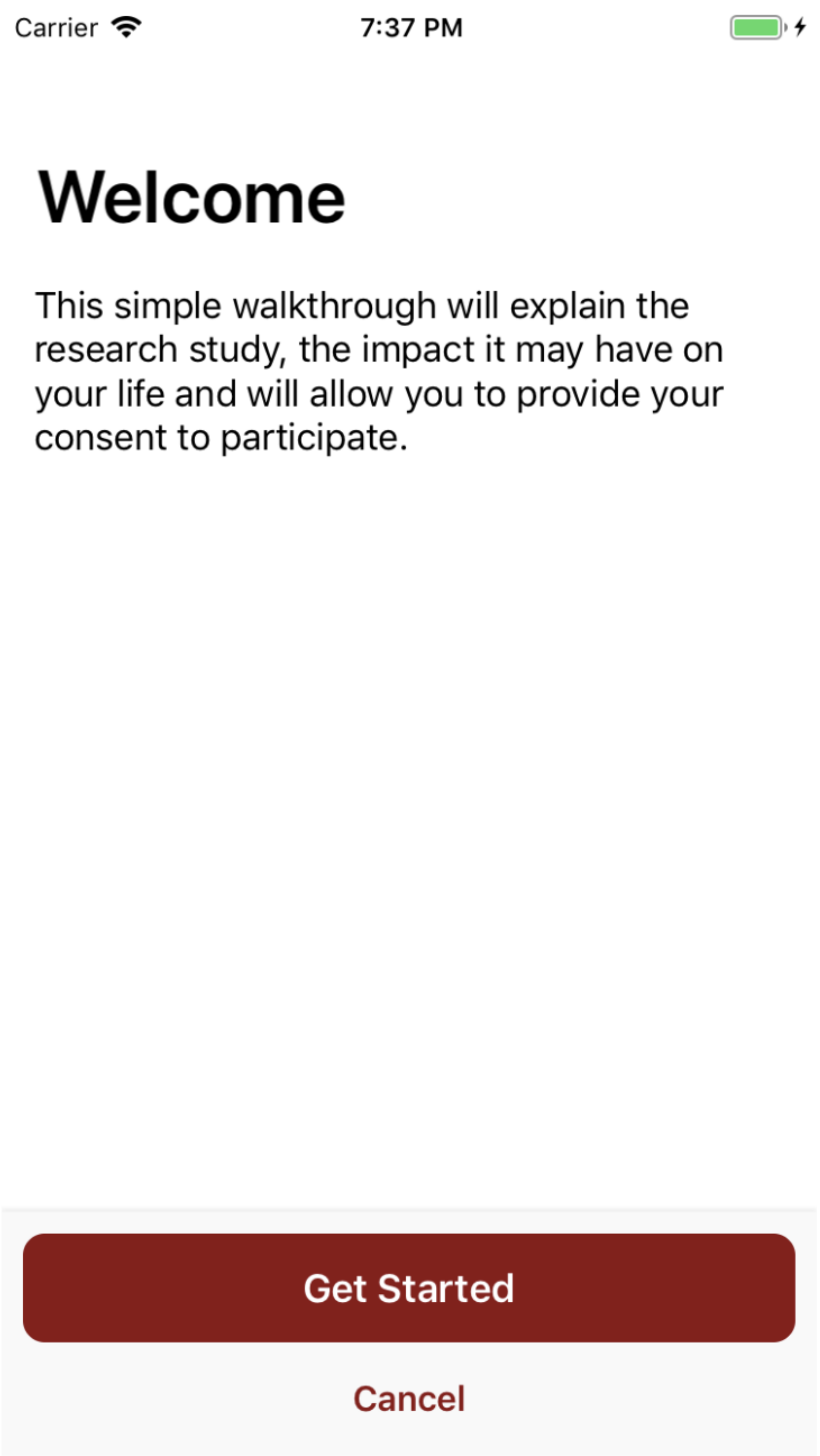
Carrier
7:37 PM
Welcome
This simple walkthrough will explain the research study, the impact it may have on your life and will allow you to provide your consent to participate.
Get Started
Cancel

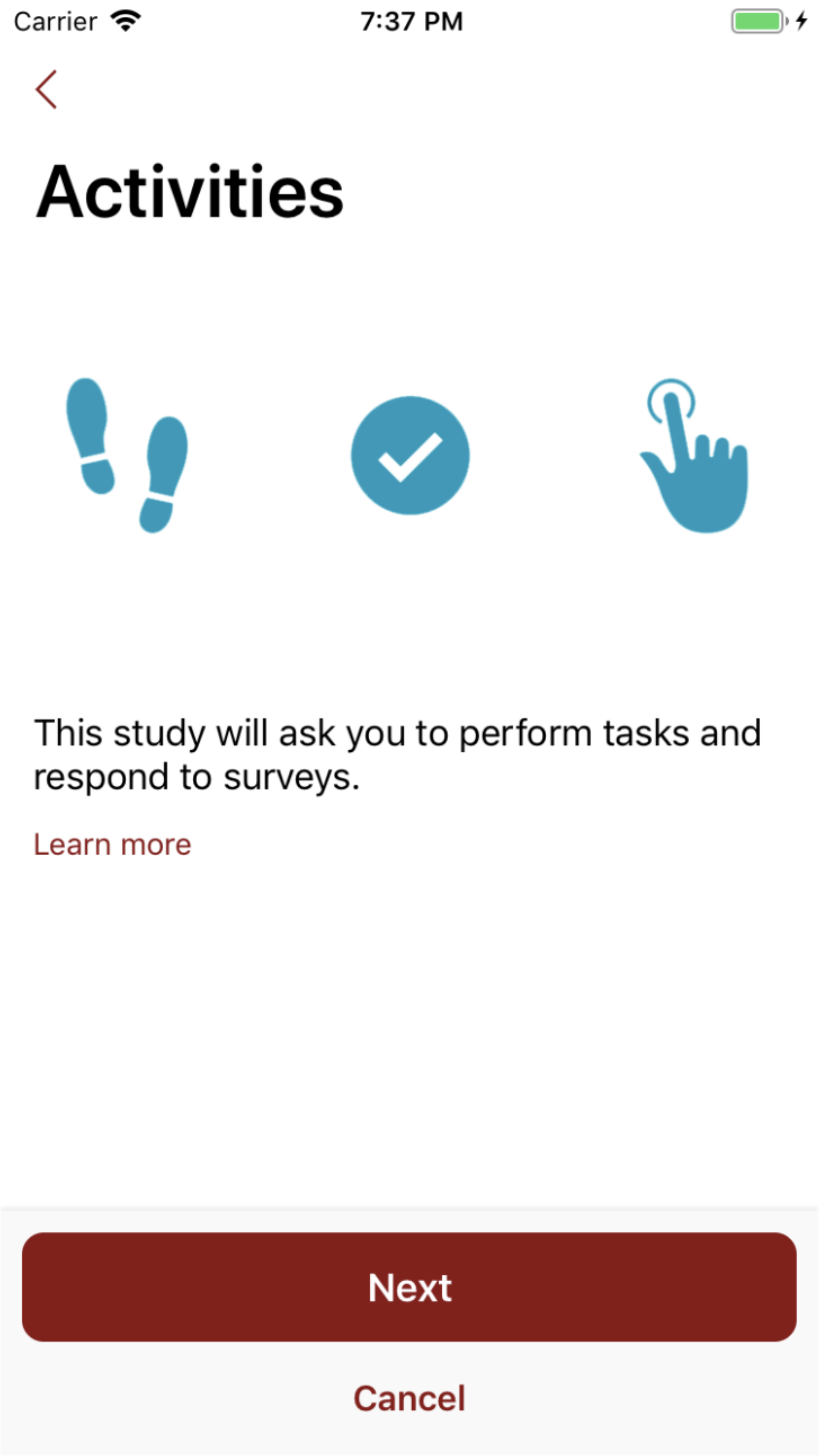
Carrier
7:37 PM
Activities
This study will ask you to perform tasks and respond to surveys.
Learn more
Next
Cancel

Carrier 7:37 PM

##### Activities

Done

The **MyHeart Counts** app will ask you to do 3 activities:

1. Use your phone, or any wearable activity device you have, to collect activity data for 7 days.
2. If you are able, perform a 6-minute walk test.
3. Enter information about risk factors and blood tests to calculate your risk score and 'heart age'.

Carrier 7:37 PM

### Sensor and Health Data

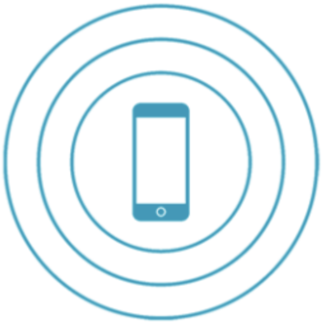

This study will also gather sensor and health data from your iPhone and personal devices with your permission.

Learn more

Next

Cancel

Carrier 7:37 PM

#### Sensor and Health Data

Done

There are sensors in your phone that can help assess activity, plus Apple's Health app on your phone can be linked with other devices to collect health and activity data, with your permission.

We will NOT access your personal contacts, other applications, personal photos, texts, or email messages.

Carrier 7:38 PM

### Data Processing

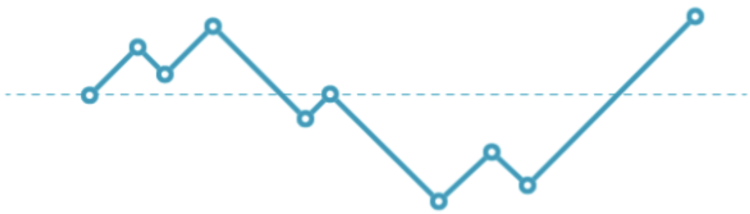

Collected data may allow researchers, as well as you, to understand patterns and details about heart health.

Next

Cancel

Carrier 7:38 PM

### Protecting your Data

Your data will be encrypted and sent to a secure database, with your name replaced by a random code.

Learn more about how your privacy and identity are protected

Next

Cancel

Carrier 7:39 PM

#### Protecting your Data

Done

We will use a random code instead of your name on all your study data. The coded study data are also encrypted and stored on a secure server to prevent improper access. This data server is run by Sage Bionetworks, a non-profit research organization. Stanford has secure servers that will maintain your consent and personal information.

Carrier 7:39 PM

### Data Use

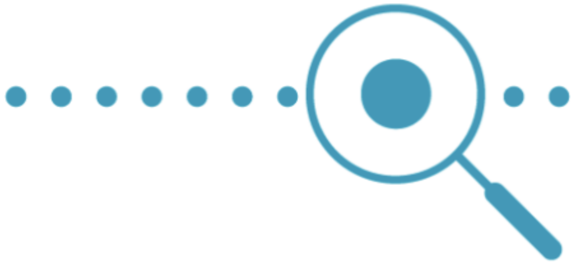

Your coded study data will be used for research by Stanford and may be shared with other researchers approved by Stanford.

Learn more about how data is used

Next

Cancel

Carrier 7:39 PM

Data Use Done

Your coded study data will be combined with data from other participants for analysis. This provides a rich database for research. It also provides a safe way to share the data with other researchers approved by Stanford to learn more about cardiovascular disease. Study data will never be sold to any third party.

Carrier 7:39 PM

### Issues to Consider

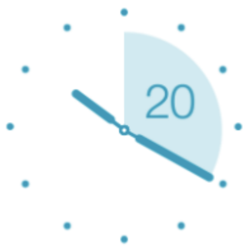

Your initial participation in this study will take 10-15 minutes per day for a week. We hope that you can contribute to the study for one week every three months.

Learn more about the study's impact on your time

Next

Cancel

Carrier 7:39 PM

Issues to Consider Done

We will ask you survey questions and have you use your phone or wearable device to collect activity data for 7 days. After that, if you are able, there is a 6-minute walk test. You will also need to have your blood pressure and cholesterol information in order to determine your risk score and 'heart age'. You can continue to use the app for activity monitoring and it will ask you to update your data every 3 months.

Carrier 7:39 PM

### Surveys

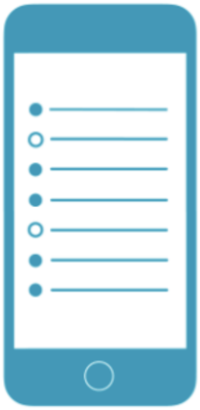

Some of the tasks in this study will require you to answer survey questions about health and lifestyle factors.

Next

Cancel

Carrier 7:39 PM

### Study Tasks

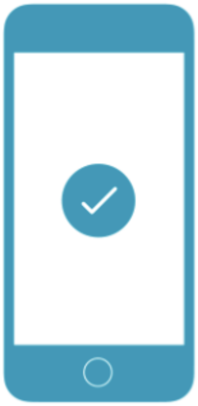

We will ask you to complete active tasks that may require physical activity.

Learn more about the tasks involved

Next

Cancel

Carrier 7:39 PM

**Study Tasks** **Done**

You will be asked to do a Physical Activity Readiness questionnaire, which will advise you about talking to your doctor before increasing physical activity or assessing fitness.

The 7-day activity and sleep assessment will use the sensors in your phone or wearable device in conjunction with the Health app on your iPhone. You will be asked to keep your phone or wearable with you for the 7 days and do your regular activities. There will be a daily check of your sleep and any missed activities.

If you are able, after the 7 days there is a 6-minute walk test as an assessment of fitness. This uses the phone (or wearable) to measure the distance you can walk in 6 minutes. This is the only time the app may use GPS information, and only to help measure distance. Your location will not be stored.

Carrier 7:39 PM

<

### Withdrawing

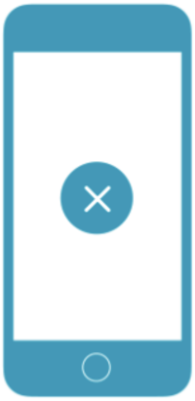

You may withdraw your consent and discontinue participation at any time.

Learn more about withdrawing

Next

Cancel

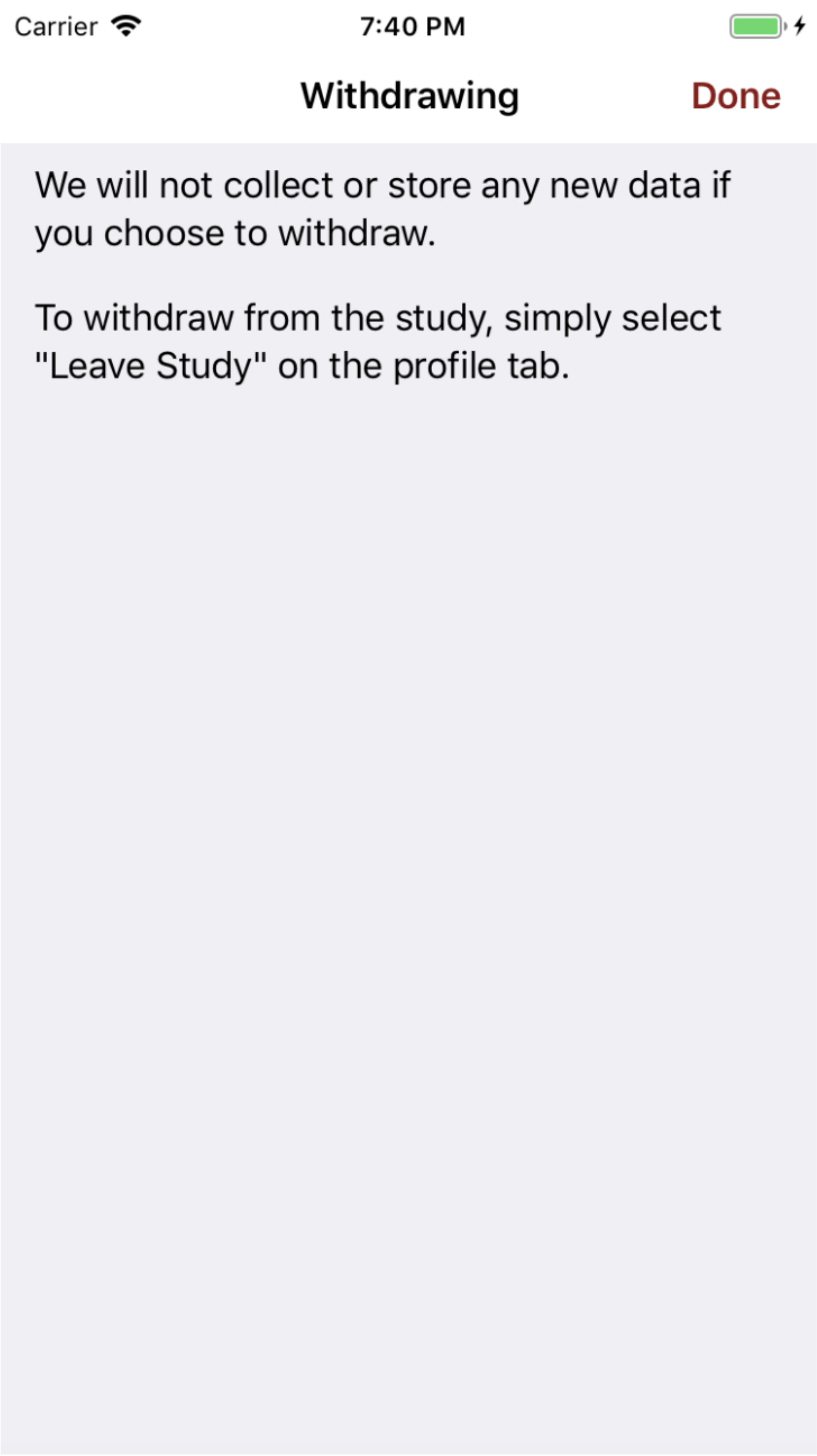
Carrier
7:40 PM
Withdrawing
Done
We will not collect or store any new data if you choose to withdraw.
To withdraw from the study, simply select "Leave Study" on the profile tab.

Carrier 7:40 PM

<

### Potential Benefits

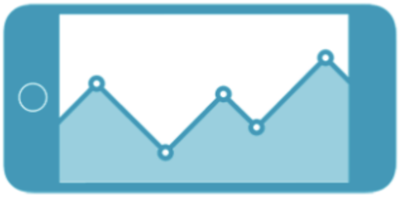

The information collected by this study may help you better understand and monitor your heart health.

Learn more

Next

Cancel

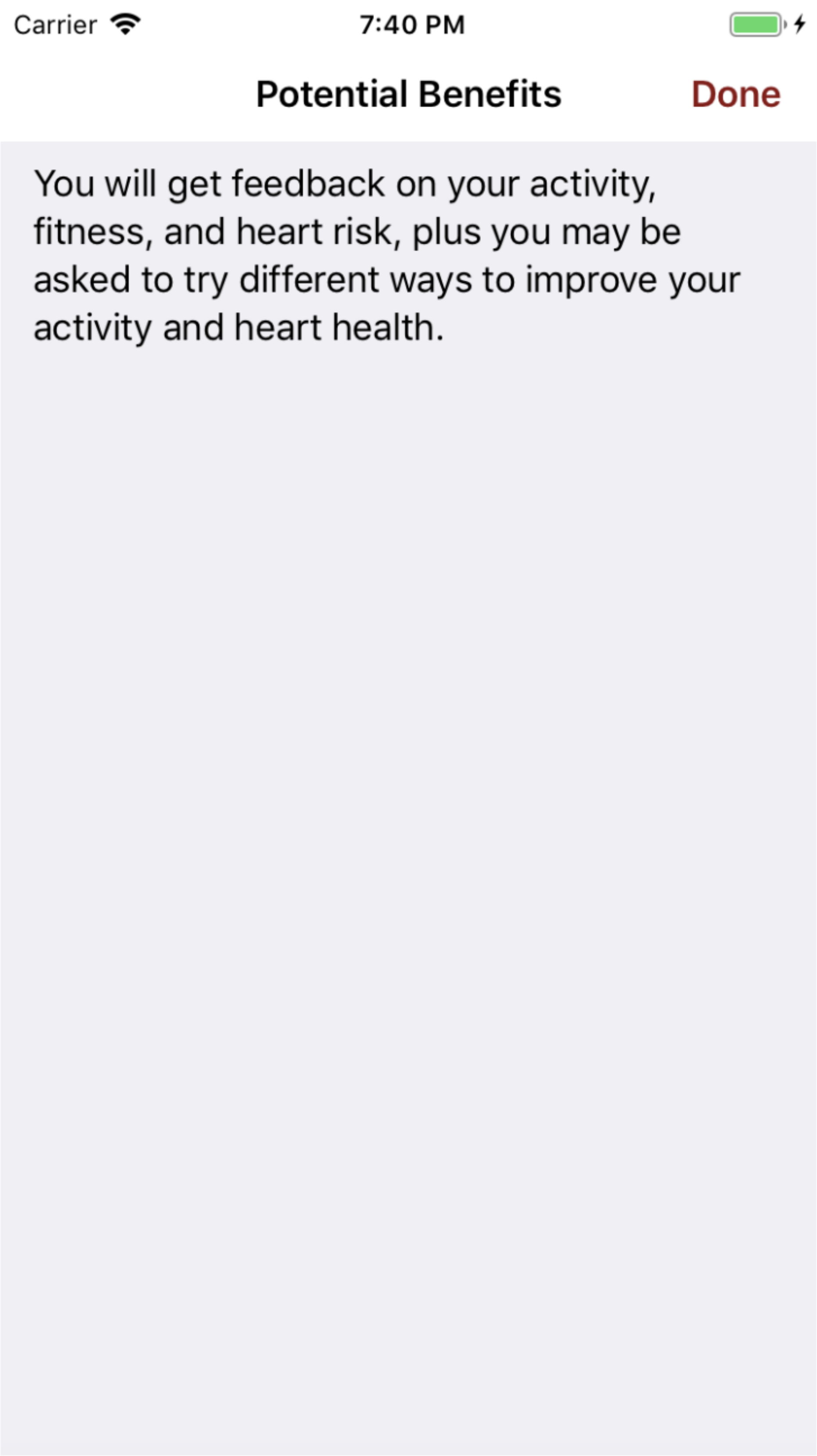
Carrier
7:40 PM
Potential Benefits
Done
You will get feedback on your activity, fitness, and heart risk, plus you may be asked to try different ways to improve your activity and heart health.

Carrier 7:42 PM

### Issues to Consider

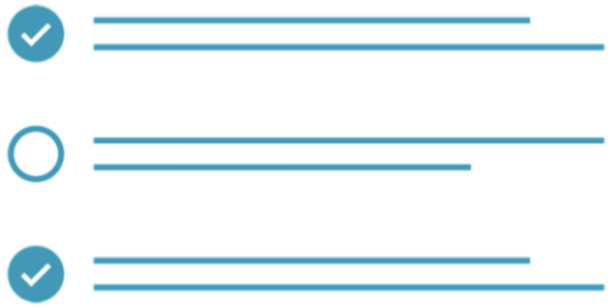

Some questions may make you uncomfortable.  Simply do not respond.

Next

Cancel

Carrier 7:42 PM

### Issues to Consider

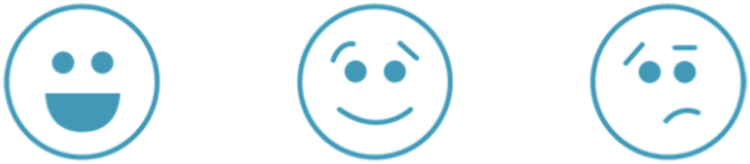

Participating in this study may change how you feel. You may feel more tired, sad, energized, or happy.

Learn more

Next

Cancel

Carrier 7:42 PM

**Issues to Consider** **Done**

The MyHeart Counts app will provide data about your activity, fitness, and cardiovascular risk. These results could certainly generate a wide range of emotions.

Carrier 7:42 PM

### Risk to Privacy

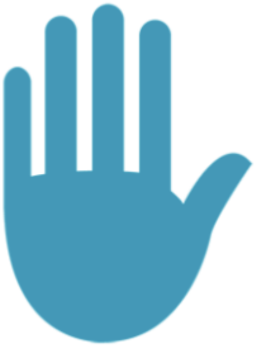

We will make every effort to protect your information, but total anonymity cannot be guaranteed. Regardless of where you are physically located when you use the app, your data will be sent to the United States and potentially other countries where laws may not protect your privacy to the same extent as in the country from which you sent the data.

Learn more

Next

Cancel

Carrier 7:42 PM

#### Risk to Privacy

Done

We view your privacy very seriously. Thus, we are requesting the least amount of personal data possible. Also, we are using strict security protocols to protect your data. Importantly, your name will be replaced with a random code before the data goes to the large computer for storage and later analysis. We cannot completely guarantee that someone cannot gain access to your private data, but importantly the main data storage is secure, encrypted, and does not contain your name.

The app is currently intended for use in only the United States, the United Kingdom, and Hong Kong. If you travel outside of these countries during the study, then you should not use the app during such travels.

As described elsewhere in the informed consent form, during the study, data pertaining to your participation in the study will be generated and recorded, including data about your health. We refer to such data as "Your Study Data." Your Study Data may be processed or used for the following purposes, which we refer to, collectively, as "Data Processing":

“Data Processing”:

- to carry out the study;
- to confirm the accuracy of the study;
- to monitor that the study complies with applicable laws as well as best practices developed by the research community;
- to comply with legal and regulatory requirements, including requirements that data from this study, without information that could directly identify you, be made available to other researchers not affiliated with the study sponsor or with the study team.

It is possible, for example, that as part of efforts to make research data more widely available to researchers, regulatory authorities in some countries may require that Your Study Data, without information that could directly identify you, be made publicly available on the internet or in other ways. The following entities and organizations may engage in Data Processing that uses Your Study Data:

Processing that uses Your Study Data:

- the study team, including other people who, and organizations that, assist the study team;
- the ethics committee or institutional review board that approved this study;
- Sage Bionetworks, a research entity that operates the secure cloud server in which Your Study Data are stored, and any other similar entities that are used to store or protect your data; and
- domestic and foreign regulatory agencies and government officials who have a duty to monitor or oversee studies like this one.

Some of the entities listed above that receive Your Study Data for Data Processing are located in the United States and potentially in other countries where, the laws do not protect your privacy to the same extent as the laws in your country of residence. In such cases, Data Processing that involves Your Study Data may be subject to the less restrictive data protection laws of these foreign countries rather than the laws of your own country. However, all reasonable steps will be taken to protect your privacy.

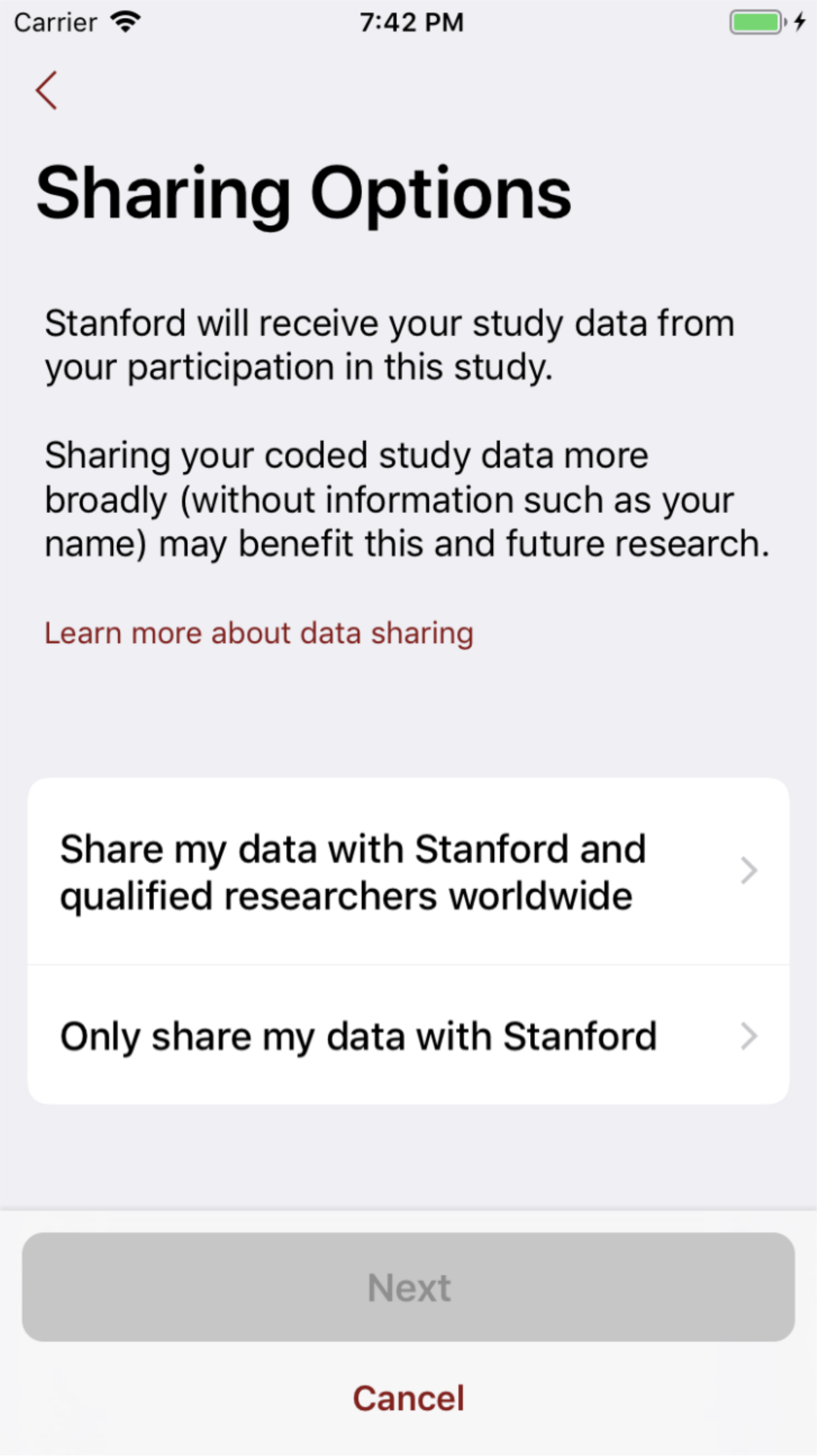
Carrier
7:42 PM
Sharing Options
Stanford will receive your study data from your participation in this study.
Sharing your coded study data more broadly (without information such as your name) may benefit this and future research.
Learn more about data sharing
Share my data with Stanford and qualified researchers worldwide
Only share my data with Stanford
Next
Cancel

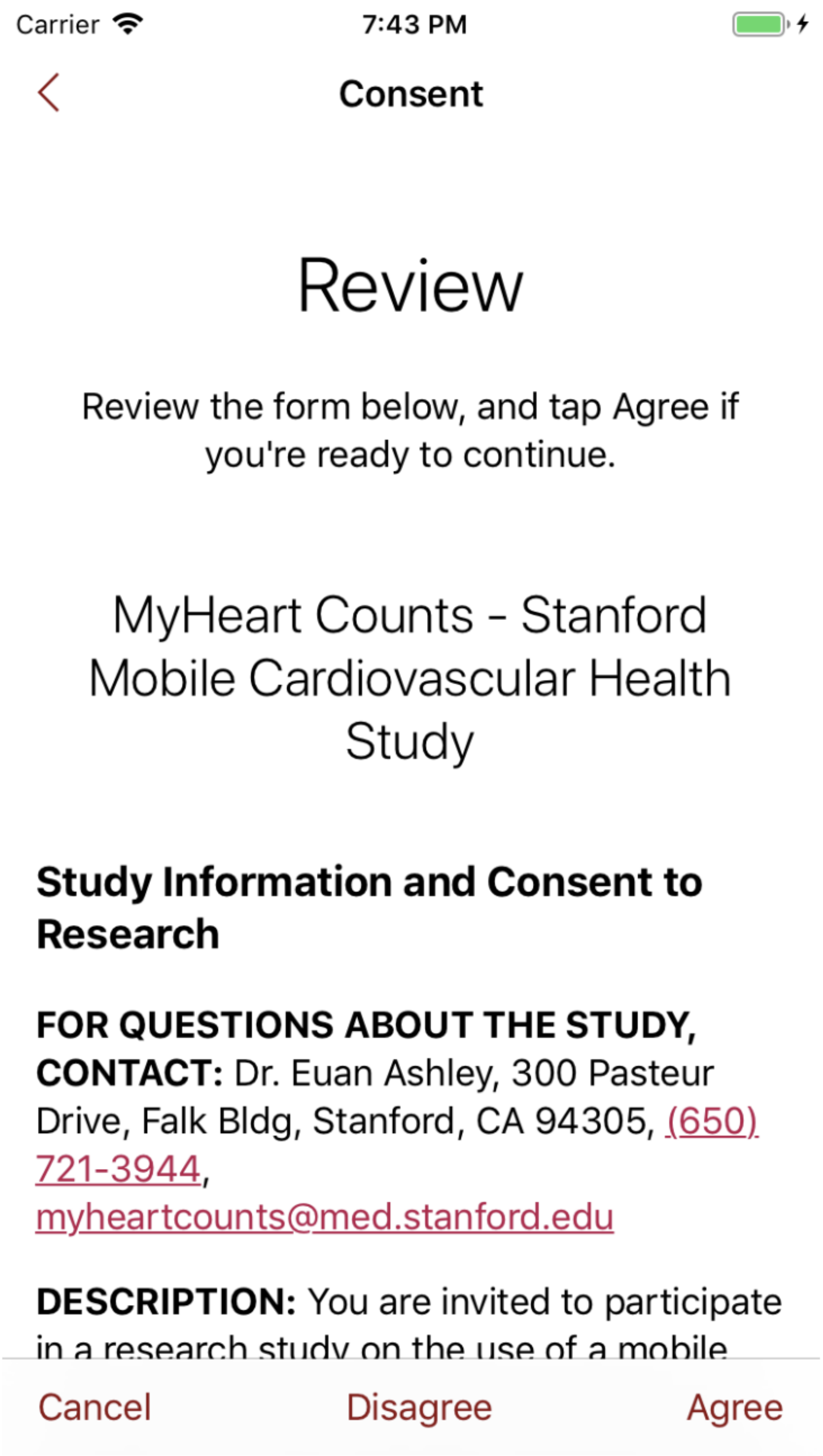
Carrier
7:43 PM
Consent
Review
Review the form below, and tap Agree if you're ready to continue.
MyHeart Counts - Stanford Mobile Cardiovascular Health Study
Study Information and Consent to Research
FOR QUESTIONS ABOUT THE STUDY, CONTACT: Dr. Euan Ashley, 300 Pasteur Drive, Falk Bldg, Stanford, CA 94305, (650) 721-3944,
DESCRIPTION: You are invited to participate
Cancel
Disagree
Agree

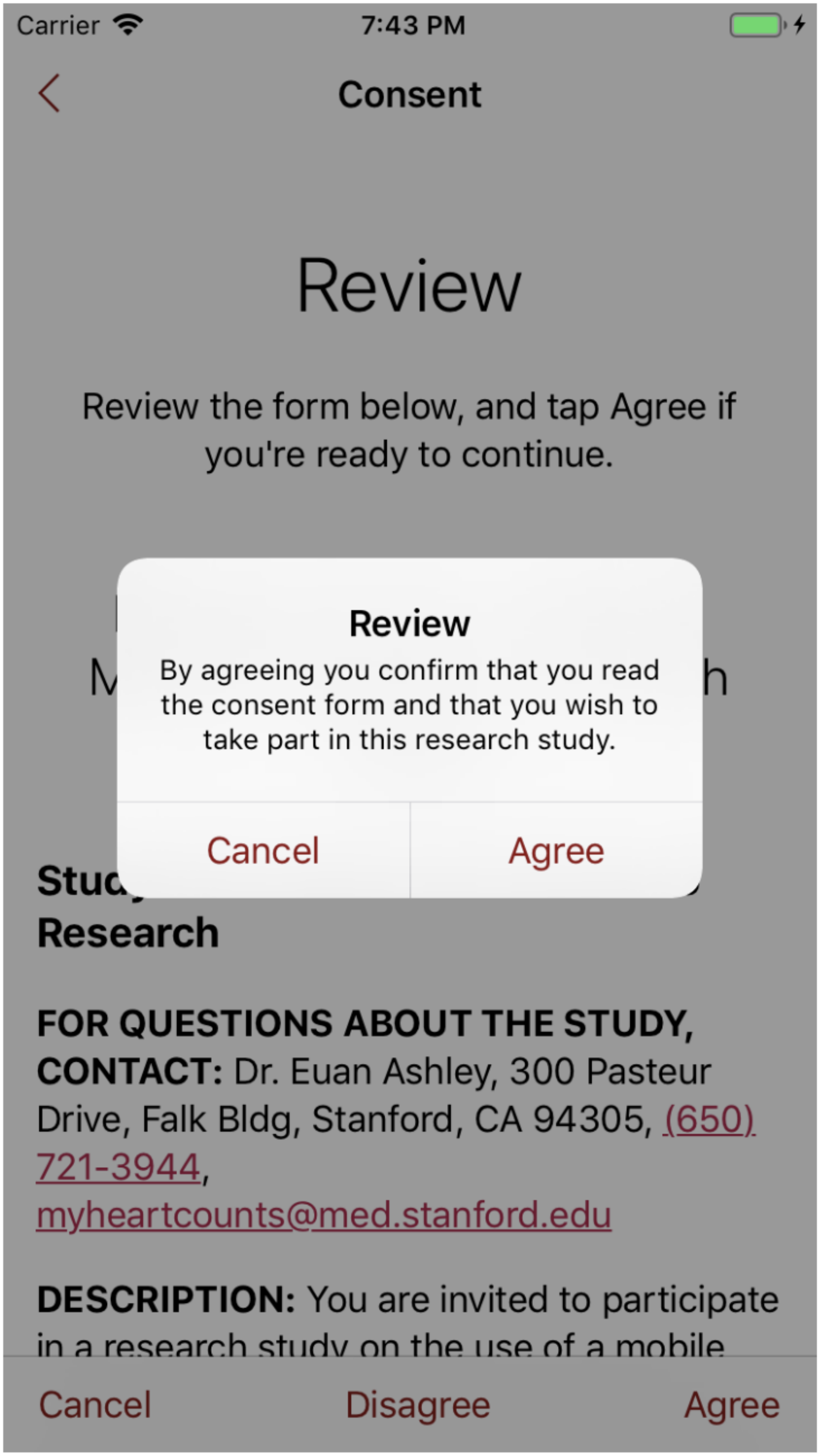
Carrier
7:43 PM
Consent
Review
Review the form below, and tap Agree if you're ready to continue.
Review
By agreeing you confirm that you read the consent form and that you wish to take part in this research study.
Cancel
Agree
Research
FOR QUESTIONS ABOUT THE STUDY, CONTACT: Dr. Euan Ashley, 300 Pasteur Drive, Falk Bldg, Stanford, CA 94305, (650) 721-3944,
DESCRIPTION: You are invited to participate
Cancel
Disagree
Agree

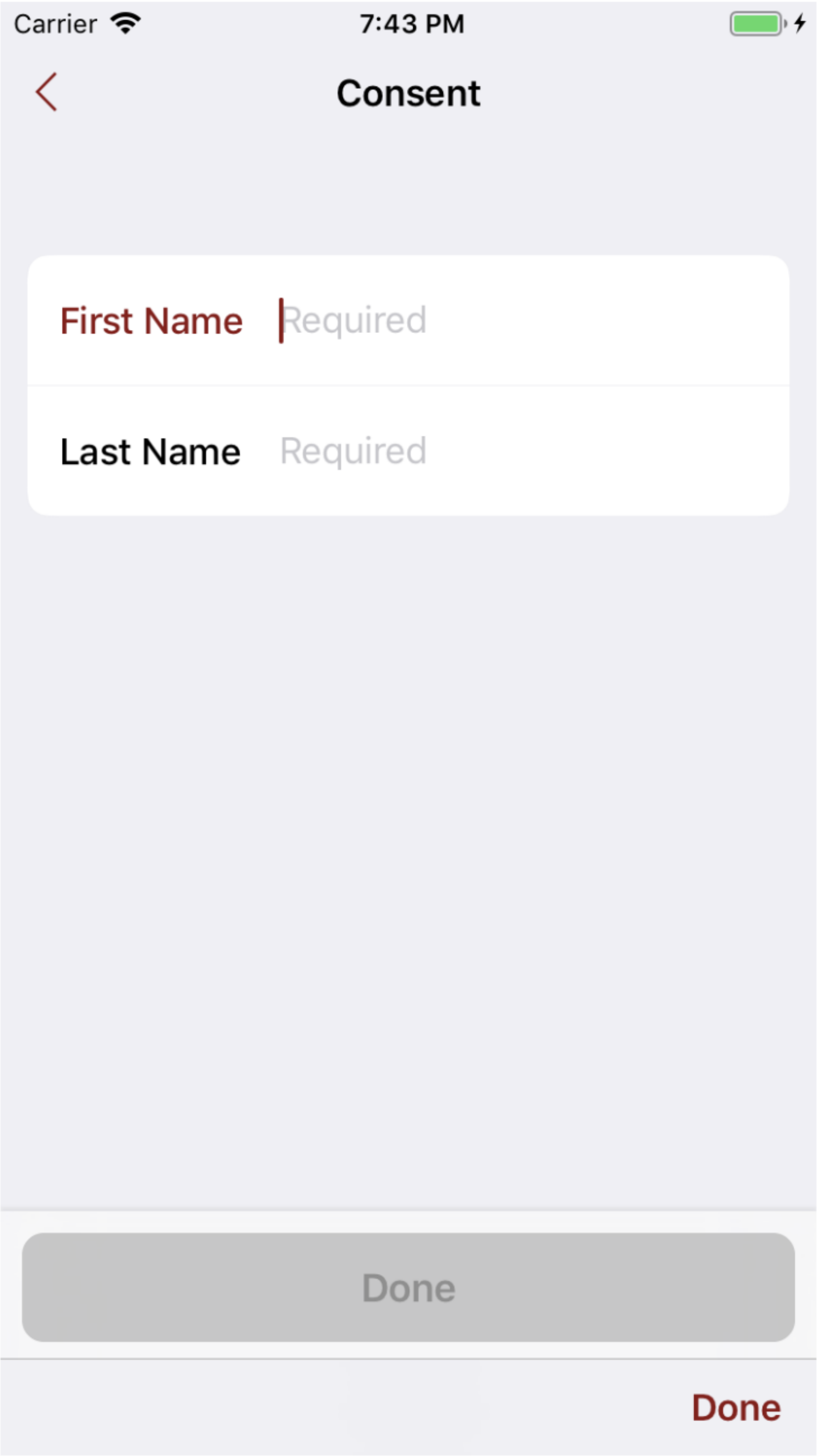
Carrier
7:43 PM
Consent
First Name
Required
Last Name
Required
Done
Done

### SCREENSHOTS OF THE CLUSTER RANDOMIZED TRIAL CONSENT

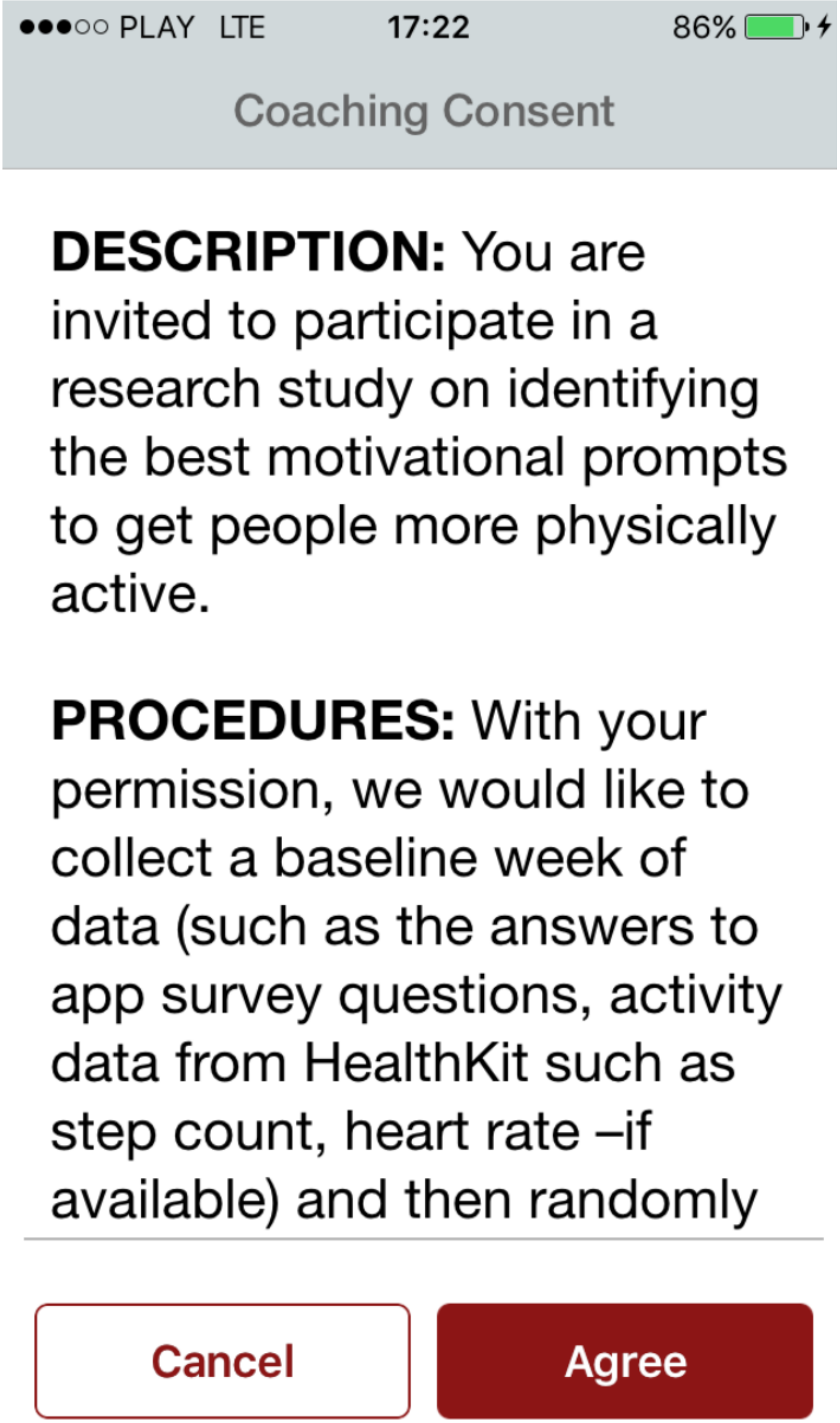
●●●○○ PLAY LTE
17:22
86%
Coaching Consent
DESCRIPTION: You are invited to participate in a research study on identifying the best motivational prompts to get people more physically active.
PROCEDURES: With your permission, we would like to collect a baseline week of data (such as the answers to app survey questions, activity data from HealthKit such as step count, heart rate –if available) and then randomly
Cancel
Agree

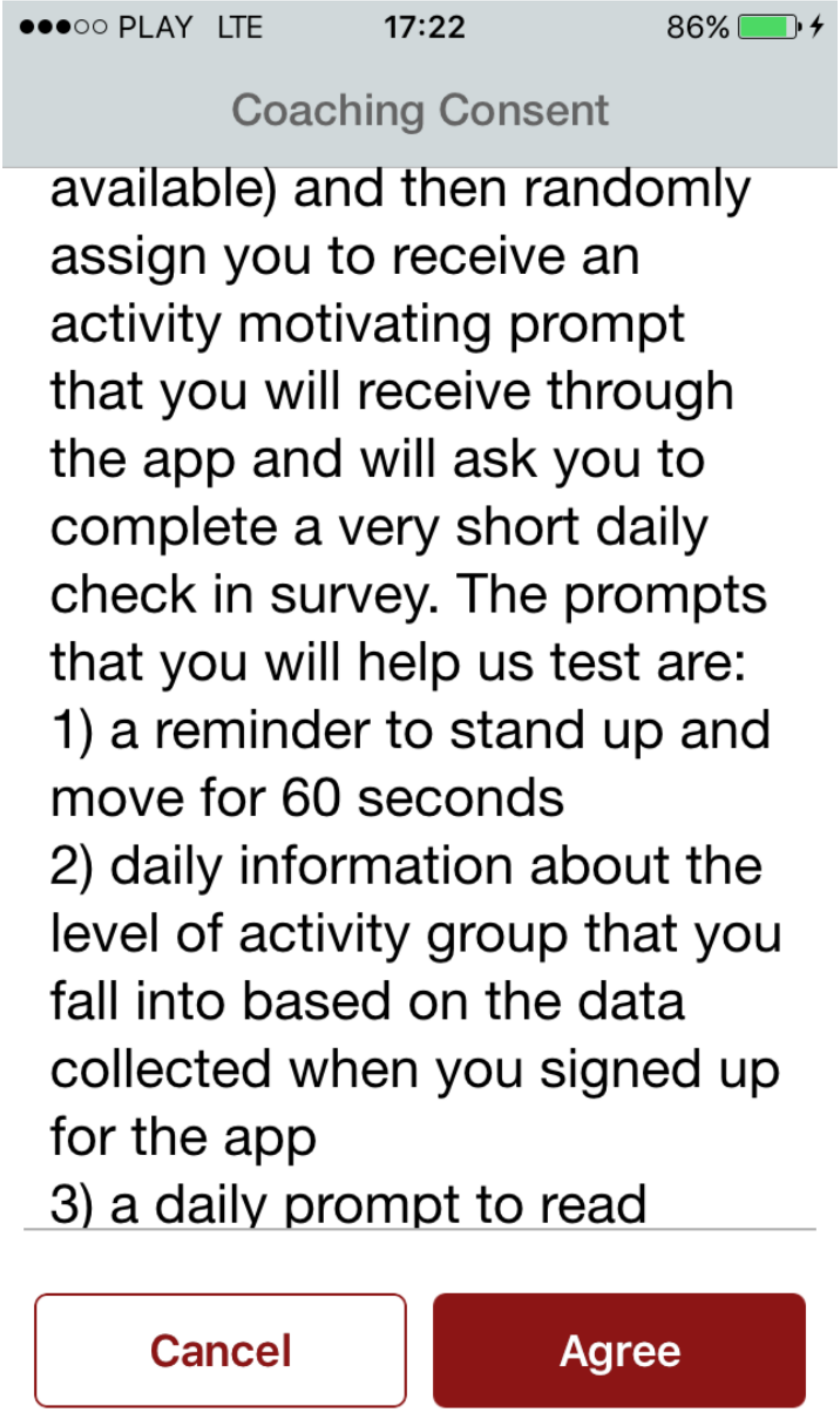
●●●○○ PLAY LTE
17:22
86%
Coaching Consent
available) and then randomly assign you to receive an activity motivating prompt that you will receive through the app and will ask you to complete a very short daily check in survey. The prompts that you will help us test are:
1) a reminder to stand up and move for 60 seconds
2) daily information about the level of activity group that you fall into based on the data collected when you signed up for the app
3) a daily prompt to read
Cancel
Agree

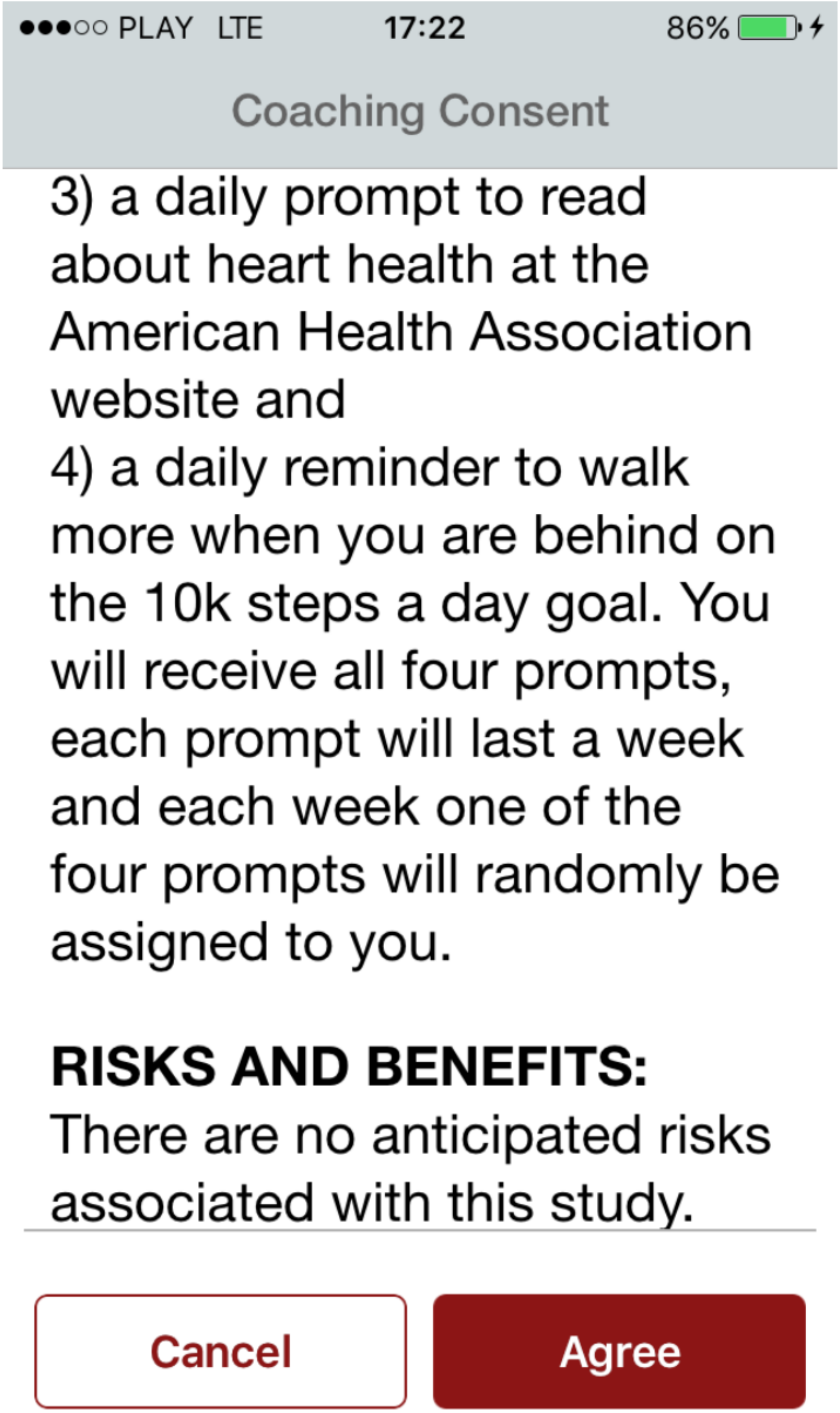
●●●○○ PLAY LTE 17:22 86%
Coaching Consent
3) a daily prompt to read about heart health at the American Health Association website and
4) a daily reminder to walk more when you are behind on the 10k steps a day goal. You will receive all four prompts, each prompt will last a week and each week one of the four prompts will randomly be assigned to you.
RISKS AND BENEFITS:
There are no anticipated risks associated with this study.
Cancel
Agree

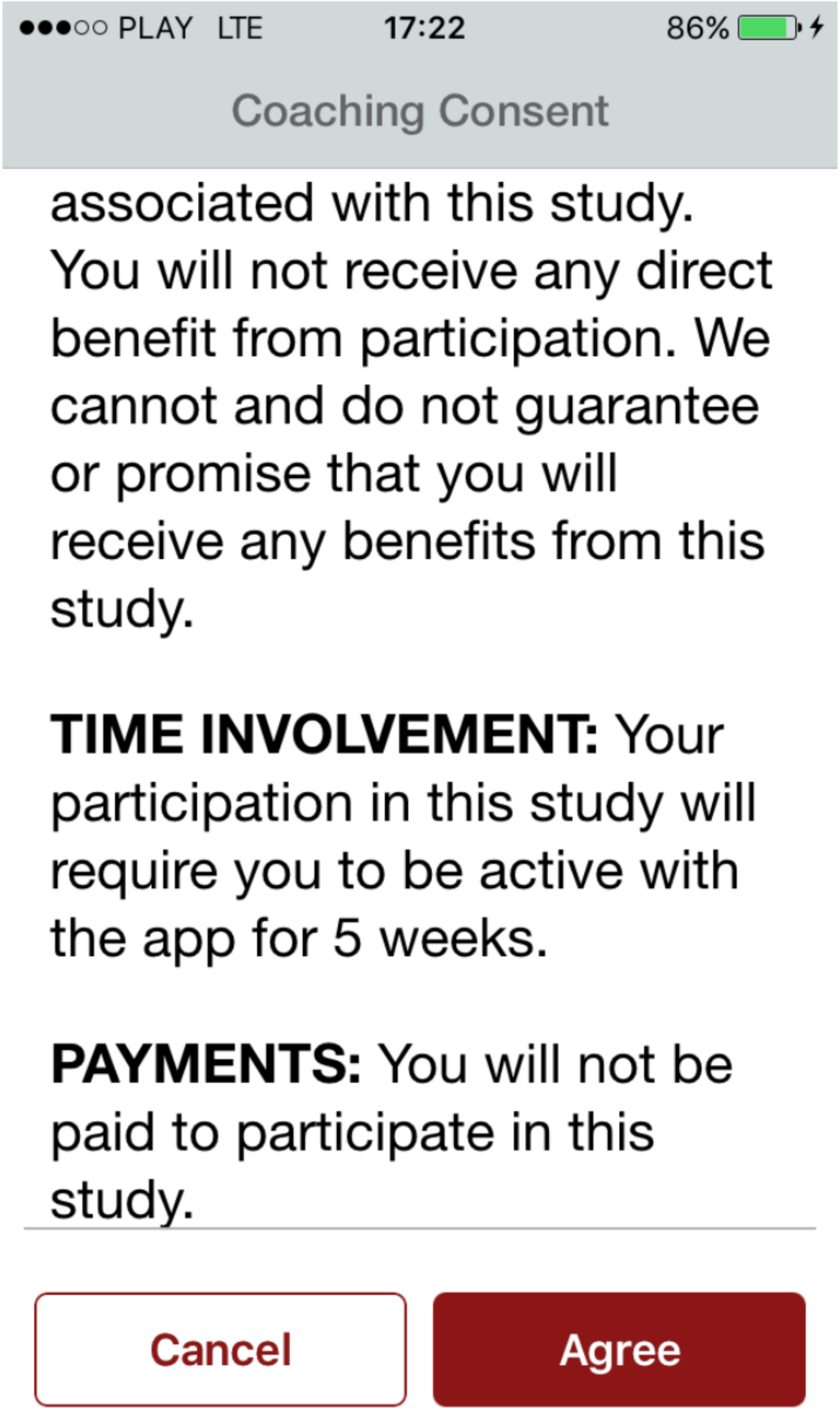
●●●○○ PLAY LTE 17:22 86%
Coaching Consent
associated with this study. You will not receive any direct benefit from participation. We cannot and do not guarantee or promise that you will receive any benefits from this study.
**TIME INVOLVEMENT:** Your participation in this study will require you to be active with the app for 5 weeks.
**PAYMENTS:** You will not be paid to participate in this study.
Cancel
Agree

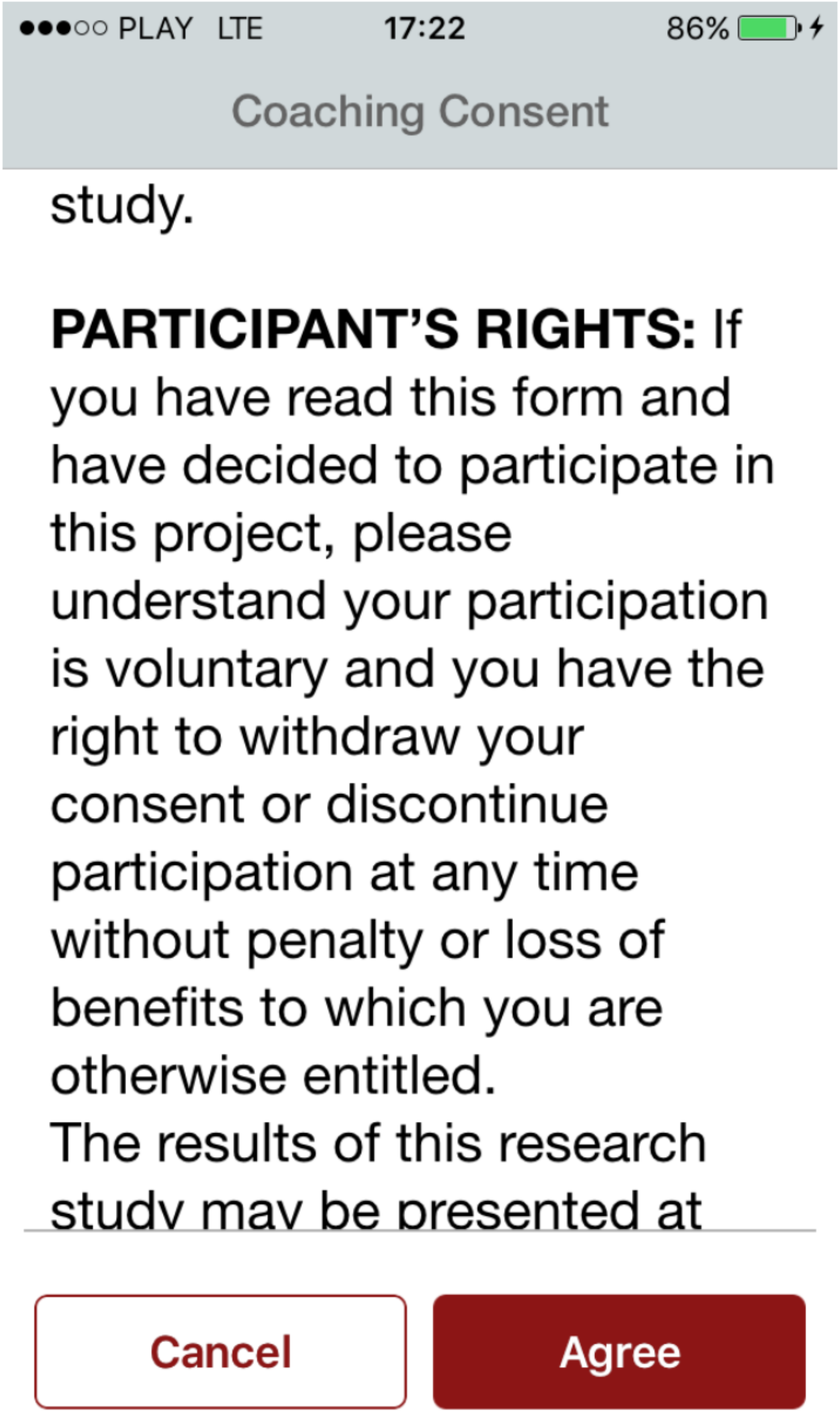
●●●○○ PLAY LTE
17:22
86%
Coaching Consent
study.
PARTICIPANT'S RIGHTS: If you have read this form and have decided to participate in this project, please understand your participation is voluntary and you have the right to withdraw your consent or discontinue participation at any time without penalty or loss of benefits to which you are otherwise entitled.
The results of this research study may be presented at
Cancel
Agree

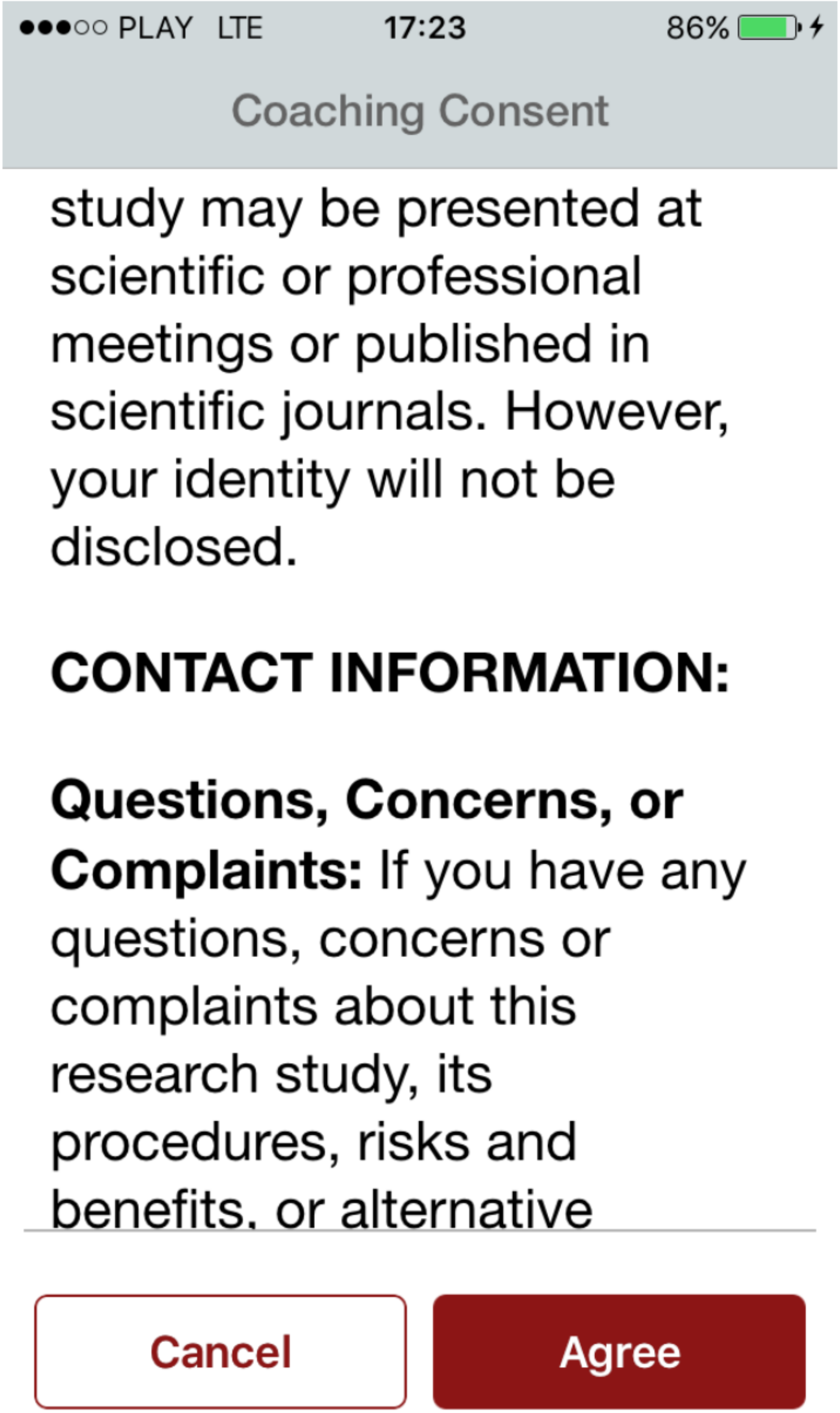
●●●○○ PLAY LTE 17:23 86%
Coaching Consent
study may be presented at scientific or professional meetings or published in scientific journals. However, your identity will not be disclosed.
CONTACT INFORMATION:
Questions, Concerns, or Complaints: If you have any questions, concerns or complaints about this research study, its procedures, risks and benefits, or alternative
Cancel
Agree

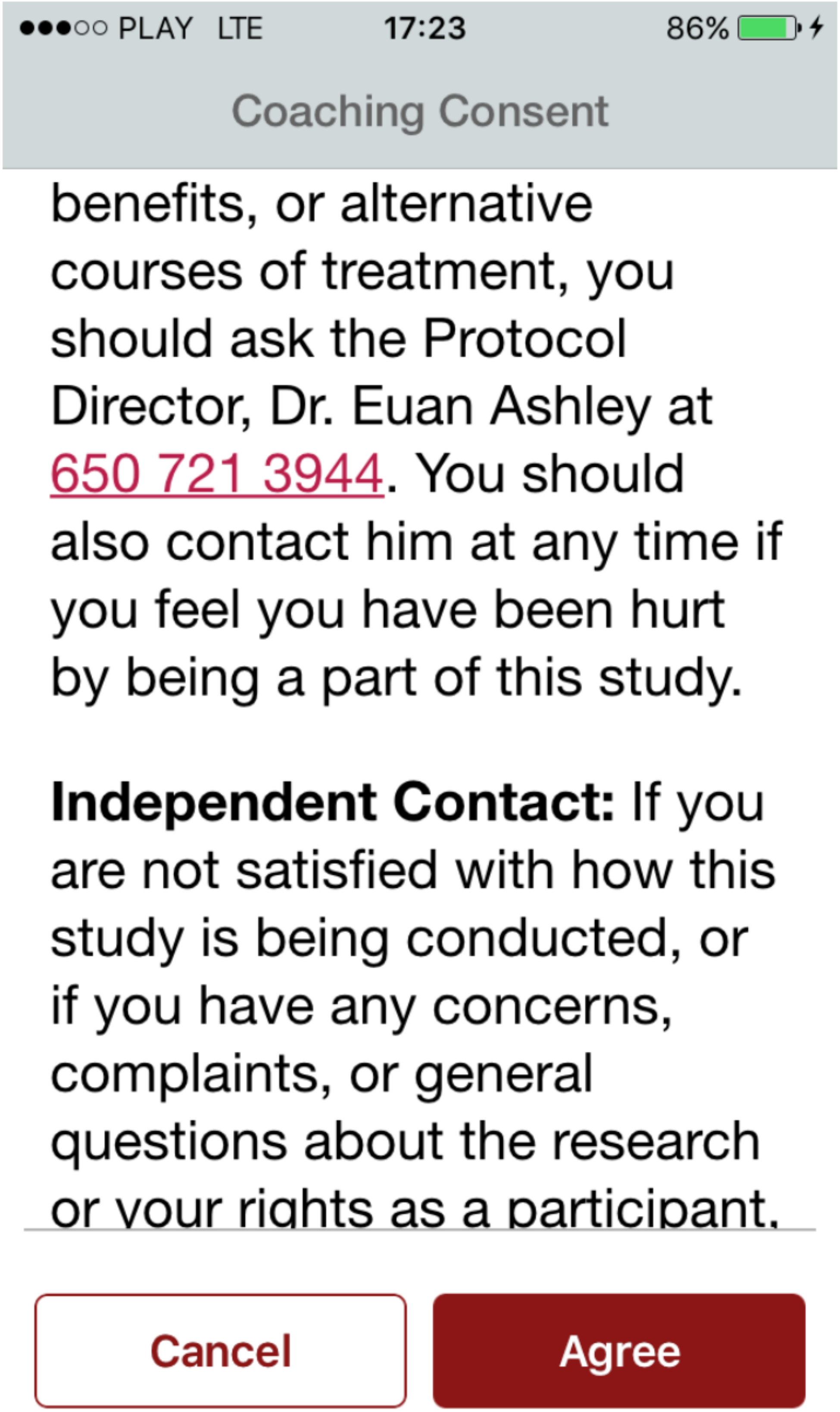
●●●○○ PLAY LTE 17:23 86%
Coaching Consent
benefits, or alternative courses of treatment, you should ask the Protocol Director, Dr. Euan Ashley at 650 721 3944. You should also contact him at any time if you feel you have been hurt by being a part of this study.
**Independent Contact:** If you are not satisfied with how this study is being conducted, or if you have any concerns, complaints, or general questions about the research or your rights as a participant,
Cancel
Agree

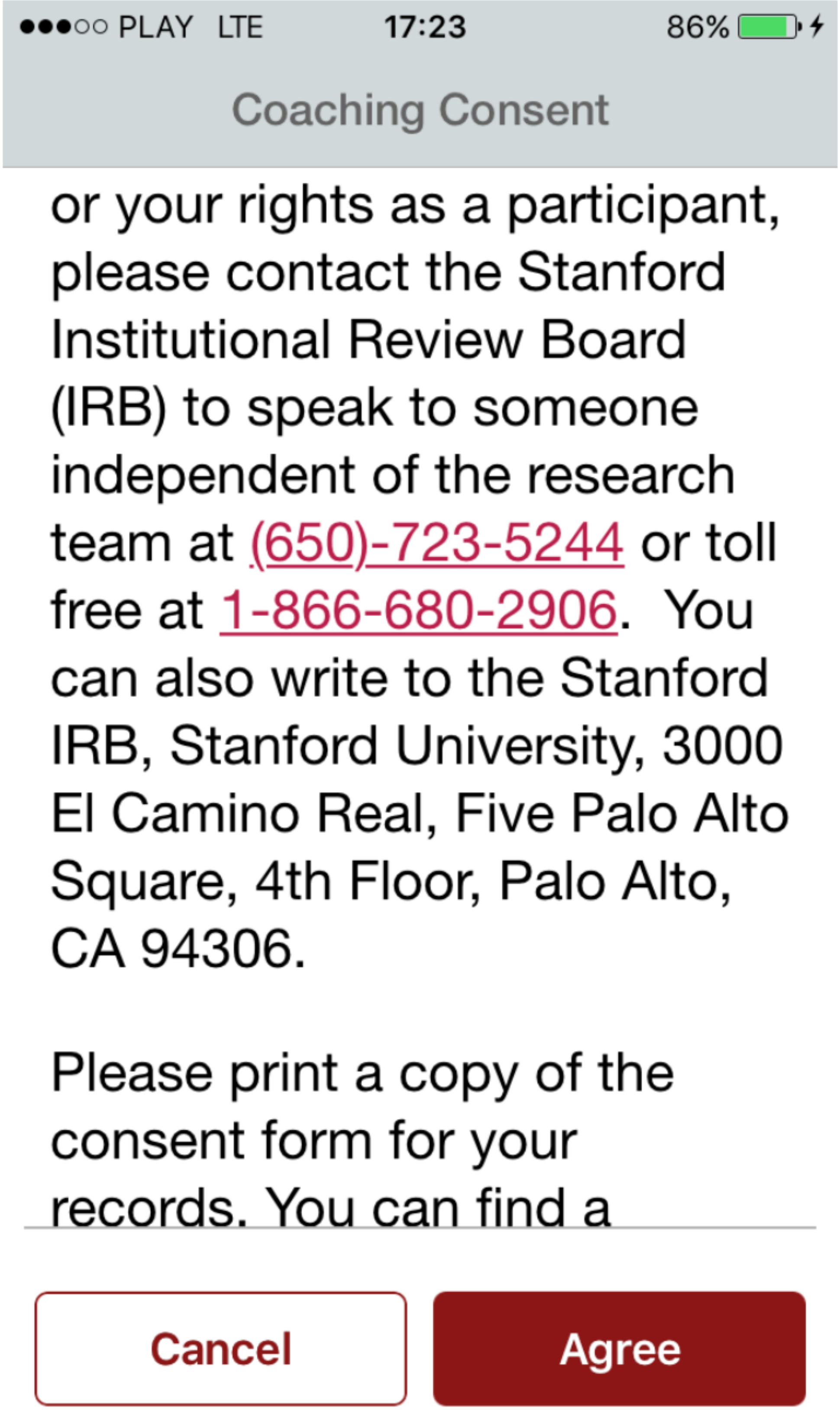
●●●○○ PLAY LTE
17:23
86%
Coaching Consent
or your rights as a participant, please contact the Stanford Institutional Review Board (IRB) to speak to someone independent of the research team at (650)-723-5244 or toll free at 1-866-680-2906. You can also write to the Stanford IRB, Stanford University, 3000 El Camino Real, Five Palo Alto Square, 4th Floor, Palo Alto, CA 94306.
Please print a copy of the consent form for your records. You can find a
Cancel
Agree

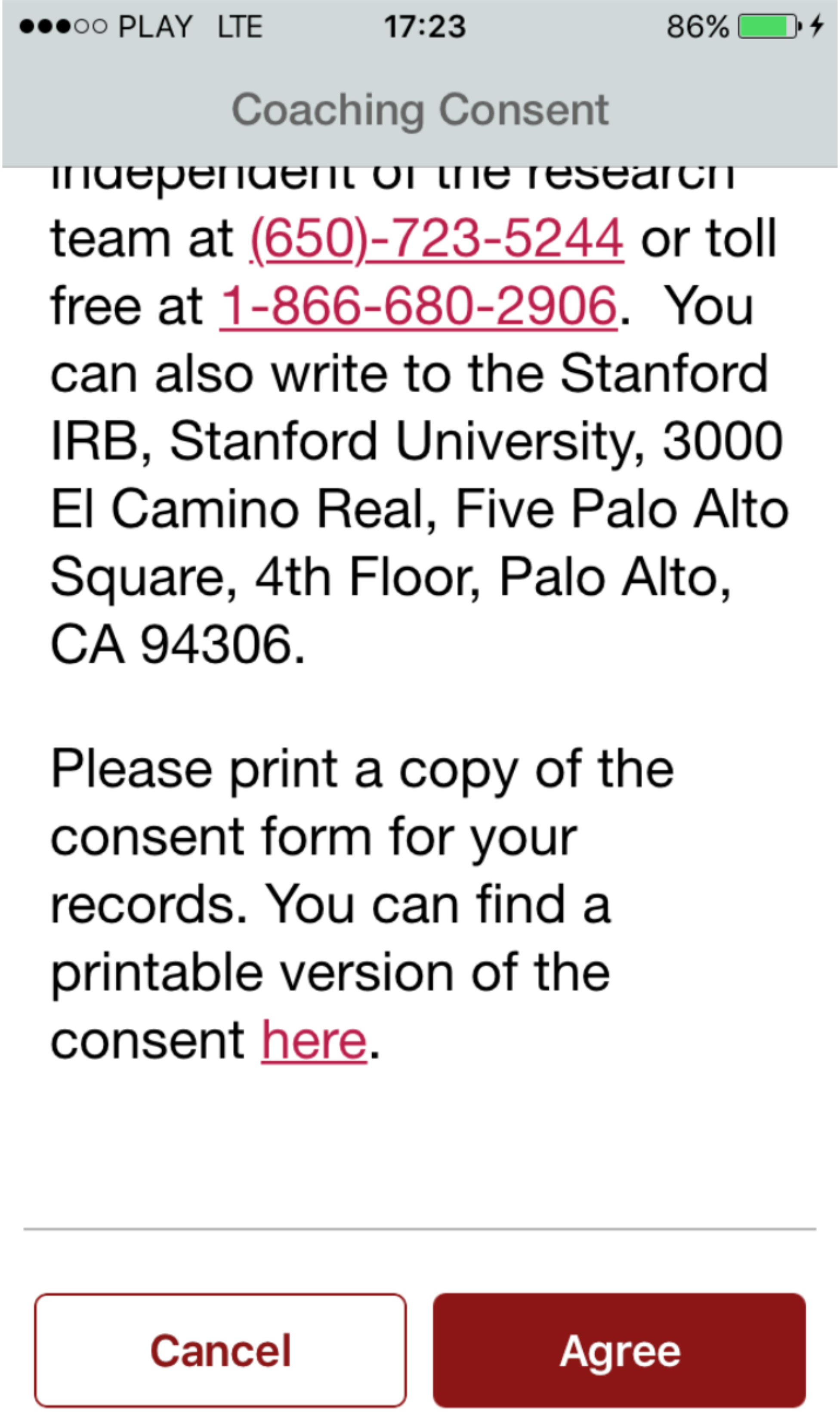
●●●○○ PLAY LTE
17:23
86%
Coaching Consent
team at (650)-723-5244 or toll free at 1-866-680-2906. You can also write to the Stanford IRB, Stanford University, 3000 El Camino Real, Five Palo Alto Square, 4th Floor, Palo Alto, CA 94306.
Please print a copy of the consent form for your records. You can find a printable version of the consent here.
Cancel
Agree

●●●○○ PLAY LTE 17:24 87%

#### Stay Active!

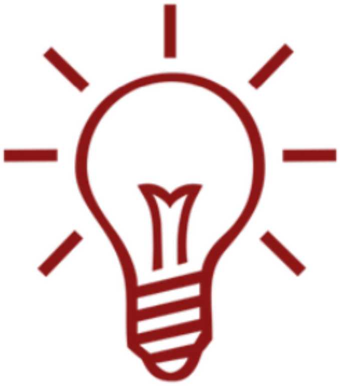

Thank you for taking part in this study.
Since we are testing for the efficacy of each module, it is important to turn off any other device prompts/reminders you may have set up (such as in Fitbit/Jawbone etc.), so that the data is not muddy. We remind you to please keep your [illegible]

**Next**

●●●○○ PLAY LTE 17:24 87%

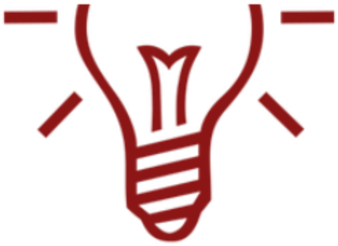

Thank you for taking part in this study.
Since we are testing for the efficacy of each module, it is important to turn off any other device prompts/ reminders you may have set up (such as in Fitbit/Jawbone etc.), so that the data is not muddy. We remind you to please keep your phone on you and MyHeart Counts running during the next 4 weeks.

Next

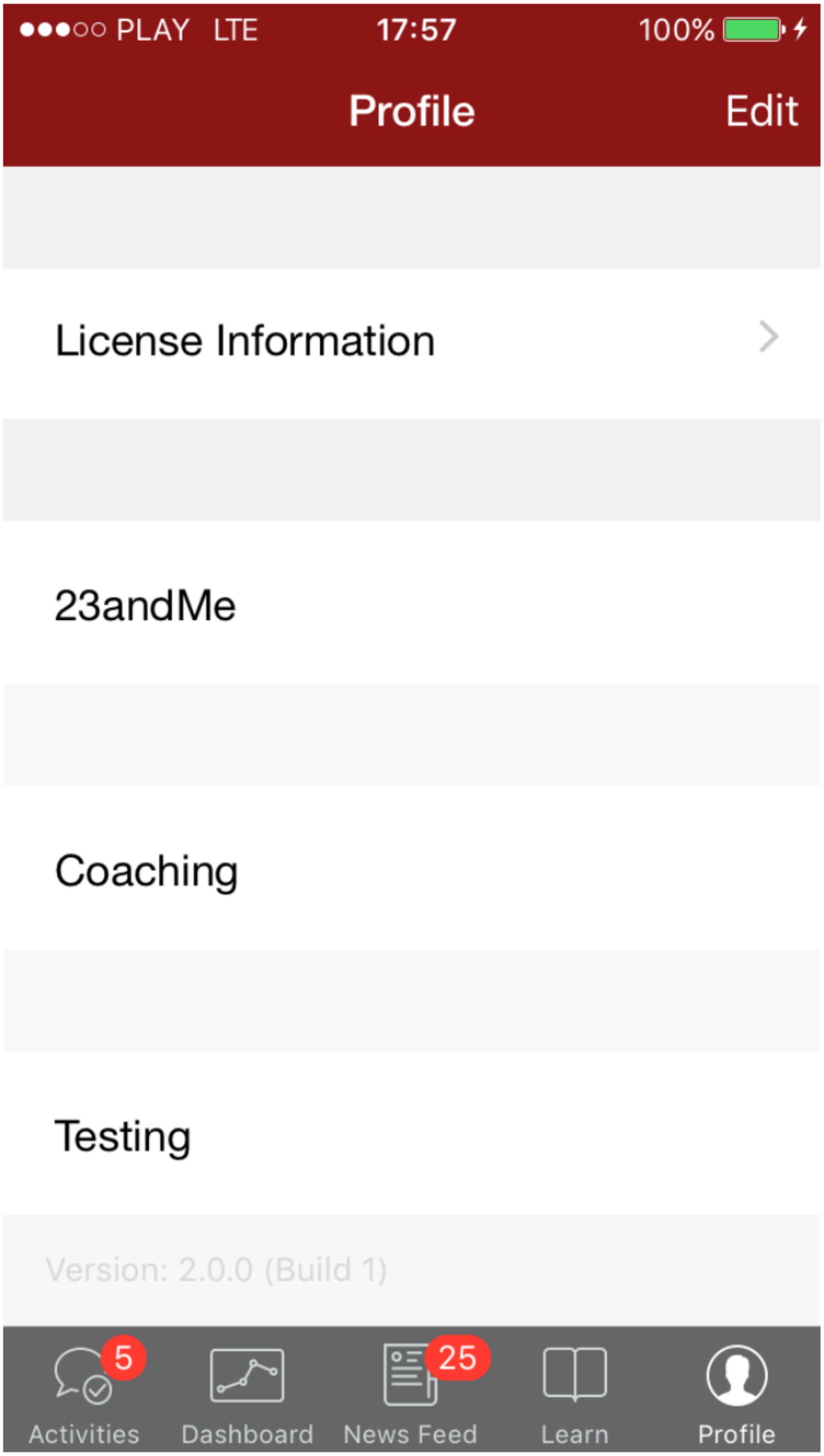
●●●○○ PLAY LTE
17:57
100%
Profile
Edit
License Information
23andMe
Coaching
Testing
Version: 2.0.0 (Build 1)
5
25
Activities
Dashboard
News Feed
Learn
Profile

●●●○○ PLAY LTE 17:58 100%

Coaching Consent Close

**DESCRIPTION:** You are invited to participate in a research study on identifying the best motivational prompts to get people more physically active.

**PROCEDURES:** With your permission, we would like to collect a baseline week of data (such as the answers to app survey questions, activity data from HealthKit such as step count, heart rate –if available) and then randomly assign you to receive an activity motivating prompt that you will receive through the app and will ask you to

●●●○○ PLAY LTE 17:58 100%

Coaching Consent Close

that you will receive through the app and will ask you to complete a very short daily check in survey. The prompts that you will help us test are:

1) a reminder to stand up and move for 60 seconds
2) daily information about the level of activity group that you fall into based on the data collected when you signed up for the app
3) a daily prompt to read about heart health at the American Health Association website and
4) a daily reminder to walk more when you are behind on the 10k steps a day goal. You

●●●○○ PLAY LTE 17:58 100%

Coaching Consent Close

the 10k steps a day goal. You will receive all four prompts, each prompt will last a week and each week one of the four prompts will randomly be assigned to you.

**RISKS AND BENEFITS:** There are no anticipated risks associated with this study. You will not receive any direct benefit from participation. We cannot and do not guarantee or promise that you will receive any benefits from this study.

**TIME INVOLVEMENT:** Your participation in this study will [illegible]

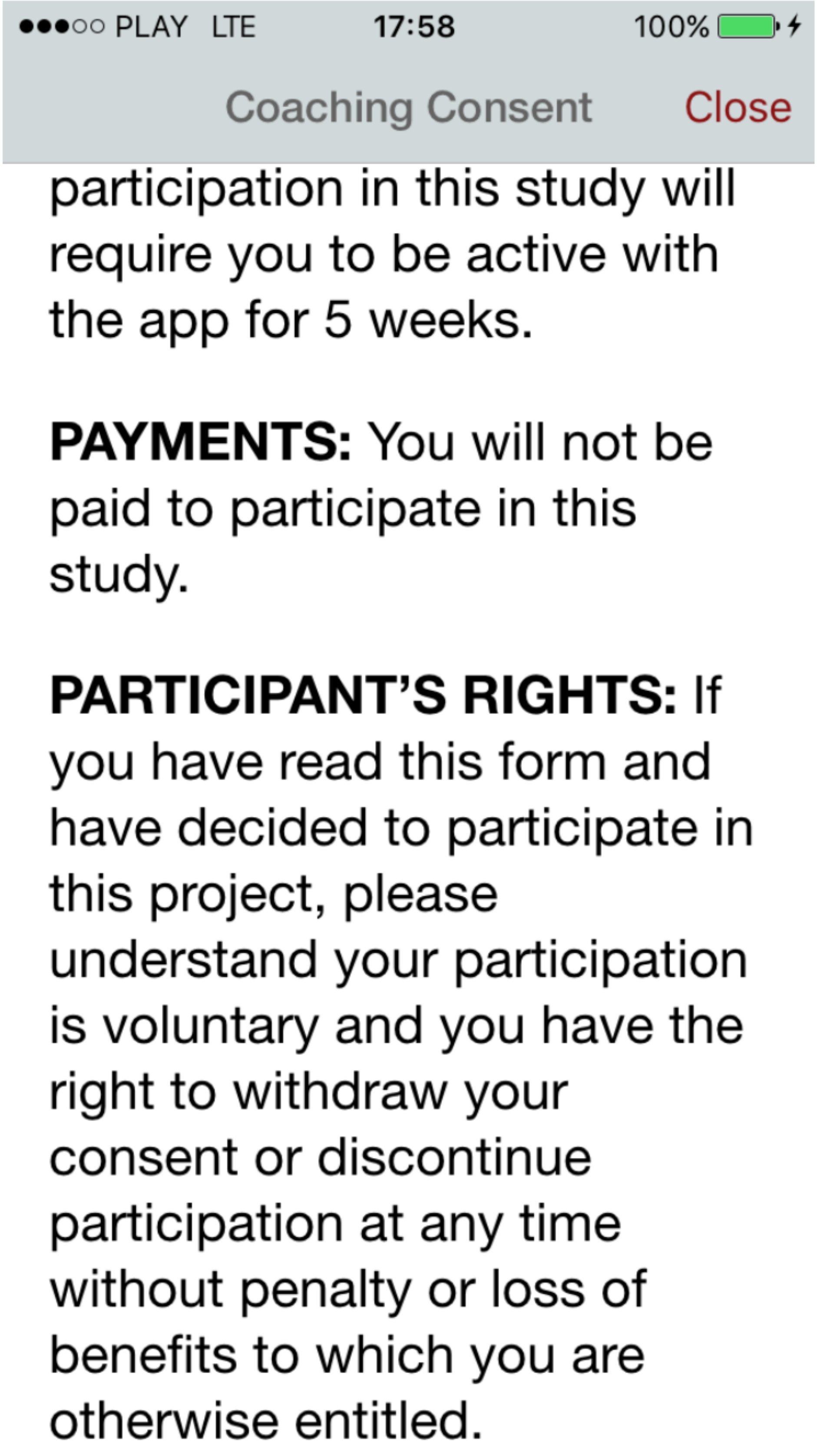
●●●○○ PLAY LTE
17:58
100%
Coaching Consent
Close
participation in this study will require you to be active with the app for 5 weeks.
PAYMENTS: You will not be paid to participate in this study.
PARTICIPANT'S RIGHTS: If you have read this form and have decided to participate in this project, please understand your participation is voluntary and you have the right to withdraw your consent or discontinue participation at any time without penalty or loss of benefits to which you are otherwise entitled.

●●●○○ PLAY LTE 17:58 100%

Coaching Consent Close

benefits to which you are otherwise entitled.
The results of this research study may be presented at scientific or professional meetings or published in scientific journals. However, your identity will not be disclosed.

**CONTACT INFORMATION:**

**Questions, Concerns, or Complaints:** If you have any questions, concerns or complaints about this research study, its procedures, risks and benefits, or alternative courses of treatment, you

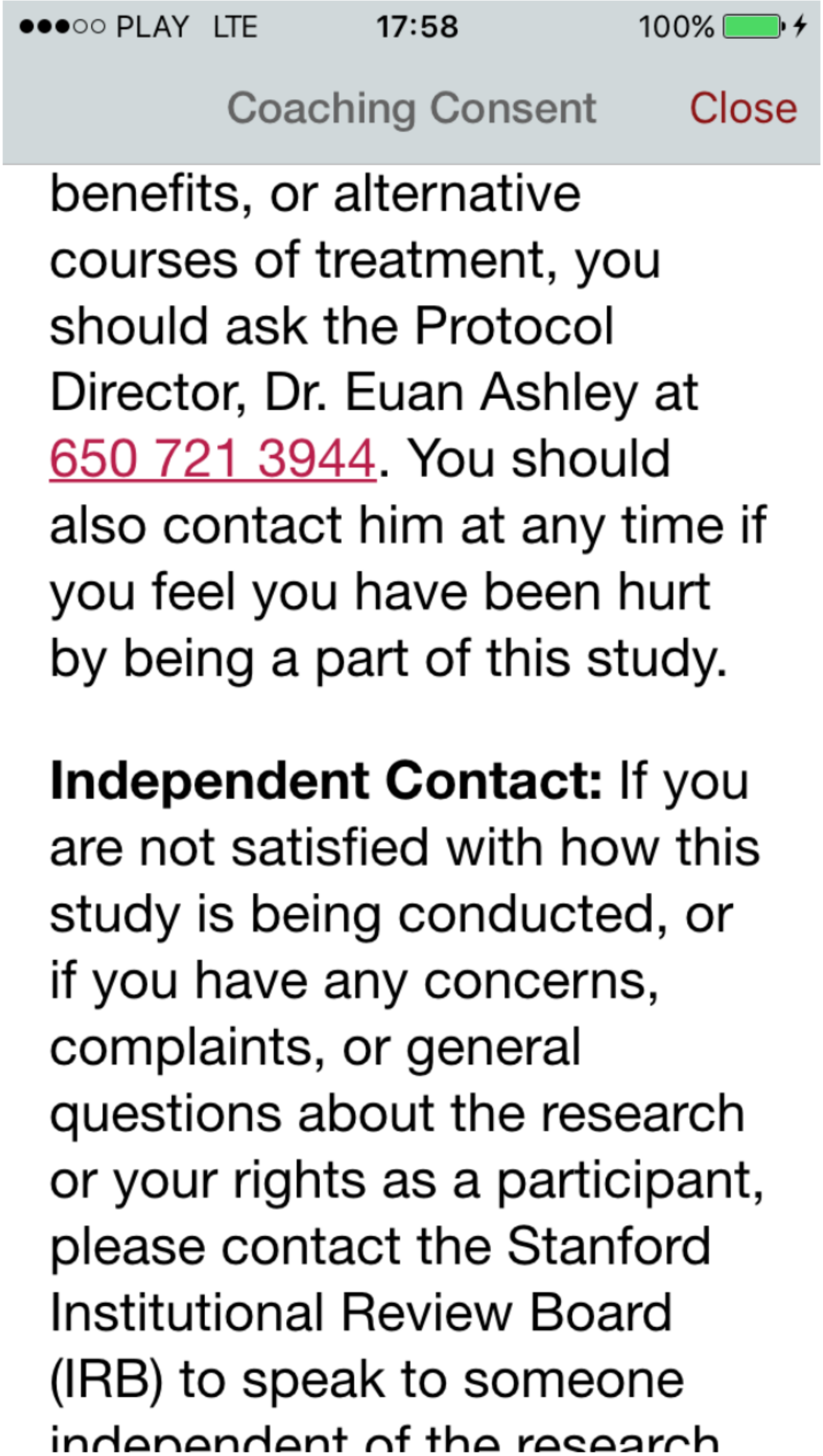
●●●○○ PLAY LTE
17:58
100%
Coaching Consent
Close
benefits, or alternative courses of treatment, you should ask the Protocol Director, Dr. Euan Ashley at 650 721 3944. You should also contact him at any time if you feel you have been hurt by being a part of this study.
Independent Contact: If you are not satisfied with how this study is being conducted, or if you have any concerns, complaints, or general questions about the research or your rights as a participant, please contact the Stanford Institutional Review Board (IRB) to speak to someone
independent of the research

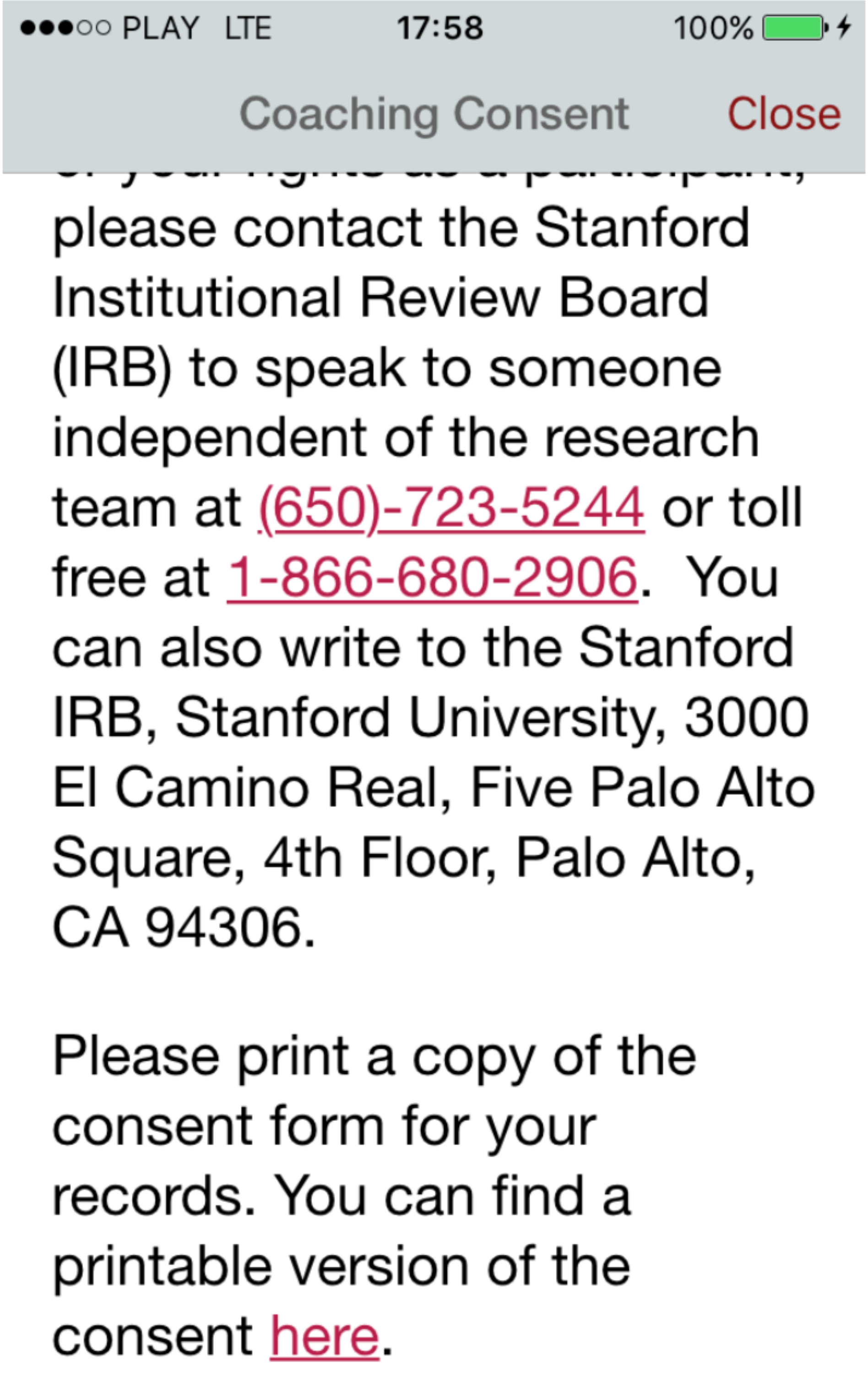

please contact the Stanford Institutional Review Board (IRB) to speak to someone independent of the research team at (650)-723-5244 or toll free at 1-866-680-2906. You can also write to the Stanford IRB, Stanford University, 3000 El Camino Real, Five Palo Alto Square, 4th Floor, Palo Alto, CA 94306.

Please print a copy of the consent form for your records. You can find a printable version of the consent here.

Back to MyHeart-QA 17:35 93%

med.stanford.edu

*IRB Use Only*
Approval Date: Monthname dd, 20yy
Expiration Date: Monthname dd, 20yy

**STANFORD UNIVERSITY Research Consent Form**

Protocol Director: Euan Ashley

Protocol Title: MyHeart Counts: Randomized Assessment of Physical Activity Prompts

**DESCRIPTION:** You are invited to participate in a research study on identifying the best motivational prompts to get people more physically activity.

**PROCEDURES:** With your permission, we would like to collect a baseline week of data (such as the answers to app survey questions, activity data from HealthKit such as step count, heart rate –if available) and then randomly assign you to receive an activity motivating prompt that you will receive through the app and will ask you to complete a very short daily check in survey. The prompts that you will help us test are: 1) a reminder to stand up and move for 60 seconds 2) daily information about the level of activity group that you fall into based on the data collected when you signed up for the app 3) a daily prompt to read about heart health at the American Health Association website and 4) a daily reminder to walk more when you are behind on the 10k steps a day goal. You will receive all four prompts, each prompt will last a week and each week one of the four prompts will randomly be assigned to you.

**RISKS AND BENEFITS:** There are no anticipated risks associated with this study. You will not receive any direct benefit from participation. We cannot and do not guarantee or promise that you will receive any benefits from this study.

**TIME INVOLVEMENT:** Your participation in this study will require you to be active with the app for 5 weeks.

**PAYMENTS:** You will not be paid to participate in this study.

**PARTICIPANT'S RIGHTS:** If you have read this form and have decided to participate in this project, please understand your participation is voluntary and you have the right to withdraw your consent or discontinue participation at any time without penalty or loss of benefits to which you are otherwise entitled.

The results of this research study may be presented at scientific or professional meetings or published in scientific journals. However, your identity will not be disclosed.

**CONTACT INFORMATION:**

**Questions, Concerns, or Complaints:** If you have any questions, concerns or complaints about this research study, its procedures, risks and benefits, or alternative courses of treatment, you should ask the Protocol Director, Dr. Euan Ashley at 650 721 3944. You should also contact him at any time if you feel you have been hurt by being a part of this study.

**Independent Contact:** If you are not satisfied with how this study is being conducted, or if you have any concerns, complaints, or general questions about the research or your rights as a participant, please contact the Stanford Institutional Review Board (IRB) to speak to someone independent of the research team at (650)-723-5244 or toll free at 1-866-680-2906. You can also write to the Stanford IRB, Stanford University, 3000 El Camino Real, Five Palo Alto Square, 4th Floor, Palo Alto, CA 94306.

______I agree to participating in this study

______I do not agree to participate in this study

Please print a copy of the consent form for your records.

Form: SUSampCons-de rev022316 1 of 2

*IRB Use Only*
Approval Date: Monthname dd, 20yy
Expiration Date: Monthname dd, 20yy

**STANFORD UNIVERSITY Research Consent Form**

Protocol Director: Euan Ashley

Protocol Title: MyHeart Counts: Randomized Assessment of Physical Activity Prompts

Back to MyHeart-QA 17:35 93%

med.stanford.edu

**CONTACT INFORMATION:**

**Questions, Concerns, or Complaints:** If you have any questions, concerns or complaints about this research study, its procedures, risks and benefits, or alternative courses of treatment, you should ask [illegible]tor, Dr. Euan Ashley at 650 721 3944. You should also contact him at any time if you [illegible] hurt by being a part of this study.

[illegible]**tact:** If you are not satisfied with how this study is being conducted, or if you have [illegible]nplaints, or general questions about the research or your rights as a participant, please contact the Stanford Institutional Review Board (IRB) to speak to someone independent of the research team at (650)-723-5244 or toll free at 1-866-680-2906. You can also write to the Stanford IRB, Stanford University, 3000 El Camino Real, Five Palo Alto Square, 4th Floor, Palo Alto, CA 94306.

2 of 2

______I agree to participating in this study

______I do not agree to participate in this study

Please print a copy of the consent form for your records.

Form: SUSampCons-dc rev022316

1 of 2

*IRB Use Only*
Approval Date: Monthname dd, 20yy
Expiration Date: Monthname dd, 20yy

**STANFORD UNIVERSITY Research Consent Form**

Protocol Director: Euan Ashley

Protocol Title: MyHeart Counts: Randomized Assessment of Physical Activity Prompts

### IRB TRIAL PROTOCOL

**Title :** MyHeart Counts: Stanford Mobile Cardiovascular Health Study
**Approval Period:** 10/18/2022 - 10/18/2023

**Title :** MyHeart Counts: Stanford Mobile Cardiovascular Health Study
**Approval Period:** 10/18/2022 - 10/18/2023

#### Continuing Review Form

##### 1. Participant Enrollment

**a. Number of participants entered (or number of specimens examined or charts reviewed) since the beginning of study. If this is a combined VA-Stanford study, in addition indicate how many of the participants (or number of VA specimens examined or VA charts reviewed) enrolled with a VA consent. If this is a multi-site study, in addition to the number of participants enrolled locally, include the number of participants enrolled study-wide.**

64,759

**b. Number of: males, females, others or individuals whose sex/gender are unknown or not reported.**

There are 26,192 males, 9,496 females, and 29,071 who did not enter a gender

**c. Minority status of participants entered since beginning of study.**

14 Alaska Natives
95 American Indian
2439 Asian
1086 African American
1578 Hispanic
57 Pacific Islander
18970 Caucasian
553 Other
262 Prefer not to indicate
39705 did not answer this question

**d. Number of children (less then 18 years) entered since beginning of study.**

Children are not enrolled in this study (self-report >18) thus to the best of our knowledge zero.

**e. Number of other potentially vulnerable subjects (if applicable) entered since the beginning of study, including prisoners, pregnant women, economically and educationally disadvantaged, decisionally impaired and homeless people.**

We are not targeting potentially vulnerable subjects as part of this study.

##### 2. Study Problems/Complications

**a. Number of withdrawals of participants from the research (both participant and investigator initiated) since the beginning of the research study. Provide reasons for the withdrawals.**

Sage Bionetworks who handles the consent/withdrawal app transaction does not collect such information, unknown who formally withdrew.

**b. Number of participants lost to follow-up since the beginning of the study.**

We currently have 945 active participants who are sharing data daily with the application.

**c. State if all adverse events have occurred at the expected frequency and level of severity as per study documents or provide a narrative summary of the adverse events since the last continuing review indicating whether the adverse events were expected and/or related to the study.**

There have been no adverse events since the last renewal.

d. **Have there been any unanticipated problems involving risks to participants or others (UPs) since the beginning of the study? A UP must be unexpected (including in severity or frequency), related, AND harmful. Confirm that all UPs have previously been reported to the IRB (guidance GUI-P13).**

There have been no unanticipated problems in the last year.

e. **Provide a narrative summary of all external (e.g. FDA, OHRP, sponsor) audit reports, monitoring visits, inspections, and multi-center trial reports received in the past year. Include corrective actions taken as a result of any audits, inspections, or monitoring visits.**

We've had no reports in the past year.

f. **Complaints about the research in the past year.**

No complaints.

g. **Have there been any instances of noncompliance or deviations since the beginning of the study that have not already been addressed in 2e? Provide a summary and indicate if it has been previously reported to the IRB. Provide a corrective action plan that includes how you will ensure the noncompliance does not recur.**

It was discovered that the agreement with 23andMe had expired and so we are currently refining a new agreement that is backlogging the agreement for data that was collected during the laps of time between agreement expiration and now.

#### 3.Study Assessment

a. **Provide a narrative summary of any interim findings from your data in the past year.**

As Covid still has a large impact on the world, the data is less predictable but activity levels are returning to a more normal state.

b. **Provide a narrative summary of any recent relevant literature.**

NA

c. **Attach Data Safety Monitoring Reports in section 16 received in the past year which have not previously been submitted to the IRB.**

NA

d. **Provide a narrative summary of benefits experienced by participants in the past year.**

It is possible that some app users got more active as a result of signing up with the app or undergoing coaching. Participants feel good helping create out digital biobank.

e. **Provide an assessment of whether the relationship of risks to potential benefits has changed.**

The risk to benefit ratio has not changed since the last revision.

#### 4. Description of remainder of project:

**a.** Y **Is the study open to enrollment?**

**b.** N **Is the study permanently closed to enrollment of new participants?**

**c.** N **Have all participants completed all research-related interventions?**

**d.** Y **Are you still engaged in research-related intervention(s)? If yes, please describe.**

Study is open to new participants to join biobank and undergo coaching. Current participants can share longitudinal data.

**e.** N **Do you wish to renew this study ONLY for long term follow-up?**

**f.** N **Are you ONLY doing data analysis?**

#### 5. Potential Conflict of Interest

N **Is there a change in the conflicting interest status of this protocol?**

#### 6. Protocol Changes

**Please note that if these changes involve changes to Radiation Safety or Biosafety, the IRB will hold its approval until Radiation Safety or Biosafety forwards its approval to the IRB. Use track changes IF revising consent, assent or HIPAA.**

- **Summarize all of the proposed changes to the protocol application including consent form changes.**

  None

- **Indicate Level of Risk**

  No Change

- **Describe any other changes.**

  Admin Contact has changed as Steve Hershman is no longer with Stanford.
  Changed end date in (8m) from (ETA Q1 2023) in Feb 2023 to (ETA Q2 2023) in May 2023.
  Updated status of Alexander Tolas's CITI training. It has been current the entire time but there was a mistake upon initially adding him as personnel.

#### Protocol Director

| **Name** | | **Degree (Program/year if student)** | | **Position, e.g. Assistant Professor, Resident, etc.** |
|---|---|---|---|---|
| Euan Angus Ashley | | M.D. | | Assoc Prof-Med Ctr Line |
| **Department**<br>Medicine - Med/Cardiovascular Medicine | 5406 | **Phone**<br>650 736 1147 | | **E-mail**<br> |
| **CITI Training current** | | | | Y |

#### Admin Contact

| **Name** | | **Degree (Program/year if student)** | | **Position, e.g. Assistant Professor, Resident, etc.** |
|---|---|---|---|---|
| Anders Johnson | | | | Temp Employee |
| **Department** | 5406 | **Phone** | | **E-mail** |

| | | | | |
|---|---|---|---|---|
| Medicine - Med/Cardiovascular Medicine | | 8016984141 | | |
| **CITI Training current** | | | | Y |

#### Investigator

| | | | | |
|---|---|---|---|---|
| **Name** | | **Degree (Program/year if student)** | | **Position, e.g. Assistant Professor, Resident, etc.** |
| **Department** | | **Phone** | | **E-mail** |
| **CITI Training current** | | | | |

#### Other Contact

| | | | | |
|---|---|---|---|---|
| **Name** | | **Degree (Program/year if student)** | | **Position, e.g. Assistant Professor, Resident, etc.** |
| **Department** | | **Phone** | | **E-mail** |
| **CITI Training current** | | | | |

#### Academic Sponsor

| | | | | |
|---|---|---|---|---|
| **Name** | | **Degree (Program/year if student)** | | **Position, e.g. Assistant Professor, Resident, etc.** |
| **Department** | | **Phone** | | **E-mail** |
| **CITI Training current** | | | | |

#### Other Personnel

| | | | | |
|---|---|---|---|---|
| **Name**<br>Alia Crum | | **Degree (Program/year if student)** | | **Position, e.g. Assistant Professor, Resident, etc.**<br>Asst Professor |
| **Department** | | **Phone** | | **E-mail**<br> |
| **CITI Training current** | | | | Y |

| | | | | |
|---|---|---|---|---|
| **Name**<br>Sean Raymond Zion | | **Degree (Program/year if student)** | | **Position, e.g. Assistant Professor, Resident, etc.** |
| **Department**<br>Psychology | 2130 | **Phone** | | **E-mail**<br> |
| **CITI Training current** | | | | Y |

| | | | | |
|---|---|---|---|---|
| **Name**<br>Danielle Zoe Boles | | **Degree (Program/year if student)** | | **Position, e.g. Assistant Professor, Resident, etc.**<br>Soc Science Rsch Coord |
| **Department**<br>Psychology | 2130 | **Phone**<br>(650) 738-4901 | | **E-mail**<br> |
| **CITI Training current** | | | | Y |

**Participant Population(s) Checklist** **Yes/No**

| | |
|---|---|
| • Children (under 18) | N |
| • Pregnant Women and Fetuses | N |
| • Neonates (0 - 28 days) | N |
| • Abortuses | N |
| • Impaired Decision Making Capacity | N |
| • Cancer Subjects | N |
| • Laboratory Personnel | N |
| • Healthy Volunteers | N |
| • Students | N |
| • Employees | Y |
| • Prisoners | N |
| • Other (i.e., any population that is not specified above) | Y |
| • International Participants | Y |

Please enter the countries separated by comma

UK, Hong Kong

#### Study Location(s) Checklist — Yes/No

| | |
|---|---|
| • Stanford University | Y |
| • Clinical & Translational Research Unit (CTRU) | |
| • Stanford Hospital and Clinics | |
| • Lucile Packard Children's Hospital (LPCH) | |
| • VAPAHCS (Specify PI at VA) | |
| • Other (Click ADD to specify details) | |

#### General Checklist

##### Multi-site — Yes/No

- Is this a multi-site study? A multi-site study is generally a study that involves one or more medical or research institutions in which one site takes a lead role.(e.g., multi-site clinical trial) — N

##### Collaborating Institution(s) — Yes/No

- Are there any collaborating institution(s)? A collaborating institution is generally an institution that collaborates equally on a research endeavor with one or more institutions. — N

##### Cancer Institute — Yes/No

- Cancer-Related Studies (studies with cancer endpoints), Cancer Subjects (e.g., clinical trials, behavior/prevention) or Cancer Specimens (e.g., blood, tissue, cells, body fluids with — N a scientific hypothesis stated in the protocol).

| **Clinical Trials** | **Yes/No** |
|---|---|
| • Investigational drugs, biologics, reagents, or chemicals? | N |
| • Commercially available drugs, reagents, or other chemicals administered to subjects (even if they are not being studied)? | N |
| • Investigational Device / Commercial Device used off-label? | Y |
| • IDE Exempt Device (Commercial Device used according to label, Investigational In Vitro Device or Assay, or Consumer Preference/Modifications/Combinations of Approved Devices) | N |
| • Will this study be registered on? clinicaltrials.gov? ( See Stanford decision tree ) | Y |
| • Who will register for ClinicalTrials.gov?<br>NCT# 03090321 | Y |

| **Tissues and Specimens** | **Yes/No** |
|---|---|
| • Human blood, cells, tissues, or body fluids (tissues)? | N |
| • Tissues to be stored for future research projects? | N |
| • Tissues to be sent out of this institution as part of a research agreement? For guidelines, please see https://sites.stanford.edu/ico/mtas | N |

| **Biosafety** (**APB**) | **Yes/No** |
|---|---|
| • Are you submitting a Human Gene Transfer investigation using a biological agent or recombinant DNA vector? If yes, please complete the Gene Transfer Protocol Application Supplemental Questions and upload in Attachments section. | N |
| • Are you submitting a Human study using biohazardous/infectious agents? If yes, refer to the Administrative Panel on BioSafety website prior to performing studies. | N |
| • Are you submitting a Human study using samples from subjects that are known or likely to contain biohazardous/infectious agents? If yes, refer to the Administrative Panel on BioSafety website prior to performing studies. | N |

| **Human Embryos or Stem Cells** | **Yes/No** |
|---|---|
| • Human Embryos or Gametes? | N |
| • Human Stem Cells (including hESC, iPSC, cancer stem cells, progenitor cells) | N |

| **Veterans Affairs** (**VA**) | **Yes/No** |
|---|---|
| • The research recruits participants at the Veterans Affairs Palo Alto Health Care System(VAPAHCS). | N |

**Title :** MyHeart Counts: Stanford Mobile Cardiovascular Health Study
**Approval Period:** 10/18/2022 - 10/18/2023

| | |
|---|---|
| • The research involves the use of VAPAHCS non-public information to identify or contact human research participants or prospective subjects or to use such data for research purposes. | N |
| • The research is sponsored (i.e., funded) by VAPAHCS. | N |
| • The research is conducted by or under the direction of any employee or agent of VAPAHCS (full-time, part-time, intermittent, consultant, without compensation (WOC), on-station fee-basis, on-station contract, or on-station sharing agreement basis) in connection with her/his VAPAHCS responsibilities. | N |
| • The research is conducted using any property or facility of VAPAHCS. | N |

| **Equipment** | **Yes/No** |
|---|---|
| • Use of Patient related equipment? If Yes, equipment must meet the standards established by Biomedical Engineering (BME) (650-725-5000) | N |
| • Medical equipment used for human patients/subjects also used on animals? | N |
| • Radioisotopes/radiation-producing machines, even if standard of care? http://www.stanford.edu/dept/EHS/prod/researchlab/radlaser/Human_use_guide.pdf More Info | N |

| **Payment** | **Yes/No** |
|---|---|
| • Subjects will be paid/reimbursed for participation? See payment considerations. | N |

| **Funding** | **Yes/No** |
|---|---|
| • Training Grant? | N |
| • Program Project Grant? | N |
| • Federally Sponsored Project? | N |
| • https://doresearch.stanford.edu/policies/research-policy-handbook/definitions-and-types-agreements/specialized-categories-sponsored-p Industry Sponsored Clinical Trial? | N |

#### Funding

##### Funding - Grants/Contracts

##### Funding - Fellowships

##### Gift Funding

##### Dept. Funding

**Department Name :** Department of Medicine

##### Other Funding

**Title :** MyHeart Counts: Stanford Mobile Cardiovascular Health Study
**Approval Period:** 10/18/2022 - 10/18/2023

#### Resources :

##### a) Qualified staff.

**Please state and justify the number and qualifications of your study staff.**

Research faculty and staff affiliated with the "Program in Population-Based  Cardiovascular Mobile Health" in the Stanford Biomedical Data Sciences Initiative and Cardiovascular Medicine. Euan Ashley is a cardiologist with expertise in exercise physiology and leads the Stanford Data Sciences Initiative, which includes mobile health data analysis. Collaborators from Alia Crum's lab in phycology were added when we added her Mindset measures as part of a collaboration under the Catalyst umbrella.

##### b) Training.

**Describe the training you will provide to ensure that all persons assisting with the research are informed about the protocol and their research-related duties and functions.**

All staff are hired on the basis of their research experience, data analytics expertise, or software development expertise. They will have further information sessions about the research protocol and their specific duties.

##### c) Facilities.

**Please describe and justify.**

Conventional office space, no wet lab.

##### d) Sufficient time.

**Explain whether you will have sufficient time to conduct and complete the research. Include how much time is required.**

We would like subjects to participate in our mobile cardiovascular health study for a minimum of 1 week every 3 months for at least one year. We anticipate enrolling participants over the course of the year.

##### e) Access to target population.

**Explain and justify whether you will have access to a population that will allow recruitment of the required number of participants.**

We will recruit mobile phone users through their phone, so we will have access to millions of participants.

##### f) Access to resources if needed as a consequence of the research.

**State whether you have medical or psychological resources available that participants might require as a consequence of the research when applicable. Please describe these resources.**

We do not expect significant negative medical or psychological consequences to this research. We do have cardiologists involved in the study and we have a clinical psychologist in CV Medicine.

##### g) Lead Investigator or Coordinating Institution in Multi-site Study.

**Please explain (i) your role in coordinating the studies, (ii) procedures for routine communication with other sites, (iii) documentation of routine communications with other sites, (iv) planned management of communication of adverse outcomes, unexpected problems involving risk to participants or others, protocol modifications or interim findings.**

**Title :** MyHeart Counts: Stanford Mobile Cardiovascular Health Study
**Approval Period:** 10/18/2022 - 10/18/2023

#### 1. Purpose

**a) In layperson's language state the purpose of the study in 3-5 sentences.**

MyHeart Counts is a smartphone-based mobile cardiovascular health research study. It will use the mobile health capabilities of smartphones and wearables to assess daily activity measures of the general public and compare these to measures of cardiovascular health - risk factors and fitness. How people divide their time among exercise, sedentary behavior, and sleep all affect cardiovascular health, yet largely go unmeasured. These can now be measured with sensors in phones or wearable devices. With the large number of smartphone users, we aim to collect activity and cardiovascular health data on many more subjects than in prior studies as well as provide much more quantitative data on type, duration, and intensity of daily activities. It also provides a platform to investigate methods to help participants increase heart-healthy activities. The overall goal is to develop an extensive source of data to help inform future cardiovascular health guidelines.
In addition we previously asked participants who have completed genetic analysis with 23andme if they consent to sharing their genetic data from 23andme with Stanford researchers. We think the opportunity to overlay activity, heart risk and genetic data for participants is a great tool for discovery. App users will have the option of tapping on a 23andMe module that will take them through the informed consent process (screenshots attached in section 16), if they chose to continue they will be taken to a 23andMe portion of the module which asks them to log in with their username and password and then grant permission to share their coded genetic information with Stanford researchers. The Stanford mHealth server (managed by Stanford IRT) pulls that data from an API on the 23andMe server. The participant gets a message saying their genome has been successfully uploaded. A printable copy of the 23andMe module consent is available online and there is a link to that in the app, there is also a copy of the consent that permanently sits in the app as

reference for the participant. While the activity is no longer available, the consent will be.

For Version 2.0 of the mobile app we hope to test out 4 simple coaching/motivational engagement tools via the app in order to better understand
which prompts are more effective than others in getting users to increase their
daily activity as measured by step count as well as mindset about activity as measured by surveys. In addition to which prompt is most
effective we would like to observer whether a particular cluster of individuals will
respond more or less favorably to a particular prompt, thus laying the groundwork
for a personalized coaching tool in the future.

**b) State what the Investigator**(s) **hope to learn from the study. Include an assessment of the importance of this new knowledge.**

With the large number of smartphone users, we aim to collect these data on many
more subjects than in prior studies. This can provide one of the largest databases
on quantitative daily activity and cardiovascular health. This can help us
understand more accurately what types of activity promote cardiovascular health,
which can ultimately help us improve future prevention guidelines.

In the second version of MHC we hope to identify effective and non effective
motivational prompts to increase activity in the MHC user population or to identify
groups of individuals that respond best to a certain prompt (thus setting the stage
for personalized coaching).

**c) Explain why human subjects must be used for this project.** (**i.e. purpose of study is to test efficacy of investigational device in individuals with specific condition; purpose of study is to examine specific behavioral traits in humans in classroom or other environment**)

The purpose of this study is to measure human daily activities and human
cardiovascular health.

#### 2. Study Procedures

**a) Please SUMMARIZE the research procedures, screening through closeout, which the human subject will undergo. Refer to sections in the protocol attached in section 16, BUT do not copy the clinical protocol. Be clear on what is to be done for research and what is part of standard of care.**

The MyHeart Counts research study was designed by Stanford faculty. The app

**Title :** MyHeart Counts: Stanford Mobile Cardiovascular Health Study
**Approval Period:** 10/18/2022 - 10/18/2023

uses features of the Apple HealthKit and ResearchKit toolkits, developed by Apple Inc.

The subject will be consented through their smartphone. After downloading the app from the App Store, the user will be shown a series of screens that explain the general nature of the study and require interaction and acknowledgment of the subject. These smartphone consent process screens (provided as an attachment) have been adapted from an opensource toolkit developed by Sage Bionetworks in collaboration with the Electronic Data Methods forum of the AHRQ (Agency for Healthcare Research & Quality). We have also reviewed this mobile consent process and screenshots with faculty in the Stanford Center for Biomedical Ethics.

After reviewing the consent screens, the user will be shown the consent form and can scroll through and accept prior to enrollment. The consent form will then be emailed to the user.

After consent, subjects will be surveyed about their current cardiovascular health and risk factors and asked to share their activity data collected by the phone and/or any wearable activity device they have. Participants are able to share health and fitness related data with our study by enabling HealthKit permissions on a per-field (ie steps, heart rate, etc) basis. Our study app transmits both historic and newly created HealthKit data to provide a baseline and continuous measure of activity and health.

They will then be asked to use their phone and/or wearable to monitor their daily activity for 1 week. They will then be asked to do a standard 6-minute walk test (with the phone/wearable measuring the distance covered).

Every 3 months for at least one year, subjects will be asked to repeat above, to assess for changes - 1) update surveys/risk factors, 2) monitor activity for 1 week, and 3) 6-minute walk.

After the initial health, activity, and fitness assessment, we plan to randomize a subset of subjects to receive additional information or behavioral motivation tools through the app to determine the effects on activity and risk factors. For example,

**Title :** MyHeart Counts: Stanford Mobile Cardiovascular Health Study
**Approval Period:** 10/18/2022 - 10/18/2023

in addition to receiving their risk assessment by the traditional measure only (American Heart Association 10-year risk of heart disease or stroke, expressed as percentage), subjects will receive the traditional measure plus their risk expressed as a "heart age" (by converting their AHA 10-year risk percentage to the equivalent age of an optimal risk individual). This will assess whether providing a potentially easier-to-understand risk score affects subsequent activity levels and risk factors. Alternatively, they may be asked to use a "heart coach" function through the phone to study the effect on physical activity, engagement, and changes in risk factors.

We also plan to perform a pilot study of up to 50 subjects to test the app prior to release to the general public. We will follow the IRB guidelines for a "pilot study," but because the subject number exceeds 10 we are including the specifics of this pilot study in this protocol, plus our plan to utilize a separate consent. Given the potential for large numbers of the general public to download the app for the main study, it is essential that bugs and usage issues with the app be identified and fixed through this larger pilot study. As per the IRB guidelines, this pilot study will only be "exploratory" in order to help "refine data collection procedures and instruments or prepare a better, more precise research design." Data collected from this pilot study will not be used as research data nor stored permanently. We plan to ask colleagues, including Stanford employees, to use the app for up to 3 months, with periodic contact by research staff for feedback on bugs and other usage issues.

The study is funded by Stanford Medicine, with in-kind software development support for the app from Apple Inc., including participation of Y Media Laboratories (Redwood City), an app development company.

For Version 2.0 of the MHC app the user will be asked to participate for 5 weeks in the study.
- A week of baseline data collection will occur if the user updates to version 2.0

**Title :** MyHeart Counts: Stanford Mobile Cardiovascular Health Study
**Approval Period:** 10/18/2022 - 10/18/2023

and agrees to be part of this trial. The baseline week is the same protocol as
Version 1-1.7 of the app with daily surveys and activity data from phone.

-Following the baseline week the user is randomized to one of the coaching
prompts for a week for 4 weeks. Thus each user is subjected to all 4 prompts in
the study (as outlined in study diagram attached in section 16)

- Step count will be used as a primary outcome to test the effectiveness of the
prompts

For Version 2.2.1 of the MHC app, a feature will be added whereby external
collaborations approved by this IRB and with an active Data Use Agreement will be
able to furnish participants with an "External Identifier" enterable under the
"Profile" tab after completing the consent process.  Sheffield University's
Pulmonary Hypertension Biobank is the only such collaboration (to be approved) at
this time.

Data from MyHeart Counts is only accessible via Syanpse.org by a subset of the
members of the Ashley Lab on this IRB and also by Michael Kellen & Brian Bot from
Sage Bionetworks, our research partner who runs the Bridge Server that receives
the bulk of the app data.  Other members on this IRB within the Ashley lab may
access this data once it is downloaded onto Sherlock.  For external collaborators,
data from individuals who furnished an "External Identifier" from their study will be
periodically downloaded from Synapse and securely sent to the collaborators.

**b) Explain how the above research procedures are the least risky that can be performed consistent with sound research design.**

Using a smartphone and wearable activity monitor for mobile health
carries a low risk. We will also have them do a Physical Activity Readiness
Questionnaire (PAR-Q).

**c) State if deception will be used. If so, provide the rationale and describe debriefing procedures. Since you will not be fully informing the participant in your consent process and form, complete an alteration of consent (in section 13). Submit a debriefing script (in section 16).**

Deception will not be used.

**d) State if audio or video recording will occur. Describe what will become of the recording after use, e.g., shown at scientific meetings, erased. Describe the final disposition of the recordings.**

No.

**e) Describe alternative procedures or courses of treatment, if any, that might be advantageous to the participant. Describe potential risks and benefits associated with these. Any standard treatment that is being withheld must be disclosed in the consent process and form. (i.e. standard-of-care drug, different interventional procedure, no procedure or treatment, palliative care, other research studies).**

None. This does not alter their standard medical treatment.

**f) Will it be possible to continue the more (most) appropriate therapy for the participant(s) after the conclusion of the study?**

This will not alter their standard medical treatment. They can choose to continue to use the mobile health system.

If subjects withdraw from or complete the study, they not be able to use the app anymore. The app's purpose is research, so it would not make sense for subjects to keep using the app when their consent is no longer valid. They would still be able to obtain the individual functions (activity measures, 6 min walk, risk score) through other separate apps that are free or minimal cost.

Additionally, if subjects refuse to allow access to Location Services, Push Notifications, and Core Motion, they will not be able to use the app or participate in the study since key functions of the app depend on allowing access (location for 6 min walk, push notifications to instruct on activity measures and 6 min walk, and core motion for activity measures).

**g) Study Endpoint. What are the guidelines or end points by which you can evaluate the different treatments (i.e. study drug, device, procedure) during the study? If one proves to be clearly more effective than another (or others) during the course of a study, will the study be terminated before the projected total participant population has been enrolled? When will the study end if no important differences are detected?**

This study is designed to compare measures of activity with measures of cardiovascular health, so there are not different medical treatments. We anticipate enrolling subjects for at least a year to generate a large data set that can be analyzed for the many potential subgroups and to gain new insights through "big data" analytics.

Version 2.0- we anticipate to be enrolling individuals continuously for at least a year, the active portion of the study that involves survey completion by user will last 5 weeks. We will analyze data in order to find associations

between groups of
people (clusters) and most effective prompt. If such associations are found we will
ask the IRB permission to do a version 3 study where a longer exposure to the
prompt is studied.

If no important differences are seen in activity levels and prompts we will revise
the
study.

#### 3. Background

a) **Describe past experimental and/or clinical findings leading to the formulation of the study.**

We have experience studying physical activity and cardiovascular health, as well
as developing/studying apps and wearable sensors for cardiovascular health
research and prevention. The development of smartphones as "hubs" of mobile
health data presents a unique opportunity to study measured daily activity and
cardiovascular health in a very broad general population.

Version 2:
We utilized the expertise and advice of Abby King- a world renowned Stanford
researcher on behavior modification to design the version 2 trial. Additionally
Dr.King reviewed all intervention text and provided guidance.

b) **Describe any animal experimentation and findings leading to the formulation of the study.**

N/A

#### 4. Radioisotopes or Radiation Machines

a) **List all standard of care procedures using ionizing radiation (radiation dose received by a subject that is considered part of their normal medical care). List all research procedures using ionizing radiation (procedures performed due to participation in this study that is not considered part of their normal medical care). List each potential procedure in the sequence that it would normally occur during the entire study. More Info**

| Identify Week/Month of study | Name of Exam | Identify if SOC or Research |
|---|---|---|

b) **For research radioisotope projects, provide the following radiation-related information:**

**Identify the radionuclide(s) and chemical form(s).**

**For the typical subject, provide the total number of times the radioisotope and activity will be administered (mCi) and the route of administration.**

**If not FDA approved provide dosimetry information and reference the source documents (package insert, MIRD calculation, peer reviewed literature).**

**c) For research radiation machine projects, provide the following diagnostic procedures:**

**For well-established radiographic procedures describe the exam.**

**For the typical subject, identify the total number of times each will be performed on a single research subject.**

**For each radiographic procedure, provide the setup and technique sufficient to permit research subject dose modeling. The chief technologist can usually provide this information.**

**For radiographic procedures not well-established, provide FDA status of the machine, and information sufficient to permit research subject dose modeling.**

**d) For research radiation machine projects, provide the following therapeutic procedures:**

**For a well-established therapeutic procedure, identify the area treated, dose per fraction and number of fractions. State whether the therapeutic procedure is being performed as a normal part of clinical management for the research participants's medical condition or whether it is being performed because the research participant is participating in this project.**

**For a therapeutic procedure that is not well-established, provide FDA status of the machine, basis for dosimetry, area treated, dose per fraction and number of fractions.**

#### 5. Devices

**a) Please list in the table below all Investigational Devices (including Commercial Devices used off-label) to be used on participants.**

##### 5. 1 Device Name : MyHeartCounts

**Describe the device to be used.**

**MyHeartCounts is a smartphone-based mobile cardiovascular health research study. It will use the mobile health capabilities of smartphones and wearables to assess daily activity measures of the general public and
compare these to measures of cardiovascular health - risk factors and fitness.**

**The MyHeartCounts app was developed by Stanford faculty specifically for this study, using features of the Apple HealthKit development toolkit, and with Y Media Laboratories (Redwood City), an app development company.**

**Manufacturer :** **LifeMap Solutions**

**Risk :** **Non-significant**

Y I confirm the above are true.

**Rationale for the device being non-significant risk:**

Mobile app being used to collect health data

##### Sponsor of Project

**Indicate who is responsible for submitting safety reports to the FDA:**

Y The sponsor is the STANFORD (SU, SHC, LPCH, VA) investigator.

**Please read the following:**

Sponsor-Investigator Research Requirements

**If you would like further information on this process and/or assistance prior to submitting your protocol contact: The Stanford Center for Clinical and Translational Education and Research (Spectrum) at or for cancer research contact:**

Y I have read and understand the above guidance.

**Ordering, Storage and Control**

**To prevent the device being used by a person other than the investigator, and in someone other than a research participant: Confirm that the device will be handled according to the SHC/LPCH policy for Investigational New Devices or as appropriate. If no, please provide an explanation. :**

N Confirm?

The MyHeartCounts app will be available to download from the App Store, but it is not fully functional without the user completing the consent process.

**b) Please list in the table below all IDE Exempt Devices (Commercial Device used according to label, Investigational In Vitro Device or Assay, or Consumer Preference/Modifications/Combinations of Approved Devices) to be used on participants.**

#### 6. Drugs, Reagents, or Chemicals and Devices

**a) Please list in the table below all investigational drugs, reagents or chemicals to be administered to participants.**

**b) Please list in the table below all commercial drugs, reagents or chemicals to be administered to participants.**

#### 7. Medical Equipment for Human Subjects and Laboratory Animals

**If medical equipment used for human patients/participants is also used on animals, describe such equipment and disinfection procedures.**

N/A

#### 8. Participant Population

**a) State the following: (i) the number of participants expected to be enrolled at Stanford-affiliated site(s); (ii)the total number of participants expected to enroll at all sites; (iii) the type of participants (i.e. students, patients with certain cancer, patients with certain cardiac condition) and the reasons for using such participants.**

(i) 400,000; (ii)400,000; (iii) Adult smartphone users, as all adults are at risk for cardiovascular disease. As described earlier, there will be an initial pilot study of up to 50 subjects.

**b) State the age range, gender, and ethnic background of the participant population being recruited.**

Adults (>=18) of all genders, races and ethnicities.

**c) State the number and rationale for involvement of potentially vulnerable subjects in the study (including children, pregnant women, economically and educationally disadvantaged, decisionally impaired, homeless people, employees and students). Specify the measures being taken to minimize the risks and the chance of harm to the potentially vulnerable subjects and the additional safeguards that have been included in the protocol to protect their rights and welfare.**

Colleagues, including employees, will be included only in the pilot study to obtain feedback before release to the general public. We believe this is the best group to obtain feedback from and that it is justifiable given it is a low-risk study with the data collected only for testing, without being stored permanently or included in the main study.

**d) If women, minorities, or children are not included, a clear compelling rationale must be provided (e.g., disease does not occur in children, drug or device would interfere with normal growth and development, etc.).**

Acquired heart disease, such as coronary atherosclerosis, is extremely rare in children. The app requires email verification and entering a date of birth. The date of birth will be used to check that age >=18.

**e) State the number, if any, of participants who are laboratory personnel, employees, and/or students. They should render the same written informed consent. If payment is allowed, they should also receive it. Please see Stanford University policy.**

Up to 50 employees will be included, but only for the pilot study, as described above. We will include a separate consent.

**f) State the number, if any, of participants who are healthy volunteers. Provide rationale for the inclusion of healthy volunteers in this study. Specify any risks to which participants may possibly be exposed. Specify the measures being taken to minimize the risks and the chance of harm to the volunteers and the additional safeguards that have been included in the protocol to protect their rights and welfare.**

We will not specifically study a group of "healthy volunteers." We will study the general public. Note that per NIH/AHA statistics, <10% of the US adult population is at "optimal" (i.e., low) cardiovascular risk, which warrants studying a broad population.

**g) How will you identify and recruit potential participants about the research study? (E.g., by: Honest Broker or other https://researchcompliance.stanford.edu/participantengagement Research Participation services; chart review; treating physician; ads). All final or revised recruitment materials, flyers, etc. must be submitted to the IRB for review and approval before use. You may not contact potential participants prior to IRB approval. See Advertisements: Appropriate Language for Recruitment Material.**

Recruitment will be through the smartphone and open to the "general public." The app will be released initially in the Apple app store with concurrent notice on Stanford website and announced through other media outlets. There will be an informational website set up with the launch of the main study. The app also contains an introductory video.

For the 23andme module:
1) 23andme will email their customers to let them know about the MyHeart Counts app and the ability to give permission within the app to share their 23andme data. A draft of this email has been attached to section 16 of the protocol.

2) For MyHeart Counts participants: the SHC team will include the information regarding 23andme in an email to MHC existing users. The email will be sent out when version 2 of the app is completed, the email will be to inform users of the new exciting features of version 2 and the 23andme material will be one of the exciting new features.

3) For future/new MyHeart Counts users the 23andme module will be in the profile section of the app, the module will pop up after the participant has signed the app consent and will live in the profile tab so if a

participant changes their mind they can access it again. Starting 9/21 we will no longer offer access to the 23andMe module (but will still allow access to the consent document).

For Version 2 release:

1) Stanford University communications office will do a press release and blog article on the release of the enhanced version 2 of MHC
2) The MHC team will email existing users to inform them of the updates and to ask that they update thier app (text of email attached in section 16)
3) We will also put an statement about the upgrades on the MHC app website and the MHC Facebook page
The app is open to the general public thus anyone (who meets inclusion criteria can enroll)

Starting May 2020:
1) Advertisements to be posted on Facebook, Google Adwords and/or Twitter
2) The MHC team will email existing users to inform them of the updates and to ask that they update their app (email attached in section 16)

**h) Inclusion and Exclusion Criteria.**

**Identify inclusion criteria.**

Adult subjects who have a smartphone. The initial version of the research app will utilize Apple's HealthKit, so the subject will need to have an iPhone running iOS8 or later software.

**Identify exclusion criteria.**

Subjects who do not own a smartphone or cannot access their mobile health data. The initial version of the research app will be in English, so patients unable to read English will be excluded.

**i) Describe your screening procedures, including how qualifying laboratory values will be obtained. If you are collecting personal health information prior to enrollment (e.g., telephone screening), please request a waiver of authorization for recruitment (in section 15).**

No additional screening data/procedures required.

**j) Describe how you will be cognizant of other protocols in which participants might be enrolled. Please explain if participants will be enrolled in more than one study.**

Participation in this program should not affect a subject's participation in other research studies as there is no medical or procedural intervention.

**k) Payment/reimbursement. Explain the amount and schedule of payment or reimbursement, if any, that will be paid for participation in the study. Substantiate that proposed payments are reasonable and commensurate with the expected contributions of participants and that they do not constitute undue pressure on participants to volunteer for the research study. Include provisions for prorating payment. See payment considerations**

None.

**l) Costs. Please explain any costs that will be charged to the participant.**

None. The app will be free of charge.

**m) Estimate the probable duration of the entire study. Also estimate the total time per participant for: (i) screening of participant; (ii) active participation in study; (iii) analysis of participant data.**

The entire study will last until shortly after a replacement study is launched on iOS and Android (ETA Q2 2023) in May 2023 when a new iOS is released. We anticipate enrolling subjects continuously until that time. The (i) consent/enrollment is expected to be 10 min, (ii) active participation is expected for 1-5 weeks every 3-6 months for at least 36 months (as above), surveys will take about 5 minutes each of those days and information will be updated every 3 months. We will be piloting these in the pilot study and will have more precise timing from that. If longer than expected we will likely reduce the number of questions, and (iii) data analysis to begin in the first year and continue throughout the study.

#### 9. Risks

a) For the following categories include a scientific estimate of the frequency, severity, and reversibility of potential risks. Wherever possible, include statistical incidence of complications and the mortality rate of proposed procedures. Where there has been insufficient time to accumulate significant data on risk, a statement to this effect should be included. (In describing these risks in the consent form to the participant it is helpful to use comparisons which are meaningful to persons unfamiliar with medical terminology.)

**The risks of the Investigational devices.**

N/A

**The risks of the Investigational drugs. Information about risks can often be found in the Investigator's brochure.**

N/A

**The risks of the Commercially available drugs, reagents or chemicals. Information about risks can often be found in the package insert.**

N/A

**The risks of the Procedures to be performed. Include all investigational, non-investigational and non-invasive procedures (e.g., surgery, blood draws, treadmill tests).**

There will be no change in the participants medical care.

**The risks of the Radioisotopes/radiation-producing machines (e.g., X-rays, CT scans, fluoroscopy) and associated risks.**

N/A

**The risks of the Physical well-being.**

N/A

**The risks of the Psychological well-being.**

N/A

**The risks of the Economic well-being.**

N/A

**The risks of the Social well-being.**

N/A

**Overall evaluation of Risk.**

Low - innocuous procedures such as phlebotomy, urine or stool collection, no therapeutic agent, or safe therapeutic agent such as the use of an FDA approved drug or device.

b) **If you are conducting international research, describe the qualifications/preparations that enable you to both estimate and minimize risks to participants. Provide an explanation as to why the research must be completed at this location and complete the [LINKFORINTERNATIONALREASEARCHFORM] International Research Form. If not applicable, enter N/A.**

this study will take place in the UK and Hong Kong as well

c) **Describe the planned procedures for protecting against and minimizing all potential risks. Include the means for monitoring to detect hazards to the participant (and/or to a potential fetus if applicable). Include steps to minimize risks to the confidentiality of identifiable information.**

There will be no change in the subject's medical care. The mobile health study is low risk. The personal identifiers will be kept to a minimum and will be encrypted and stored separately from the main study data (refer to later privacy and data handling sections).

**d) Explain the point at which the experiment will terminate. If appropriate, include the standards for the termination of the participation of the individual participant Also discuss plans for ensuring necessary medical or professional intervention in the event of adverse effects to the participants.**

The patient can terminate their participation at any time. With the low-risk nature of the study and that it will not involve medical care, we do not foresee any situation that would require termination of the overall study prior to completion.

**e) Data Safety and Monitoring Plan (DSMP). See guidance on Data Safety and Monitoring.**

**A Data and Safety Monitoring Plan (DSMP) is required for studies that present Medium or High risk to participants. (See Overall Evaluation of Risk above). If Low Risk, a DSMP may not be necessary. Multi-site Phase III clinical trials funded by NIH require the DSM Plan to have a Data Safety Monitoring Board or Committee (DSMC or DSMB). The FDA recommends that all multi-site clinical trials that involve interventions that have potential for greater than minimal risk to study participants also have a DSMB or DSMC.**

**The role of the DSMC or DSMB is to ensure the safety of participants by analyzing pooled data from all sites, and to oversee the validity and integrity of the data. Depending on the degree of risk and the complexity of the protocol, monitoring may be performed by an independent committee, a board (DSMC/DSMB), a sponsor's Data Safety Committee (DSC), a Medical Monitor, a sponsor's safety officer, or by the Protocol Director (PD).**

**Describe the following:**

**What type of data and/or events will be reviewed under the monitoring plan, e.g. adverse events, protocol deviations, aggregate data?**

N/A

**Identify who will be responsible for Data and Safety Monitoring for this study, e.g. Stanford Cancer Institute DSMC, an independent monitoring committee, the sponsor, Stanford investigators independent of the study, the PD, or other person(s).**

N/A

**Provide the scope and composition of the monitoring board, committee, or safety monitor, e.g., information about each member's relevant experience or area of expertise. If the Monitor is the Stanford Cancer Center DSMC or the PD, enter N/A.**

N/A

**Confirm that you will report Serious Adverse Events (SAEs), Suspected Unexpected Serious Adverse Reactions (SUSARs), or Unanticipated Problems (UPs) to the person or committee monitoring the study in accordance with Sponsor requirements and FDA regulations.**

N/A

**If applicable, how frequently will the Monitoring Committee meet? Will the Monitoring Committee provide written recommendations about continuing the study to the Sponsor and IRB?**

N/A

**Specify triggers or stopping rules that will dictate when the study will end, or when some action is required. If you specified this in Section 2g [Study Endpoints], earlier in this application enter 'See 2g'.**

N/A

**Indicate to whom the data and safety monitoring person, board, or committee will disseminate the outcome of the review(s), e.g., to the IRB, the study sponsor, the investigator, or other officials, as appropriate.**

N/A

**Select One:**

Y The Protocol Director will be the only monitoring entity for this study.
This protocol will utilize a board, committee, or safety monitor as identified in question #2 above.

#### 10. Benefits

a) **Describe the potential benefit(s) to be gained by the participants or by the acquisition of important knowledge which may benefit future participants, etc.**

There are potential benefits to the participants and to future subjects. While the information about activity, fitness, and cardiovascular risk can be obtained from existing (separate) smartphone apps/devices, participation in this study may help subjects become more informed about their activity, fitness, and cardiovascular risk. The data from the study in the future can improve our understanding of heart-healthy behaviors with quantitative data on the optimal levels of daily activities to promote heart health.

#### 11. Privacy and Confidentiality

##### Privacy Protections

a) **Describe how the conditions under which interactions will occur are adequate to protect the privacy interests of participants (e.g., privacy of physical setting for interviews or data collection, protections for follow-up interactions such as telephone, email and mail communications).**

The interactions with the participant will all be through their smartphone. The participant will need to log in separately to the MyHeart Counts app, beyond just logging into the phone, as an additional privacy protection.

##### Confidentiality Protections

b) **Specify PHI (Protected Health Information). PHI is health information linked to HIPAA identifiers (see above). List BOTH health information AND HIPAA identifiers. If you are using STARR, use the Data Privacy Attestation to ensure that your request will match your IRB-approved protocol. Be consistent with information entered in section 15a.**

The app will obtain the subject's name, date of birth and contact email, primarily for the purpose of validating identity. The app is designed for the general public, so no health care provider, health care facility, or health plan information will be collected directly from providers. The app will survey

participants about demographics (including zip code), any cardiovascular health history and, as described previously, collect data on activity, fitness, and cardiovascular risk. For those individuals who provide consent through the MyHeart Counts app we will upload their coded genetic data from 23andMe and store it on the Stanford mHealth server in coded form. If participants consent to providing their coded genetic data for research use and then later change their mind and wish to withdraw that consent they are instructed to informing us of their wishes. At that point one of the MHC team members will inform Stanford IRT (Who manage the API pull from 23andme) to discard this user code from future API pulls and we will inform the bioinformatician on our team who would be working with the genome data to exclude that genome from planned studies.
HealthKit data may be collected by the application with participants' permissions on a field-by-field basis both for historic and going data for fields including Symptoms, mobility metrics, Active Energy, Blood Glucose, Cycling Distance, Date of Birth, Diastolic Blood Pressure, Flights Climbed, Heart Rate, Height, Oxygen Saturation, Resting Energy, Sex, Sleep Analysis, Stand Hours, Steps, Systolic Blood Pressure, Walking + Running Distance, Weight, Workouts, bloodType, fitzpatricSkinType, lowHeartRateEvent,highHeartRateEvent, irregularHeartRythemEvent, restingHeartRate, heartRateVariabilitySDNN, walkingHeartRateAverage, Heartbeat series, oxygenSaturation, respiratoryRate, vo2Max, mindfulSession, appleStandTime, appleStandHour, bodyTemperature, basalBodyTemperature, bloodType, bloodAlcoholContent, dietaryVitaminD, inhalerUsage, Symptoms, ECG such as from Apple Watch, Mobility Metrics (Walking speed and step length. Walking asymmetry and double support percentage. Stair ascent and descent speed, Six minute walk test distance) and Health Records.
Activity, Health Records and other of the HealthKit fields may include identifiable information if a participant uses information such as their name to name their device (ie "Steve's iPhone" or "Steve's Apple Watch"). When making a public portal, HealthKit and HealthRecord data are either de-identified or to be excluded from the release.

c) **You are required to comply with University Policy that states that ALL electronic devices: computers (laptops and desktops; OFFICE or HOME); smart phones; tablets; external hard disks, USB drives, etc. that may hold identifiable participant data will be password protected, backed up, and encrypted. See http://med.stanford.edu/datasecurity/ for more information on the Data Security Policy and links to encrypt your devices.**

**Provide any additional information on ALL data security measures you are taking. You must use secure databases such as https://researchcompliance.stanford.edu/panels/hs/redcap RedCap. If you are**

**unsure of the security of the system, check with your Department IT representative. Please see http://med.stanford.edu/irt/security/ for more information on IRT Information Security Services and http://www.stanford.edu/group/security/securecomputing/mobile_devices.html for more information for securing mobile computing devices. Additionally,any PHI data on paper must be secured in an locked environment.**

**By checking this box, You affirm the aforementioned.** Y

We have been working closely with Michael Halaas (CIO, Stanford SoM), the SoM IRT team, and Stanford Privacy Office regarding the data security approach. By their determination, our mobile health research study of the general public is not within the typical clinical trial of patients by a health care provider. Thus, they are designating our "Program in Population-based Cardiovascular Mobile Health" overseeing this project as not a "covered entity" and not subject to HIPAA. We will still make every effort to minimize privacy risk for the personal identifiers we are collecting. A secure API will be used to send encrypted data from the smartphone to a data protection and data sharing service overseen by Sage Bionetworks, a non-profit research organization. This has a BRIDGE server that will remove some personal identifiers and send the confidential data to their SYNAPSE server where it can be analyzed and shared with other researchers. A study ID# (healthCode) will be included with the data going to the SYNAPSE server. Thus, the BRIDGE server is the initial location where the main research data are separated from account-related personal identifiers (name, email). Stanford will maintain access to the study ID#. The main research data will leave the BRIDGE server and go to the SYNAPSE server, where it can be accessed by Stanford and its research partners, and if the user opts in, the coded data can also be shared with other qualified researchers who meet criteria determined by Stanford after having all personal identifers including in device names removed. Through the study ID#, only Stanford will have the link of the person and the study data, in order to enable future research where additional health data can be added to the MyHeart Counts data. The participant will be re-contacted for any such request or to participate in other future research opportunities.The Sage Bionetworks data protection/handling

system will undergo security review by Stanford IRT prior to operation
for this
study. As this is not under the auspices of a "covered entity," a BAA
with Sage is
not required. As part of the non-covered-entity Program, the
MyHeartCounts
study will keep all data and activities separate from the covered-
function activities
of the University/SOM. In coordination with SoM CIO Michael Halaas, SoM
IT will
provide a separate infrastructure for the MyHeartCounts data stored at
Stanford
that is not intermingled with infrastructure storing PHI of the Covered
Entity.Note
that we have added language to the consent form regarding use of the app
outside
of the US, advising against use and indicating that US privacy laws may
not protect
their privacy to the extend as the country from which their data may be
originating.
Data from 23andMe will be pulled through an API and stored on the
Stanford
mHealth server, the genetic data will be stripped of identifiers and be
given a
random unique code. It will be stored with this code. Only the
researchers on this
protocol will have access to the code key. The researchers will use the
code to link
back to participants activity data in the mobile app.

**d) Describe how data or specimens will be labeled (e.g. name, medical record number, study number, linked coding system) or de-identified. If you are de-identifying data or specimens, who will be responsible for the de-identification? If x-rays or other digital images are used, explain how and by whom the images will be de-identified.**

As described above, account-related personal identifiers will be removed
by the
BRIDGE server, so the data going to the SYNAPSE server for analysis will
only have
essential identifiers.

For 23andMe genetic data we will assign a random code instead of
using any
identifiers. Only the research staff will have access to they code
key.

**e) Indicate who will have access to the data or specimens (e.g., research team, sponsors, consultants) and describe levels of access control (e.g., restricted access for certain persons or groups, access to linked data or specimens).**

Sage Bionetworks technical staff will operate the BRIDGE servers,
however the
identifiers are encrypted. Stanford researchers will maintain access
to the study
ID#. Sage Bionetworks technical staff will also operate the SYNAPSE
server. The de-identified (of non-essential identifiers) data on the
SYNAPSE server will be accessible

to
Stanford researchers with the participant having the option to share
more broadly,
as described above.

**f) If data or specimens will be coded, describe the method in which they will be coded so that study participants' identities cannot be readily ascertained from the code.**

As mentioned above, a randomly generated study ID# from the BRIDGE
server will
accompany the confidential study data on the SYNAPSE server, with
Stanford
maintaining access to the study ID# and the consent and personal
information.

**g) If data or specimens will be coded, indicate who will maintain the key to the code and describe how it will be protected against unauthorized access.**

The key to link study ID# with personal identifiers will be stored in
the BRIDGE
server accessibly in an unencrypted form only by Stanford staff.

**h) If you will be sharing data with others, describe how data will be transferred (e.g., courier, mail) or transmitted (e.g., file transfer software, file sharing, email). If transmitted via electronic networks, describe how you will secure the data while in transit.See http://www.stanford.edu/group/security/securecomputing/ http://www.stanford.edu/group/security/securecomputing/. Additionally, if you will be using or sharing PHI see https://uit.stanford.edu/security/hipaa https://uit.stanford.edu/security/hipaa.**

The confidential study data will be on the SYNAPSE server described
above.
Stanford researchers will control access. The participant will have
the option to
share these data more broadly, as described above in which
case all
identifiers, including in device names, are removed.  When sending raw
data with collaborators covered under DUA (such as Sheffield) or summary
tables (such as to Sage), data will be put in a secured Box folder.

**i) How will you educate research staff to ensure they take appropriate measures to protect the privacy of participants and the confidentiality of data or specimens collected (e.g. conscious of oral and written communications, conducting insurance billing, and maintaining paper and electronic data)?**

We have already had extensive discussions among project leaders and
staff
regarding both privacy protection and HIPAA rules. This will be
reinforced with all
project staff, researchers we will make sure all are current with
their IRB-related
training.  Mike Halaas, privacy, & security teams have been kept abreast
of our
study updates.

#### 12. Potential Conflict of Interest

Investigators are required to disclose any financial interests that " https://researchcompliance.stanford.edu/eprotocol-coi" target="_blank" reasonably appear to be related/li to this protocol.

Financial Interest Tasks

| Investigators | Role | Potential COI? | Date Financial Interest Answered | Date OPACS Disclosure Submitted | COI Review Determination |
|---|---|---|---|---|---|
| Euan Angus Ashley | PD | N | 09/15/2022 | | N/A |
| Alia Crum | OP | N | 09/30/2022 | | N/A |

#### 13. Consent Background

##### 13. 1 Waiver of Documentation Backbone consent - with UK HK - 2017-2018

**Check if VA related**

a) **Describe the informed consent process. Include the following.**
 i) **Who is obtaining consent?** (**The person obtaining consent must be knowledgeable about the study.**)
 ii) **When and where will consent be obtained?**
 iii) **How much time will be devoted to consent discussion?**
 iv) **Will these periods provide sufficient opportunity for the participant to consider whether or not to participate and sign the written consent?**
 v) **What steps are you taking to minimize the possibility of coercion and undue influence?**
 vi) **If consent relates to children and if you have a reason for only one parent signing, provide that rationale for IRB consideration.**

i,ii: The consent will be obtained through the smartphone by taking the subject through multiple screens describing the study (see attachment). iii,iv,v: The subject can take their time going through the smartphone consent process, so they should have sufficient opportunity to consider participation/consent and there will be no coercion. vi: N/A

b) **What is the Procedure to assess understanding of the information contained in the consent? How will the information be provided to participants if they do not understand English or if they have a hearing impairment? See HRPP Chapter12.2 for guidance.**

The subject will have to go through each screen of the consent process and asked to review the consent form and agree to the consent on the phone. In the initial version of the app we are excluding non-English participants. Hearing ability is not required to go through the consent process and perform the study tasks

c) **What steps are you taking to determine that potential participants are competent to participate in the decision-making process? If your study may enroll adults who are unable to consent, describe** (**i**) **how you will assess the capacity to consent,** (**ii**) **what provisions will be taken if the participant regains the capacity to consent,**(**iii**) **who will be used as a legally authorized representative, and** (**iv**) **what provisions will be made for the assent of the participant.**

Even though this is a low-risk study, we are not aiming to recruit subjects who are not competent to consent. As mentioned, the subject will have to go through each screen of the consent process and review the consent form and agree to the consent on the phone.

**Select ALL applicable regulatory criteria for a Waiver of Documentation and provide a protocol-specific**

**justification:**

**1)** **45 CFR 46.117(c)(1)(i)., that the only record linking the participants and the research would be the consent document, and the principal risk would be potential harm resulting from a breach of confidentiality; each participant (or legally authorized representative) will be asked whether he/she wants documentation linking the participant with the research, and the participant's wishes govern.**

**2)** **45 CFR 46.117(c)(1)(ii)., that the research presents no more than minimal risk of harm to participants and involves no procedures for which written consent is normally required outside of the research context.**

**3)** **45 CFR 46.117(c)(1)(iii)., if participants or legally authorized representatives (LAR) are members of a distinct cultural group in which signing forms is not the norm, the research presents no more than minimal risk and there is an appropriate alternative mechanism for documenting that informed consent was obtained.**

**4)** Y **21 CFR 56.109(c)(1)., presents no more than minimal risk of harm to participants and involves no procedures for which written consent is normally required outside of the research context.**

**Rationale for above selection:**

The consent is being obtained through a smartphone, so documentation of signature is not practical. There are still multiple steps the user must follow in order to consent to the study and be enrolled.

#### 13. 2 Waiver of Documentation edited Coaching Module Consent - 2017-2018

**Check if VA related**

**a)** **Describe the informed consent process. Include the following.**

- **i)** **Who is obtaining consent? (The person obtaining consent must be knowledgeable about the study.)**
- **ii)** **When and where will consent be obtained?**
- **iii)** **How much time will be devoted to consent discussion?**
- **iv)** **Will these periods provide sufficient opportunity for the participant to consider whether or not to participate and sign the written consent?**
- **v)** **What steps are you taking to minimize the possibility of coercion and undue influence?**
- **vi)** **If consent relates to children and if you have a reason for only one parent signing, provide that rationale for IRB consideration.**

i,ii: The consent will be obtained through the smartphone by taking the subject through multiple screens which include the verbatim consent form language (see attachment 23andMe consent module in section 16). iii,iv,v: The subject can take their time going through the smartphone consent process, so they should have sufficient opportunity to consider participation/consent and there will be no coercion. vi: N/A. The subject will be able to download the the consent form (attached in section 13) when they get to the last module. After the consent process is complete, a summary of the modules will remain in the smartphone for future reference

**b)** **What is the Procedure to assess understanding of the information contained in the consent? How will the information be provided to participants if they do not understand English or if they have a hearing impairment? See HRPP Chapter12.2 for guidance.**

The subject will have to go through each screen of the consent process and asked to perform certain actions to indicate understanding. They will also need to review the consent form and agree to the consent on the phone. In the initial version of the app we are excluding non-English participants. Hearing ability is not required to go through the consent process and perform the study tasks.

**c)** **What steps are you taking to determine that potential participants are competent to participate in the decision-making process? If your study may enroll adults who are unable to consent, describe (i) how you will assess the capacity to consent, (ii) what provisions will be taken if the participant regains the capacity to consent,(iii) who will be used as a legally authorized representative, and (iv) what provisions will be made for the assent of the participant.**

Even though this is a low-risk study, we are not aiming to recruit subjects who are not competent to consent. As mentioned, the subject will have to go through each screen of the consent process and will be asked to perform certain actions to indicate understanding. They will also need to review the consent form and agree to the consent on the phone.

**Select ALL applicable regulatory criteria for a Waiver of Documentation and provide a protocol-specific justification:**

1) **45 CFR 46.117(c)(1)(i)., that the only record linking the participants and the research would be the consent document, and the principal risk would be potential harm resulting from a breach of confidentiality; each participant (or legally authorized representative) will be asked whether he/she wants documentation linking the participant with the research, and the participant's wishes govern.**

2) **45 CFR 46.117(c)(1)(ii)., that the research presents no more than minimal risk of harm to participants and involves no procedures for which written consent is normally required outside of the research context.**

3) **45 CFR 46.117(c)(1)(iii)., if participants or legally authorized representatives (LAR) are members of a distinct cultural group in which signing forms is not the norm, the research presents no more than minimal risk and there is an appropriate alternative mechanism for documenting that informed consent was obtained.**

4) Y **21 CFR 56.109(c)(1)., presents no more than minimal risk of harm to participants and involves no procedures for which written consent is normally required outside of the research context.**

**Rationale for above selection:**

The consent is being obtained through a smartphone, so documentation of signature is not practical. There are still multiple steps the user must follow in order to consent to the study and be enrolled.

#### 13. 3 Waiver of Documentation Consent for UK with GDPR

**Check if VA related**

a) **Describe the informed consent process. Include the following.**
- i) **Who is obtaining consent? (The person obtaining consent must be knowledgeable about the study.)**
- ii) **When and where will consent be obtained?**
- iii) **How much time will be devoted to consent discussion?**
- iv) **Will these periods provide sufficient opportunity for the participant to consider whether or not to participate and sign the written consent?**
- v) **What steps are you taking to minimize the possibility of coercion and undue influence?**
- vi) **If consent relates to children and if you have a reason for only one parent signing, provide that rationale for IRB consideration.**

i,ii: The consent will be obtained through the smartphone by taking the subject through multiple screens describing the study (see attachment). iii,iv,v: The subject can take their time going through the smartphone consent process, so they should have sufficient opportunity to consider participation/consent and there will be no coercion. vi: N/A

b) **What is the Procedure to assess understanding of the information contained in the consent? How will the information be provided to participants if they do not understand English or if they have a hearing impairment? See HRPP Chapter12.2 for guidance.**

The subject will have to go through each screen of the consent process and asked to review the consent form and agree to the consent on the phone. In the initial version of the app we are excluding non-English participants. Hearing ability is not required to go through the consent process and perform the study tasks

c) **What steps are you taking to determine that potential participants are competent to participate in the decision-making process? If your study may enroll adults who are unable to consent, describe (i) how you will assess the capacity to consent, (ii) what provisions will be taken if the participant regains the capacity to consent,(iii) who will be used as a legally authorized representative, and (iv) what provisions will be made for the assent of the participant.**

Even though this is a low-risk study, we are not aiming to recruit subjects who are not competent to consent. As mentioned, the subject will have to go through each screen of the consent process and review the consent form and agree to the consent on the phone.

**Select ALL applicable regulatory criteria for a Waiver of Documentation and provide a protocol-specific justification:**

1) **45 CFR 46.117(c)(1)(i)., that the only record linking the participants and the research would be the consent document, and the principal risk would be potential harm resulting from a breach of confidentiality; each participant (or legally authorized representative) will be asked whether he/she wants documentation linking the participant with the research, and the participant's wishes govern.**

2) **45 CFR 46.117(c)(1)(ii)., that the research presents no more than minimal risk of harm to**

**Title :** MyHeart Counts: Stanford Mobile Cardiovascular Health Study
**Approval Period:** 10/18/2022 - 10/18/2023

**participants and involves no procedures for which written consent is normally required outside of the research context.**

**3)** **45 CFR 46.117(c)(1)(iii)., if participants or legally authorized representatives (LAR) are members of a distinct cultural group in which signing forms is not the norm, the research presents no more than minimal risk and there is an appropriate alternative mechanism for documenting that informed consent was obtained.**

**4)** Y **21 CFR 56.109(c)(1)., presents no more than minimal risk of harm to participants and involves no procedures for which written consent is normally required outside of the research context.**

**Rationale for above selection:**

The consent is being obtained through a smartphone, so documentation of signature is not practical. There are still multiple steps the user must follow in order to consent to the study and be enrolled.

#### 14. Assent Background (less than 18 years of age)

#### 15. HIPAA Background

#### 16. Attachments

| Attachment Name | Attached Date | Attached By | Submitted Date |
|---|---|---|---|
| scientific_scholarly_review form | 11/10/2014 | joannec2 | |
| Draft slides of MyHeart Counts website | 02/20/2015 | mvm | |
| Updated CV Health survey | 02/25/2015 | mvm | |
| Updated Diet survey | 02/25/2015 | mvm | |
| Updated Well-Being and Risk survey | 02/25/2015 | mvm | |
| Updated Activity and Sleep survey | 02/25/2015 | mvm | |
| Physical Activity Readiness Questionnaire | 02/25/2015 | mvm | |
| Final script for intro video | 03/02/2015 | mvm | |
| MHC screen shots 3/2/15 | 03/02/2015 | mvm | |
| Facebook page screen shots | 03/05/2015 | apavlovi | |
| March 5th 2015 Website Screenshots | 03/05/2015 | apavlovi | |
| MHC Press release | 03/05/2015 | apavlovi | |

**Title :** MyHeart Counts: Stanford Mobile Cardiovascular Health Study
**Approval Period:** 10/18/2022 - 10/18/2023

| | | |
|---|---|---|
| support page | 03/06/2015 | aeslinge |
| Justification for International Data use | 03/25/2015 | apavlovi |
| Email 1 to Participants | 05/27/2015 | apavlovi |
| 1.5 MHC edits July 2015 | 07/21/2015 | apavlovi |
| Newsfeed Screenshot | 07/21/2015 | apavlovi |
| UK HK US Privacy policy | 10/19/2015 | apavlovi |
| 23andme module info | 12/07/2015 | apavlovi |
| 23andme collab agmt | 12/07/2015 | apavlovi |
| Examples of Sleep/Happines feedback graphs | 02/18/2016 | apavlovi |
| 23andmeapi diagram | 04/12/2016 | apavlovi |
| correspondence with M. Halaas April 21st 2016 | 04/21/2016 | apavlovi |
| 23and Me Modules | 04/29/2016 | manjit |
| updated_irb_text-2 june 7th | 06/08/2016 | manjit |
| Study Design for Version 2 of MHC | 06/20/2016 | apavlovi |
| Day by Day Coaching Protocol | 06/20/2016 | apavlovi |
| AHA License Agreement | 06/20/2016 | apavlovi |
| Announcement email to users for MHV 2.0 | 06/20/2016 | apavlovi |
| Steps graph learn more text | 06/20/2016 | apavlovi |
| Vegetable intake graph learn more text | 06/20/2016 | apavlovi |
| Screenshots Version 2.0 | 06/20/2016 | apavlovi |
| Coaching Module- IRB revisions | 08/10/2016 | apavlovi |
| New and Returning Users Consent Module Diagram | 08/10/2016 | apavlovi |
| Coaching Consent Wireframes | 08/29/2016 | apavlovi |
| coaching_consent_screenshots SEP 8 | 09/08/2016 | sstolar |
| Recruitment email 9_13_16 | 09/13/2016 | manjit |
| 20170913 Upgrade section | 09/13/2017 | hershman |

| | | |
|---|---|---|
| added to coaching_consent | | |
| Sep 2017 Email Current users | 09/13/2017 | hershman |
| Sep 2017 Email New Users | 09/13/2017 | hershman |
| Sep 2017 Feedback Survey | 09/13/2017 | hershman |
| App Store Text (2.0.2) | 09/13/2017 | hershman |
| 23andMe Spring 2018 outreach | 04/12/2018 | hershman |
| 2018_04 Feedback Survey 2 | 04/12/2018 | hershman |
| Smoking History | 09/04/2018 | hershman |
| Process Mindset Inventory 10.25.18ocx | 12/06/2018 | hershman |
| Adequacy of Activity Mindset Measure 10.25.18 | 12/06/2018 | hershman |
| Illness Mindset Inventory - 5.1.18 | 12/06/2018 | hershman |
| External Identifier | 02/13/2019 | hershman |
| STH-Obs Research Tissue Bank Protocol IRB Approved | 02/13/2019 | hershman |
| eConsent (and with GDPR) | 09/04/2019 | hershman |
| COVID-19 hg Survey + Modifications | 04/01/2020 | hershman |
| AdvertCOVIDJoined | 04/22/2020 | hershman |
| MHC+COVID-19 hg modifications & repeated for 2.3.2 | 04/22/2020 | hershman |
| May 2020 Email to current users | 05/01/2020 | hershman |

#### Obligations

The Protocol Director agrees to:

- Adhere to principles of sound scientific research designed to yield valid results
- Conduct the study according to the protocol approved by the IRB
- Be appropriately qualified to conduct the research and be trained in Human Research protection, ethical principles, regulations, policies and procedures
- Ensure all Stanford research personnel are adequately trained and supervised
- Ensure that the rights and welfare of participants are protected including privacy and confidentiality of data

- Ensure that, when de-identified materials are obtained for research purposes, no attempt will be made to re-identify them.
- Disclose to the appropriate entities any potential conflict of interest
- Report promptly any new information, modification, or unanticipated problems that raise risks to participants or others
- Apply relevant professional standards.

Any change in the research protocol must be submitted to the IRB for review prior to the implementation of such change. Any complications in participants or evidence of increase in the original estimate of risk should be reported at once to the IRB before continuing with the project. Inasmuch as the Institutional Review Board (IRB) includes faculty, staff, legal counsel, public members, and students, protocols should be written in language that can be understood by all Panel members. The investigators must inform the participants of any significant new knowledge obtained during the course of the research.

IRB approval of any project is for a maximum period of one year. For continuing projects and activities, it is the responsibility of the investigator(s) to resubmit the project to the IRB for review and re-approval prior to the end of the approval period. A Notice to Renew Protocol is sent to the Protocol Director 7 weeks prior to the expiration date of the protocol.

https://stanfordmedicine.box.com/shared/static/qbsi8u8h47qsotxhdpuzz50xlrqa0sgo.pdf Report promptly any new information, complaints, possibly serious and/or continuing noncompliance, or unanticipated problems involving risks to participants or others.

All data including signed consent form documents must be retained for a minimum of three years past the completion of the research. Additional requirements may be imposed by your funding agency, your department, or other entities. (Policy on Retention of and Access to Research Data, Research Policy Handbook, http://doresearch.stanford.edu/policies/research-policy-handbook/conduct-research/retention-and-access-research-data)

APPROVAL LETTER/NOTICE NOTE: List all items (verbatim) that you want to be included in your approval letter (e.g., Amendment date, Investigator's Brochure version, consent form(s) version(s), advertisement name, etc.) in the box below.

Y By checking this box, I verify that I, as the Protocol Director (PD) responsible for this research protocol, have read and agree to abide by the above obligations, or that I have been delegated authority by the PD to certify that the PD has read and agrees to abide by the above obligations.
